# Early detection of pancreatic ductal adenocarcinomas with an ensemble learning model based on a panel of protein serum biomarkers

**DOI:** 10.1101/2021.12.02.21267187

**Authors:** Nuno R. Nené, Alexander Ney, Tatiana Nazarenko, Oleg Blyuss, Harvey E. Johnston, Harry J. Whitwell, Eva Sedlak, Aleksandra Gentry-Maharaj, Eithne Costello, William Greenhalf, Ian Jacobs, Usha Menon, Justin Hsuan, Stephen P. Pereira, Alexey Zaikin, John F. Timms

**Affiliations:** Department of Women’s Cancer, EGA Institute for Women’s Health, University College London, 84-86 Chenies Mews, London, WC1E 6HU, UK; Institute for Liver and Digestive Health, University College London, Upper 3rd Floor, Royal Free Campus, Rowland Hill Street, London NW3 2PF, UK; Department of Mathematics, University College London, London WC1H 0AY, UK; Department of Applied Mathematics, Lobachevsky University of Nyzhniy Novgorod, Nizhniy Novgorod 603105, Russia; School of Physics, Engineering & Computer Science, University of Hertfordshire, Hatfield, UK; Babraham Institute, Babraham Research Campus Cambridge, CB22 3AT, UK; National Phenome Centre and Imperial Clinical Phenotyping Centre, Department of Metabolism, Digestion and Reproduction, IRDB Building, Imperial College London, Hammersmith Campus, London, W12 0NN, UK; Section of Bioanalytical Chemistry, Division of Systems Medicine, Department of Metabolism, Digestion and Reproduction, Imperial College London, South Kensington Campus, London, SW7 2AZ, UK; MRC Clinical Trials Unit at UCL, Institute of Clinical Trials and Methodology, UCL, 90 High Holborn, 2nd Floor, London, WC1V 6LJ, UK; Department of Molecular and Clinical Cancer Medicine, University of Liverpool, Liverpool, UK; Liverpool Experimental Cancer Medicine Centre, University of Liverpool, Liverpool, L69 3GL, UK; University of New South Wales, Sydney, NSW, 2052, Australia; World-Class Research Center “Digital biodesign and personalized healthcare”, Sechenov First Moscow State Medical University, Moscow, Russia

## Abstract

Earlier detection of pancreatic ductal adenocarcinoma (PDAC) is key to improving patient outcomes, as it is mostly detected at advanced stages which are associated with poor survival. Developing non-invasive blood tests for early detection would be an important breakthrough. The primary objective of the work presented here was to use a unique dataset, that is both large and prospectively collected, to quantify a set of 96 cancer-associated proteins and construct multi-marker models with the capacity to accurately predict PDAC years before diagnosis. The data is part of a nested case control study within UK Collaborative Trial of Ovarian Cancer Screening and is comprised of 219 samples, collected from a total of 143 post-menopausal women who were diagnosed with pancreatic cancer within 70 months after sample collection, and 248 matched non-cancer controls. We developed a stacked ensemble modelling technique to achieve robustness in predictions and, therefore, improve performance in newly collected datasets. With a pool of 10 base-learners and a Bayesian averaging meta-learner, we can predict PDAC status with an AUC of 0.91 (95% CI 0.75 - 1.0), sensitivity of 92% (95% CI 0.54 - 1.0) at 90% specificity, up to 1 year to diagnosis, and at an AUC of 0.85 (95% CI 0.74 - 0.93) up to 2 years to diagnosis (sensitivity of 61%, 95 % CI 0.17 - 0.83, at 90% specificity). These models also use clinical covariates such as hormone replacement therapy use (at randomization), oral contraceptive pill use (ever) and diabetes and outperform biomarker combinations cited in the literature.

## Introduction

Pancreatic ductal adenocarcinoma (PDAC) is associated with dismal 5-year survival rates (∼3-7%) and is projected to become the second cause of cancer death by 2030 ^1–3^. A non-specific clinical course leading to late-stage diagnosis is a feature of pancreatic cancer, and only 15% of the patients are diagnosed at early stages with resectable tumours ^2–5^. Following surgery and adjuvant therapy however, less than 30% of patients survive 5 years after diagnosis ^5^, compared with a less than 10% 5 year survival in those with unresectable disease ^6^. The development of new tests that could improve detection of early-stage disease is pivotal for improving outcomes for pancreatic cancer patients. Indeed, it has been shown that if tumour size at detection can be reduced from 3 to 2 cm, improved oncological resection rates (7% to 83%, respectively) and increased median survival (from 7.6 to 17.2 months) can be achieved ^7, 8^. CA19-9 ^9, 10^ is the only serological tumour marker used routinely for the detection and monitoring of PDAC progression, however, with 79-81% test sensitivity and 82-90% specificity for diagnosis of PDAC at best ^11^. Despite this, we have recently shown that CA19-9 (and CA125) can be used to detect pancreatic cancer up to 2 years before clinical diagnosis, using samples from a repository collected as part of the UK Collaborative Trial of Ovarian Cancer Screening (UKCTOCS) ^12^. These samples, which were prospectively and longitudinally collected months and years prior to diagnosis, enabled the detection of rising levels of potential serological biomarkers ahead of diagnosis with high reliability. We proposed that CA19-9 in combination with markers identified in this cohort and their use in multi-marker longitudinal algorithms, may improve test performances and lead time to enable early detection of PDAC. Combining markers into multi-marker models has traditionally involved the application of simple cut-off rules and machine learning methods such as multivariate logistic regression ^12^, neural networks and support vector machines ^13–15^. Here, to attain better performances and robustness, we applied an ensemble modelling technique ^16–19^ and a repeated cross-validation re-sampling strategy. Ensemble methods have been immensely successful in producing accurate predictions for many complex classification tasks ^16, 18, 20^, because they address fundamental problems in data analysis. For example, these methods avoid overfitting by combining single learners with a local search heuristic which decreases the risk of obtaining a local performance minimum. Issues surrounding dimensionality are also addressed with ensemble models. By allowing each classifier to focus on sub-spaces of features, the burden of large search spaces is reduced. This specific field in machine learning is consistent with the well-known Condorcet’s jury theorem, which states that if each classifier has a probability larger than 0.5 of being correct then increasing the pool of classifiers increases the probability of making the correct decision by majority voting. The task of finding successful ensembles is, nevertheless, more complex and dependent on a balance between diversity and consensus among classifiers. A definitive recipe to achieve this goal has yet to be completely defined ^16, 18, 20^. Two widely cited examples following the ensemble paradigm that focus on complementary and heterogeneity are, for instance, stacking, a form of meta-learning ^18^, and ensemble selection ^16^. Stacking constructs a higher-level predictive model over the predictions of base classifiers. Ensemble selection, on the other hand, uses an incremental strategy to select base predictors for the ensemble while balancing diversity and performance ^16^. Here we will apply the stacking approach due to its simplicity and computational efficiency. Ensemble like modelling has also been used in the design of the CancerSEEK multi-cancer blood test, which assesses blood levels of multiple analytes such as circulating proteins and cell free DNA mutations ^21^, but also in the creation of indices evaluating changes in DNA methylation patterns ^22^. The use of multi-datasets and multi-platform integration in pancreatic cancer studies ^23^ are essential for early detection and aligns at a fundamental level with the data analysis methodology applied in our work. Here, we demonstrate how using a stacked ensemble approach which relies on a panel of 20 features, including cancer-associated proteins and clinical covariates, outperforms state-of-the-art multi-biomarker combinations previously applied in pancreatic ductal adenocarcinoma early detection.

## Results

### Data set characteristics

Our dataset is part of a nested case control study within UK Collaborative Trial of Ovarian Cancer Screening (UKCTOCS) ^24, 25^ and is comprised of 143 individuals with PDAC and 248 controls (see Materials and Methods). 35 of the PDAC-diagnosed patients provided longitudinal samples, with between 2 and 6 annual samples per individual collected prior to diagnosis. These samples were considered to be independent for the purpose of data analysis and were divided into a training (2/3) set and test (1/3) set, by stratifying for age, hormone replacement therapy (HRT) use at randomization, oral contraceptive pill (OCP) use (ever), diabetes status (detailed information about duration and type was not available for this work), body-mass index (BMI) quartile, PDAC or control status and for sample single time-group, i.e., 0-1,1-2,2-3,3-4 and 4+ years to diagnosis (YTD), attributed to each sample and determined by the time to diagnosis at sample collection (compare Table 1 with Tables S2 and S3, see also Table S4 for the total number of cases and controls per single time-group). This stratification enabled a clearer evaluation of PDAC classifier panel performances in collected samples not used in training, i.e., the test set, and ensured that the results are realistic and representative, and are not biased by data or information leakage ^26^.

**Table 1.** Study data set description. P values were calculated according to a logistic regression model with a bias reduction method (see Methods), for the whole sample distribution. For the same variables in the subsets used as training and test sets see Tables S2 and S3. This table corresponds to taking all samples from all time-groups, i.e., 0 to 4+ years to diagnosis. See also Figs. S3 and S4.

| Variable | Cases | Controls | P value |
| --- | --- | --- | --- |
| No. individuals | 143 | 248 |  |
| No. samples | 219 | 248 |  |
| Tumour site |  |  |  |
| tail | 10 | na |  |
| body | 22 | na |  |
| body/tail | 3 |  |  |
| head | 88 | na |  |
| unspecified | 95 | na |  |
| Mean time to spin (h) (range) | 21.7 (0.48 - 46.53) | 21.82 (0.32 - 46.24) | 0.93 |
| Mean age at sample draw (yr) (range) | 64.94 (51.19 - 74.87) | 62.48 (50.44 - 76.86) | < 0.0001<br>(OR= 1.06) |
| Mean BMI (kg/m2) (range) | 27.46 (17.80 - 43.74) | 26.64 (17.91 - 44.39) | 0.078 |
| Mean time from sample collection to diagnosis (months) (range) | 26.07 (0.99-70.09) | na |  |
| HRT use (at randomisation) |  |  |  |
| yes | 19 | 48 | 0.0010 |
| no | 199 | 201 | (OR = 0.41) |
| OCP use (ever) |  |  |  |
| yes | 132 | 127 | 0.039 |
| no | 86 | 122 | (OR = 1.47) |
| Diabetes |  |  |  |
| yes | 42 | 11 | <0.0001 |
| no | 176 | 238 | (OR = 4.99) |

Time intervals between sample collection and serum/plasma isolation, i.e., time to spin, were comparable between PDAC cases and controls. There was no significant difference in the mean time to spin between cases and controls for the whole study data set (Table 1), with the ranges being also very similar. The same is observed in the training and test sets (Tables S2 and S3). The distribution of ages at sample draw showed a significant association (OR=1.06, *P* < 0.0001) with PDAC status, with cases having a mean value at 64.94 years and controls at 62.48 years. Once again, a similar observation regarding significance can be made for the training and test sets. In both, we verify odds-ratios favouring PDAC status (Tables S2 and S3). Through further analysis in the training set, we verified that age at sample draw was, nevertheless, only significantly associated with PDAC in training samples obtained 4+ years prior to diagnosis (Fig. S1 and S3A). Moreover, by applying the logistic regression model used for the ranking of individual features which was developed in the training set (see Methods), we observed that age did not generate significant AUCs in the test set, and the sensitivities were very low (Fig. S1). To qualify as a predictive marker, subject age had to be combined as a co-variate in a multi-marker model as is reported below (see also Supplementary Information for an extensive study on single and typical biomarker combinations in simple state-of-the-art logistic regression models). As a single variate, subject BMI was neither a significant predictor in the entire data set (Table 1) nor was it in the training and test sets (Tables S2, S3 and Fig. S1C and D). Regarding HRT, it was significant in all data sets (Tables 1, S2 and S3). In the training set, this result stemmed mostly from the association of HRT use with a lower risk of pancreatic cancer (OR =0.22 < 1, 95% CI 0.04-0.85, *P* = 0.027, coloured in blue in Fig. S1E) in the 2-3 single time-group samples. Its significant predictive potential for distinguishing between cases and controls, however, was not reproducible in the test set 2-3 time-group by applying a logistic regression model developed in the training set, AUC=0.56 (95% CI 0.50-0.67) (Fig. S1F). Similarly to age, HRT was expected to add value to a PDAC predictive index as a covariate. Although demonstrating an association with PDAC status across the entire study set (Table 1) and the training set (Table S2), OCP was not a significant predictor in any of the single time-group samples evaluated independently (Fig. S1E). Overall, diabetes was the strongest predictor of risk for PDAC among the clinical covariates (Table 1 and Table S2). While consistent across both the entire study set and our training set in all single time-groups excluding the 1-2 YTD samples, the largest risk was in fact observed in the 3-4 YTD subgroup (OR=13.26 (95% CI 1.36 - 1781.23), *P* =0.022) and 4+ YTD (OR=8.43 (95% CI 1.64 - 84.86), *P* = 0.0090). These findings were however not reproducible in our test set despite the proportions pertaining to OCP being roughly the same as those in the training (Fig. S1E, F and Tables S2 and S3). Once again, diabetes had to be combined with other co-variates to enhance performances (see section below on ensemble stacking of multi-marker models).

Another aspect of the data that should be reported is the presence of outliers with respect to CA19-9. Despite not having been confirmed, we assumed that Lewis-antigen negative or positive status for each individual could be determined by the lack of expression of CA19-9, verified by a peak at the minimum value in the data used here. Since CA19-9 is not expressed in 8-10% of Caucasians (Lewis-negative blood group), predictive indices should account for this phenomenon. The proportion of individuals in which CA19-9 was not detected was approximately the same in the training and test sets and, apart from in samples belonging to the 0-1 single time-group, there was no significant association with PDAC status in the training set (Fig. S5). Furthermore, logistic regression models, trained to distinguish cases from controls, based on the presence or absence of measurable CA19-9, did not perform well in the test set, with performances always around AUCs of 0.5 and lacked statistical significance (Fig. S5).

### Ensemble learning multi-marker models improve performance and robustness

The group of base-learners chosen for the ensemble analysis reported here covered a large and diverse set of approaches, which was beneficial, as different characteristics of the training set were captured by different classifier techniques (see comments below on variable importance attributed by each classifier). This, in turn, will increase classifier heterogeneity and therefore the likelihood of success when predicting outcomes in unseen data ^16–19^. Here, we will focus on the results of the JTG2L ensemble classifier, which was trained on all samples collected 0-1, 0-2, 0-3, 0-4 or 0-4+ YTD (described in the Methods section), since it was the most robust. For the results on other stacking ensemble strategies see the Supplementary Information section on ensemble classifiers specialized in single time-group samples.

As we increased the interval of joined time-groups from 0-1 to 0-4+, and thus the number of samples used to train, the performances in the test set decreased (when the training and testing time-groups are the same), as expected, since the median time to diagnosis increased (see Fig. 1B and the diagonal values of the heatmap in Fig. 1C). A similar trend was roughly observed for the sensitivity, positive predictive value (PPV) and negative predictive value (NPV) achieved (see Fig. 1D to F), with the 0-2 group appearing as the outlier. The use of the JTG2L Bayesian Model Averaging (BMA) stacked ensemble was beneficial both for improving performance as well as decreasing variability across folds (Fig. 1A). Models trained in each joined time-groups attained, nevertheless, better performances in the test set, in certain instances, when evaluated in samples belonging to narrower time-groups, e.g. AUC^test^_(0-3)_ (0-3,training)=0.79 was considerably smaller than AUC^test^_(0-3)_ (0-4+,training)=0.84, with the difference being borderline statistically insignificant (*P*=0.06) (Fig. 1C). This result probably stemmed from the additional non-specific information contained in the single 3-4 and 4+ time-groups, which helped to correct for the poor performances in the 1-2 single time-group (Fig. S28). Under normal circumstances, when evaluating the PDAC risk for samples in newly collected data, early diagnosis would only be a comprehensive effort if evaluated with the classifier trained with 0-4+ joined time-group samples, as time to diagnosis is obviously not available for data collected from new patients. Reassuringly, the cross time-group performances in the test set, generated with the 0-4+ classifier, were not statistically different from those with narrower joined time-groups (*P^test^*_(0-1)_ (0-4+,training)=0.66, *P^test^*_(0-2)_ (0-4+,training)=0.68, *P^test^*_(0-3)_ (0-4+,training)=0.06, *P^test^*_(0-4)_ (0-4+,training)=0.41, when comparing the last column in Fig. 1C with the diagonal), under this broad classifier, which justifies its use in external validation sets. In addition, the JTG2L BMA ensemble approach outperformed the single marker and multi-marker models relying on simple logistic regression (Fig. S1 and Fig. S2); trained with 0-4+ samples and evaluated in the 0-1 subgroup of the test set, it reached an AUC^test^_(0-1)_(0-4+,training)=0.94 (95% CI 0.83 - 1), which far exceeded the results with CA19-9 alone or in combination with THBS2, MUC16 or CEACAM5 when predicting PDAC status in the same time-group (see Fig. S1 and Fig. S2 and Supplementary Information on single and multi-marker combination models developed with simple logistic regression techniques). The sensitivity at 90% specificity and the respective PPV and NPV were also enhanced for the JTG2L BMA ensemble model. An improvement in performance with respect to previous studies in pre-diagnostic samples was also achieved with samples collected up to 2 years; there a sensitivity of 0.406 at 0.905 specificity was reported ^12^. The JTG2L BMA stack was capable of attaining a performance of 0.84 (0.72 - 0.93), when trained in samples belonging to the same joined time-group, i.e., 0-2, or 0.85 (95% CI 0.74-0.93), when trained in 0-4+ samples, the difference between the two was insignificant (*P^test^*_(0-2)_ (0-4+,training)=0.68). The use of the larger feature input space characterising the JTG2L BMA stack is further justified given that the significant single marker models in the training set (Fig. S11) did not reach significance in the test set in 0-2 samples, with exception of CA19-9, with an AUC^test^_(0-2)_ < 0.75. A previously published logistic regression model combining CA19-9 and MUC16 gave AUCs of 0.76 for the up to 1 year group in a previous study validation set ^12^. In the current data set, the same bi-dimensional model predicted PDAC with an AUC^training^_(0-1)_ (CA19-9, MUC16) = 0.81 (95% CI 0.72 - 0.88), and AUC^training^_(0-2)_ (CA19-9, MUC16)=0.74 (95% CI 0.67-0.81), in the training set (reported in Supplementary Information). The same models tested at roughly the same level in unseen data, here the test set: AUC^test^_(0-1)_ (CA19-9, MUC16) = 0.76 (95% CI 0.55 - 0.93) and AUC^test^_(0-2)_ (CA19-9, MUC16) = 0.66 (95% CI 0.51-0.80) (see Supplementary Information and Fig. S2), the latter considerably lower than the JTG2L BMA stack performance of 0.84 (trained in 0-2 samples, *P*= 0.0060) and 0.85 (trained in 0-4+ samples, *P*= 0.0061) (see Fig. 1C). The sensitivity, PPV and NPV at 90% specificity also improved with the algorithm proposed here, with sensitivities as high as 0.92 (95% CI 0.54 - 1), 0.61 (95% CI 0.17 - 0.83) and 0.63 (95% CI 0.29 - 0.77), and PPVs as high as 0.91 (95% CI 0.85-0.92), 0.81 (95% CI 0.55-0.85) and 0.84 (95% CI 0.70-0.87), in 0- 1, 0-2 and 0-3 test samples, depending on the joined/combined time-group used to train (Figs. 1D to 1I). This outcome came at a cost of increased model complexity and input space dimensionality. The features selected from the 101-marker input panel are plotted in Fig. 2 and the enriched terms are presented in Fig. 3. The selected panel of biomarkers for the JTG2L BMA stack meta-learner developed with all time-group samples combined (0-4+), shows typical gene ontology and pathway terms with ‘Constitutive signalling by aberrant PI3K in cancer’ [REAC], ‘Pancreatic adenocarcinoma pathway’ [WP], ‘Pathways in cancer’ and ‘Pancreatic cancer and proteoglycans in cancer’ [KEGG], being significantly over-represented (Fig. 3).

**Fig. 1.**
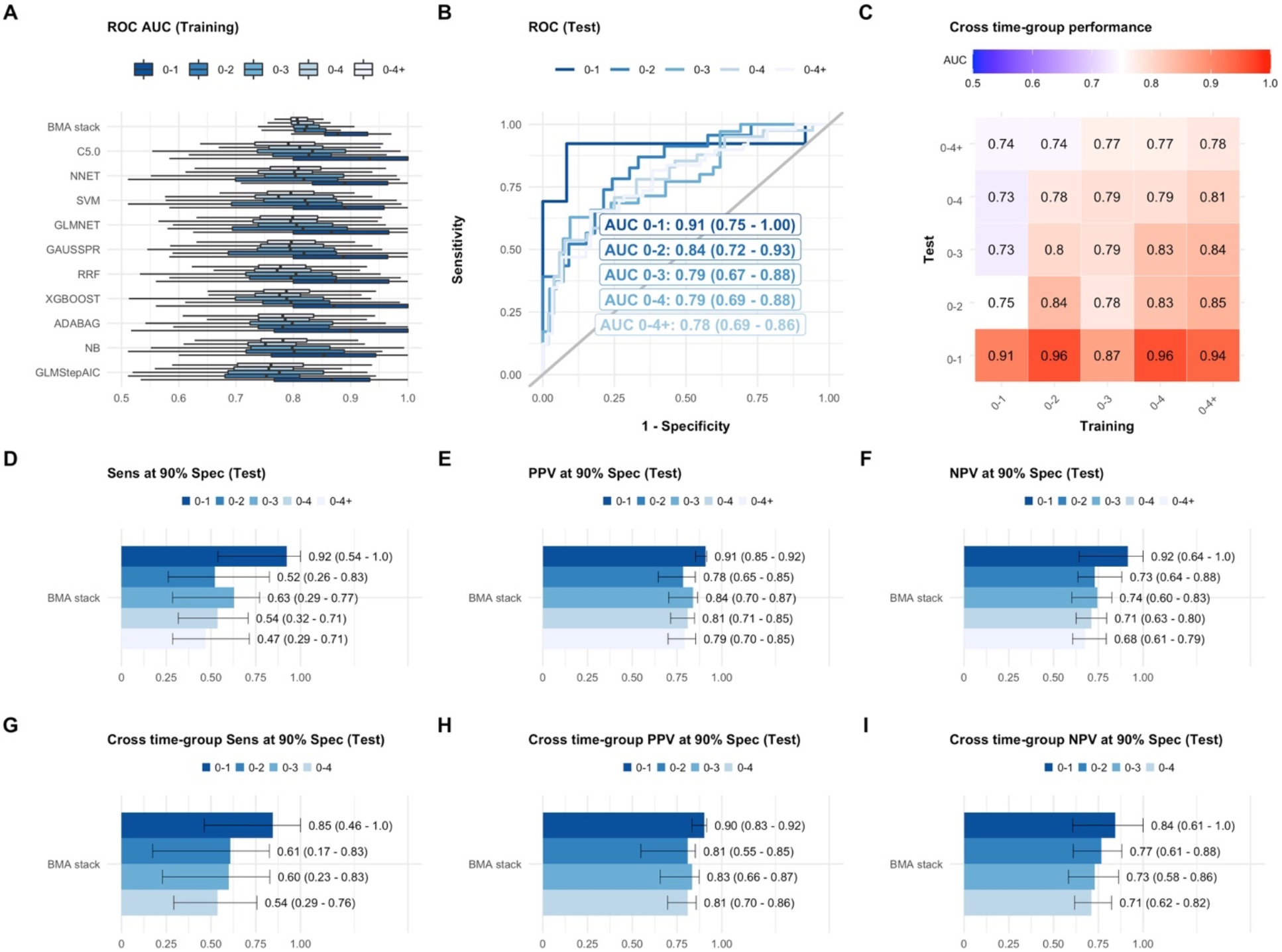
Ensemble model performance per joined/combined time-group. **A** Distribution of ROC AUCs across training folds for each of the base-learners and the BMA stack meta-learner (JTG2L, see Methods). See also Figs. S20 to S23 for alternative stacking methods**. B** Receiver Operating Characteristic (ROC) curves in the test set for the BMA stack per joined time-group. AUC 95% CI were determined by stratified bootstrapping. **C** Cross-time group performance of BMA stack developed in the training set and evaluated in specific time-groups in the test set. 95% CI for AUCs are not shown but the predictions were all significant**. D** Sensitivity**, E** Positive predictive value (PPV) and **F** Negative predictive value (NPV) at 90% specificity. **G, H** and **I** Cross time-group performances for the ensemble trained in 0-4+ samples (last column in C).

**Fig. 2.**
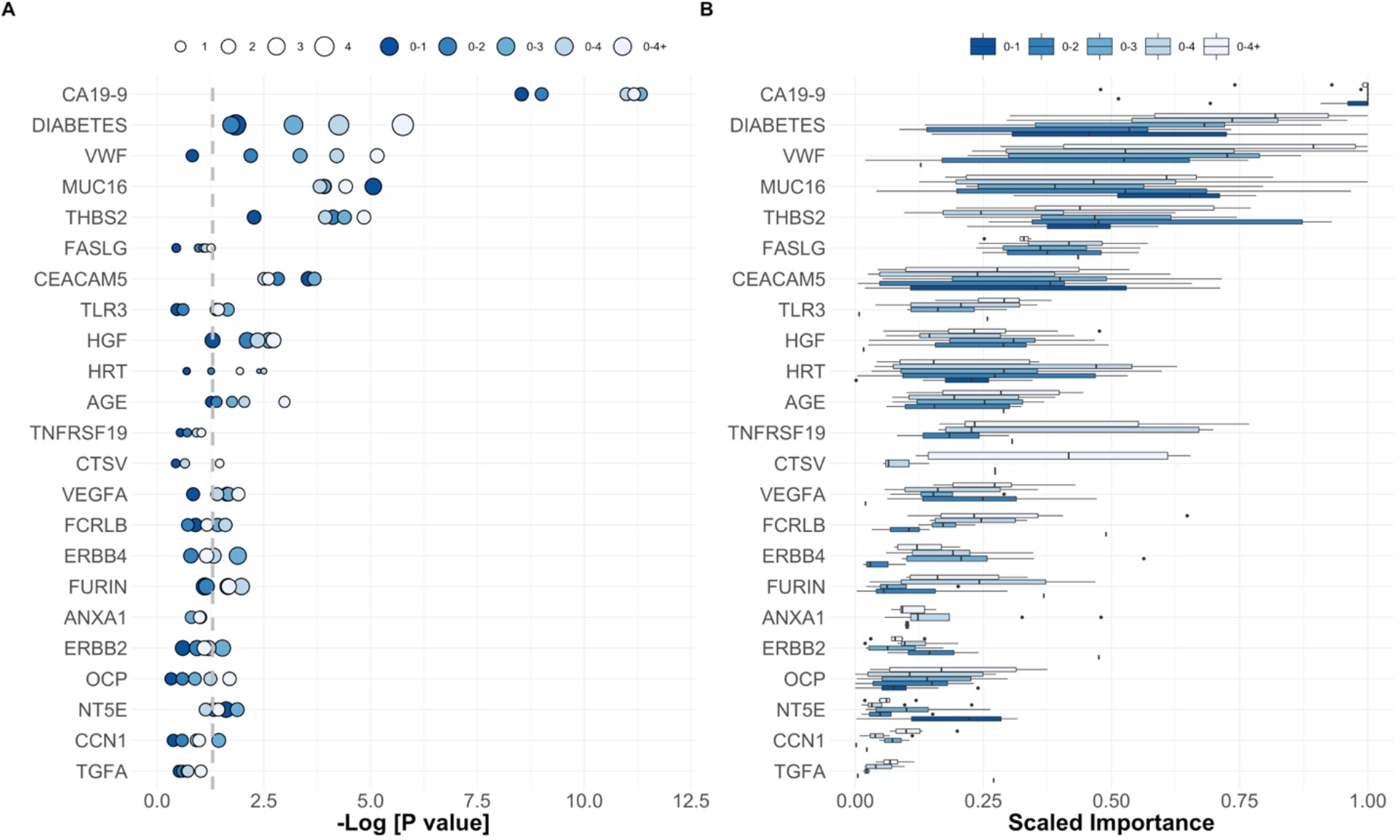
Feature importance across PDAC base-learner signatures. **A** Odds-ratios and P-values for the ranking procedure according to a logistic regression model using Firth’s bias reduction method (see Methods). **B** Feature importance (see Methods) across all base learners and joined time-groups. All the features presented in this figure were selected when training with 0-4+ samples. The importance plotted for the remaining joined time-groups is the importance of each feature in their respective models. See also Fig. S26 for the full plots.

**Fig. 3.**
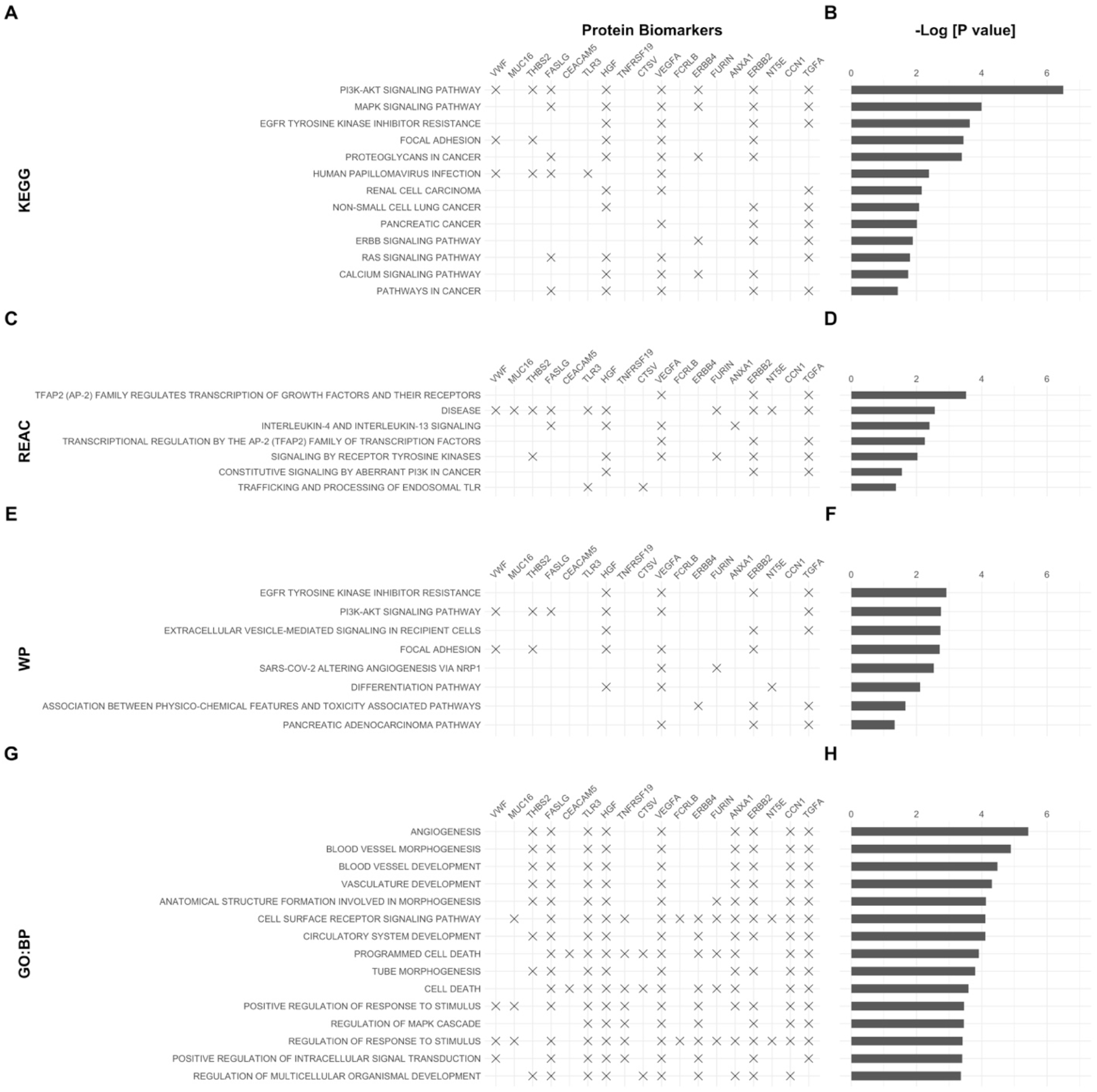
Enrichment analysis. g:Profiler terms for the set of features selected for the optimal classifier trained in 0-4+ samples. **A** Kyoto Encyclopaedia of Genes and Genomes (KEGG) pathways. **C** Reactome Pathway Database (REAC). **E** WikiPathways (WP). **G** Gene ontology terms biological process (GO: BP). The respective adjusted p-values associated with each enrichment term or pathway are plotted in **B**, **D**, **F** and **H**. See also Fig. 2.

One of the striking aspects of the 0-4+ PDAC predictive index was the presence of the commonly cited markers such as CA19-9, MUC16, THBS2, CEACAM5 and VWF and 4 of the clinical variables, excluding BMI. Diabetes was ranked just below CA19-9 according to the median importance across base-learners and time-groups, something which is consistent with what is observed in Fig. S2. Also remarkable was the variance in the importance attributed to the same markers across models developed in narrower time-groups, except for CA19-9 which almost always was allocated a scaled importance of 1. These observations are in line with those reported in the literature as strong arguments for using stacking and ensembles of classifiers, e.g., a pool of base-learners outperforms single classifiers by enabling heterogeneity within the pool of the base-learners and thus robustness in the predictions ^16, 18^. There is, nevertheless, a caveat to this as diversity and performance are not strictly directly proportional and while there is a strong dependency between the former and the latter, diversity might hinder performance of the ensemble ^18, 27^. Given that we started with a reduced pool of base-learners, the true relationship between heterogeneity/diversity and performance could not be analysed extensively. Yet, upon searching all possible combinations of base-learners from the 10-dimensional input space, the use of all 10 classifiers in the stack always outperformed the rest across a set of 10 times 10-fold cross-validation resampling strategy. Although there wasn’t a clear trend for base-learner pair-wise diversity with time-group across base-learners (see Cohen k-statistic in Fig. S17), the JTG2L BMA stack did, in fact, outperform the best base-learner in the training set (Fig. 1A), and any of the remaining stacks (see Fig. S22 and S23). This was particularly clear in the trend observed among stacks from best to worse in Fig.S22A. The BMA stack is the obvious choice across time-groups and, consequently, constitutes the model of choice for validation in new data. It was interesting to note that the MEAN stack outperforms the GEOMEAN stack, a mixture of experts focusing on consensus among base-learners, and the MAX stack, choosing the maximum probability for PDAC status among the base-learners, which, effectively, amounts to highlighting the classifier that has the highest degree of certainty for each sample. The BMA stack and the MEAN stack provide a balance between base-learners, the former being weighted, which increased the performance in the test set, especially in the 0-1 and 0-2 joined time-groups.

Another prominent trend in the importance attributed to each marker was the fact that the more significant a marker is (Fig. 2A), the higher the variance in importance it registered across base-learners (Fig. 2B, see also Fig. S26). By observing the association of each of the markers with increased (OR > 1) or decreased (OR < 1) risk for PDAC (Fig. S6-S15), we also verified that the ensemble trained on 0-4+ samples had a greater proportion of OR <1 markers than, for instance, the ensemble trained on 0-2 samples.

Regarding the importance of developing a classifier that is reliant on features covering both the population that expresses CA19-9 (Lewis-positive) and does not express CA19-9 (Lewis-negative), we also tested the BMA stack model in the subset of subjects in the test set that are Lewis-positive. The number of Lewis-negative subjects was not sufficient to generate significant performances. Overall, the BMA stacked ensemble was not affected substantially and a performance in the test set of AUC=0.89 (95% CI 0.70 - 1.00) in 0-1 samples and of AUC= 0.83 (95% CI 0.70 – 0.93) in 0-2 samples was obtained. This confirms that the association between the CA19-9 peak at the level associated with absence of expression and PDAC was not affecting the results. Further tests in larger pools of confirmed Lewis-negative samples should clarify the performance of the ensembles developed here.

One particular aspect of PDAC is its low prevalence in the general population, at 8–12 per 100 000 per year and a 1.3% lifetime risk of developing the disease ^28^. The models developed and tested here relied on data where the prevalence was approximately between 40% to 50% (see Table 1 and Table S4) and on the maximization of the ROC AUC, which is independent from prevalence. At these values, the BMA stack JTG2L reached PPVs as high as 0.91 (95% CI 0.85 – 0.92) in 0-1 samples and 0.84 (95% CI 0.70 – 0.87) in 0-3 samples (Fig. 1E). If the prevalence of the disease in the test set was changed the PPV and NPV at 90% specificity decreased and increased, respectively, at lower disease prevalence, as expected (Fig. S25) ^29^. The ROC AUC was nevertheless stable and the tendency with time-groups observed in Fig. 1 was also verified (see Fig. S24). The larger variances at lower prevalence stem from the smaller datasets.

## Discussion

From the classifiers developed here, those trained with all available samples (0-4+) would be the most appropriate under a clinical setting, as time to diagnosis is not available in newly collected data. The features highlighted as predictive under these circumstances included important and widely referenced markers which we confirmed as having among the strongest association with PDAC, namely CA19-9, CEACAM5, MUC16 (CA125), THBS2 and diabetes. Compared to the rest, CA19-9 was the most prominent marker as all base-learner classifiers attributed it almost always the highest importance. CA19-9 is an indicator of aberrant glycosylation in pancreatic cancer and it is considered as a biomarker, predictor, and promoter in pancreatic cancer, although it is often found elevated in benign pancreatic biliary diseases such as pancreatitis, cholangitis and obstructive jaundice, giving a significant rise in false positives ^11, 30^. Moreover, CA19-9 expression is absent in 8-10% of Caucasians with a Lewis-negative blood group, as the CA19-9 epitope is in fact, the sialylated Lewis A blood group antigen ^11^. Although CA19-9 levels are routinely used in detection, determination of resectability and monitoring of PDAC progression ^31, 32^, its low predictive value and a low prevalence of pancreatic cancer in the general population exclude it as a robust screening tool^3, 33^. The combination of CA19-9 with other markers proposed here is, therefore, advantageous. Regarding CEACAM5 and the role of carcinoembryonic (CEA) related cell adhesion molecules (CEACAMs) 1, 5 and 6 in progression of solid tumours (such as colorectal, lung, melanoma, breast, liver) including pancreatic cancer is well established, and their expression varies between different tumour histological subtypes ^34–36^. With respect to pancreatic cancer, CEACAM5 has been widely described as having a variable diagnostic value in PDAC detection, while its expression has also been reported to inversely correlate with disease stage ^37^. Pancreatic cyst fluid levels (>192 ng/mL) of CEACAM5 are also predictive of mucinous rather than serous cystic lesions ^36, 38–41^, and endoscopic sampling with fluid CEA measurement is recommended by international guidelines for identification of malignant transformation ^42–44^. In a recent report, CEACAM5 was found to be persistently elevated up to 26.5 months prior to pancreatic cancer diagnosis, in a large cohort (n=1196) of longitudinally sampled subjects ^45^. The predictive performance of CEACAM5 as a single analyte in pancreatic cancer however, is poor ^35^. In our work, CEACAM5 taken as a single predictor achieves only significant performances in 0-1 YTD samples, but is among the top covariates in terms of importance across base-learners.

MUC16/CA125 also ranked high in importance across the base-learner classifiers and time-groups despite not performing well as a single predictor. MUC16 is a cell surface glycoprotein which can be elevated in tissue and sera of patients with various cancers and is mostly used in the diagnosis and prognostication of ovarian cancer ^46, 47^. MUC16 mediated metabolic reprogramming in pancreatic cancer is associated with tumour progression and metastasis through stromal remodelling ^47, 48^. Due to its overexpression on the surface of pancreatic cancer and absence from normal tissue, the value of MUC16 as a biomarker of PDAC has been investigated ^48^. A progressive change in expression of MUC16 throughout different stages of disease progression is already evident at pre-malignant (pancreatic intra-epithelial neoplasia; PaNIN) stages, highlighting its potential value as a diagnostic marker of early cancer ^46, 49^. With respect to its diagnostic performance, one meta-analysis which included 1235 patients reported a pooled sensitivity of 0.59 (95% CI 0.54-0.62; at 0.78 specificity, 95% CI 0.75-0.82) for detecting PDAC using CA125 (an epitope of MUC16) ^50^ as a single marker ^51^. When combined with CA19-9 (AUC 0.85) CA125 increased the overall accuracy of the former (AUC 0.89), demonstrating superior diagnostic accuracy compared to CA125 (or CA19-9) alone ^51^. The clinical value of MUC16 in prediction of metastasis and prognosis in PDAC has been also previously reported ^52, 53^. Serum levels of MUC16/CA125 have been shown to be the strongest predictor of metastatic disease (AUC of 0.892 at 95% CI 0.846 - 0.938, *p*<0.001) and survival (HR: 1.804, 95% CI 1.22 - 2.66, *p*=0.003) compared to other markers (including CEA) in 180 PDAC patients ^53^.

THBS2 and THBS1 were both significantly associated with PDAC, albeit not across all time-groups, the latter only in a subset of samples (see Supplementary Information). Thrombospondins are glycoproteins which mediate cancer growth and progression through cell-cell and cell-matrix interactions, tissue remodelling and regulation of inflammation, immunity and angiogenesis ^54, 55^. A specific role for THBS2 in tumour associated vascularisation has been reported in various cancers (including colon, liver, lung and melanoma) in which its aberrant expression was reported to be of both diagnostic and prognostic value ^54, 56^. With respect to PDAC, recent observations in a cohort of 493 (263 with PDAC) showed that THBS2 serum levels significantly differed and differentiated PDAC from high-risk individuals (familial pancreatic cancer patients) with a 55.9% test sensitivity (at 100% specificity; 100% PPV and 66.5% NPV). Moreover, PDAC patients with higher serum THBS2 (also termed TSP-2) levels showed worse clinical outcomes (hazard ratio = 1.54, 95% CI 1.143 – 2.086, *P* = 0.005). Interestingly, when combined with CA19-9 an improved panel sensitivity for PDAC (90.5% at 98.7 % specificity) was reported^54^.Taken as part of a signature as was presented here, THBS2 is expected to help robustly with early detection of PDAC.

In this study, among all interrogated clinical covariates, diabetes was the strongest predictor of a higher risk for PDAC (OR=13.26, 95% CI 1.36 - 1781.23, *P* =0.022), in our 3-4+ YTD cohort. An association between long-standing type 2 diabetes and a 1 to 1.5 fold increased risk of PDAC compared to non-diabetics has been reported ^57, 58^. The value of diabetes as a clinical covariate in a multi-marker panel is supported by the fact that the presence of new onset diabetes (NOD) (less than 3 years prior to diagnosis of PDAC) increases this risk 5-fold and is induced by various pancreatic conditions including chronic pancreatitis and PDAC-induced hyperglycaemia (recently described as type 3c diabetes). Abnormal fasting glucose or glucose intolerance are observed in the majority (>80%) of PDAC patients ^3, 58, 59^; PDAC as an underlying cause of NOD, is found in around 1% of individuals aged over 50 and therefore NOD might be considered an early warning sign of a pancreatic malignancy ^59, 60^.

Despite numerous reports of recent biomarker discoveries, lack of standardisation in sample identification and handling, methodologies of analysis as well as data capture and bioanalytical interpretation challenge their later validation in larger cohorts ^61^. Moreover, due to the biological complexity in PDAC, tested biological fluids and the overlapping features with high risk conditions (e.g. chronic pancreatitis), the predictive capabilities of single analytes are clearly insufficient to meet acceptable diagnostic performances^62^. Considering their individual, relative poor performance (79-81% test sensitivity and 82-90% specificity in the case of CA19-9 for example) as well as the low prevalence of PDAC in the general population however, accepting low values for test sensitivity and specificities results in a significant number of false positives. The combination of multiple analytes in diagnostic panels clearly enhances CA19-9 performance, as well as being able to compensate for cases in which CA19-9 detection is limited (Lewis body negative patients) ^63^. A mere combination of multiple biomarkers with low sensitivities at high specificities based on individual levels however, would be insufficient, and their independent contribution to the overall risk should be considered ^64^. With this in mind, the aim of the ensemble modelling strategy explored here was to improve on single classifiers by combining diverse techniques in a way such that the predictive performance of the ensemble would be greater and more robust when resorting to multi-marker models, thus addressing the issues highlighted above. The pool of chosen classifiers proved to be useful in highlighting different aspects of the data by selecting a larger panel of biomarkers and allocating different importance to each. Despite finding that the stacking protocols explored offered substantial and statistically significant improvements over the previous state-of-the-art prediction methods, we expect that scanning the space of all classifier combinations from available code libraries will improve the results. Yet, we must resort to other methods for finding better performing ensembles when predicting PDAC. Early detection is a notoriously difficult problem due to the extreme class imbalance, missing values and the necessity to create predictive indices performing well at representative disease prevalence. Because the robustness of ensembles is an emergent, not an explicit property, different directions for future work should be taken by formalizing the effects of calibration on heterogeneous ensemble performance and explicitly incorporating diversity in the search ^18, 65^. In fact, this will be even more prominent when combining longitudinal methods ^66^ with ensemble selection and stacking techniques. To our knowledge longitudinal samples and associated time-series classification techniques have never been applied to early detection of pancreatic cancer. Dynamic changes of CA19-9 and MUC16 identified in previous work ^12^ are an indication of the importance of these types of studies. The importance of trends and rates of change in each biomarker taken separately or in multi-marker models has also proven to be advantageous in improving early detection in ovarian cancer longitudinal models ^66^ and mechanistic tumour and biomarker secretion models^67^. Further developments in PDAC longitudinal data sets and ensemble model selection routines should highlight the importance of early and late biomarker panel dynamic changes and increase performance in a clinical setting, in addition to contributing with invaluable methodologies to the field of machine learning and artificial intelligence in early detection of cancer ^68^.

## Materials and Methods

### Study Design

This nested case control discovery study was approved by the Joint UCL/UCLH Research Ethics Committee A (Ref. 05/Q0505/57). Written informed consent was obtained from donors and no data allowing identification of patients was provided. The study set comprised serum from post-menopausal women aged 50-74 recruited to UKCTOCS between 2001 and 2005 and collected according to an SOP ^24, 25^. All participants were ‘flagged’ with the national agencies for cancer registrations and deaths using their NHS number. Women subsequently diagnosed with pancreatic ductal adenocarcinoma (cases) were identified by cross-referencing with the Health and Social Care Information Centre cancer registry codes and death codes (ICD10 C25.0/1/2/3/9). Confirmation of diagnosis was sought from GPs and consultants through questionnaire and from the Hospitals Episode Statistics database. In total, 143 cases were identified (with 219 associated serum samples) that had not been registered as having any other cancer since randomization and that had a confirmed diagnosis of pancreatic cancer. Matched non-cancer controls, i.e., with no cancer registry code, from individual women were selected based on collection date and centre to minimize variation due to handling and storage. From this set, 248 controls were selected. 35 of the PDAC cases had longitudinal data, with between 2 and 6 annual longitudinal samples per individual years before diagnosis. Due to the design of the UKCTOCS study, PDAC stage for all the cases at the time of diagnosis was not available.

### Serum Analyte Measurements

All serum samples were randomized for testing. A total of 111 analytes were tested. CA19-9(A) was measured using the Mucin PC/CA19-9 ELISA Kit (Alpha Diagnostic International) according to the manufacturer, using a 1:4 serum dilution. CA125(A) (MUC16) assay was performed using the Cobas CA125 II CLIA with a CA125 II Calibrator Set (Roche and Fujirebio Diagnostics) on a Cobas E411 analyzer with PreciControl Tumour Marker to monitor assay imprecision. LRG1 level was assessed using the human LRG1 ELISA Assay Kit (Immuno-Biological Laboratories) at a 1:2000 serum dilution. PIGR was measured using the human secretory component (SC) ELISA Kit (Cusabio) at a 1:500 serum dilution. REG3A/PAP level was determined using the PANCREPAP ELISA Kit (DynaBio) at a 1:100 serum dilution and Factor XII (F12) using the Factor XII Human ELISA Kit (abcam) at a 1:1000 serum dilution. For vWF, we resorted to the Von Willebrand Factor Human ELISA Kit (abcam) at a 1:100 serum dilution. THBS1 (TSP1) level was evaluated using the Quantikine Human Thrombospondin-1 Immunoassay (R&D Systems) at a 1:100 serum dilution. AGR2 was calculated using the Anterior Gradient Protein 2 ELISA kit (USCN Life Science) at a 1:25 serum dilution. A1AT (SERPINA1) was measured by α-1-Antitrypsin ELISA kit (Immunodiagnostik AG) and IL6ST (IL6RB) by Quantikine human soluble gp130 (R&D Systems). THBS2 (TSP2) was measured using the Quantikine Human Thrombospondin-2 Immunoassay (R&D Systems) at a 1:10 serum dilution and TEK using the Quantikine human TIE-2 ELISA Assay Kit (R&D Systems) at a 1:10 serum dilution and IGFBP1 by human IGFBP1 ELISA Assay Kit (abcam) at a 1:50 serum dilution. Finally, IL17RA was measured using the human IL17RA ELISA Assay Kit (Abnova). Assays were performed on singlet test samples and values not measurable on the standard curves were given the value ‘low’.

The group of markers AFP, TEK and IGFBP1 was only measured in a subset of subjects from the total used to develop the classifiers presented in the main text. A similar situation occurred for the group of markers LRG1, PIGR, REG3A, F12, AGR2, IL17RA, SERPINA1 and THBS1. Some of these biomarkers were motivated by a previous paper ^12^. See Supplementary Information for further details and results.

Samples were also tested using Olink’s multiplex immunoassay Oncology II panel. Known cancer antigens, growth factors, receptors, angiogenic factors and adhesion regulators were measured (see Table S1).

### Statistical Analysis

We divided the whole set of samples into a training (2/3) and validation (1/3) set, by stratifying for age quartile, HRT use at randomization, OCP use (ever), Diabetes status, BMI quartile, PDAC and control status and sample single time-group, i.e., 0-1,1-2,2-3,3-4 and 4+ YTD, attributed to each sample determined by the time from collection to diagnosis. Despite the fact that 35 of the PDAC cases had longitudinal data, with between 2 and 6 annual longitudinal samples per individual years before diagnosis, with an average of 1.53 samples per individual (see Table 1), all samples were taken as independent, and no intra-individual correlation was imposed or explicitly modelled during data analysis in this instance.

Receiver operating characteristic (ROC) curves were constructed for each model to assess diagnostic accuracy. The AUC for the ROC curves was used as the performance metric. Models were selected based on their rank in the training set across cross-validation folds. ROC curves were generated with the *pROC* R package (version 1.15.3). 95% CI for AUCs were determined by stratified bootstrapping. All AUC confidence intervals crossing 0.5 were deemed insignificant. In addition, sensitivity, positive and negative predictive values at 90% specificity are also reported.

In order to evaluate the association between each of the single markers, including the clinical covariates (see Table 1), and PDAC status, we resorted to the logistic regression model implemented in the *logistf* R package (https://cran.r-project.org/web/packages/logistf/logistf.pdf, version 1.23). This approach fits a logistic regression model using Firth’s bias reduction method. The reported confidence intervals for odds ratios and tests were based on the profile penalized log likelihood and incorporate the ability to perform tests where contingency tables are asymmetric or contain zeros. P values were used to rank markers. The performance of single marker models was also verified in the test set (Fig. S3 to S14 and Supplementary Information section on single marker performance).

Multi-dimensional analysis of the data was performed under two separate frameworks: a brute-force algorithm scanning through combinations of up to 3 markers and fitting a logistic regression model (see Supplementary Information), and a stacked ensemble algorithm with 10 base-learners. The models stemming from the brute-force approach were ranked according to their performance across re-sampling training folds. The ensemble models relied on the performance of the base-learners presented in Fig. 1 and highlighted below:

- Decision trees and rule-based models for pattern recognition (C50, https://cran.r-project.org/web/packages/C50/C50.pdf);
- Support vector machines with radial basis function kernel (SVM, https://cran.r-project.org/web/packages/kernlab/kernlab.pdf);
- Regularized random forests (RRF, https://cran.r-project.org/web/packages/RRF/RRF.pdf);
- Neural networks with feature extraction (NNET, https://cran.r-project.org/web/packages/nnet/nnet.pdf);
- Gaussian process with radial basis function kernel (GAUSSPR, https://cran.r-project.org/web/packages/kernlab/kernlab.pdf)
- Lasso and elastic-net regularized generalized linear models (GLMNET, https://cran.r-project.org/web/packages/glmnet/glmnet.pdf)
- Bagged Adaptive Boosting (ADABAG, https://cran.r-project.org/web/packages/adabag/adabag.pdf)
- Extreme gradient boosting (XGBOOST, https://cran.r-project.org/web/packages/xgboost/xgboost.pdf)
- Generalized Linear Model with Stepwise Feature Selection with Akaike Information criterion (GLMStepAIC, https://cran.r-project.org/web/packages/MASS/MASS.pdf)
- Naïve Bayes classifier (NB, https://cran.r-project.org/web/packages/klaR/klaR.pdf)

The selection of base-learners was grounded on covering a number of state-of-art methods and algorithmic families, from bagging and boosting to general linear models with in-built feature extraction, previously referenced in the literature ^16, 19^, that would be able to capture different aspects of the data with an efficient computational effort and that had, for the most part, typical hyperparameter ranges published in the literature ^19^, some with applications in biology ^18^. Due to the size of the data sets, we narrowed down the size of set of base-learners to 10. Further work on ensemble selection from libraries of models should contribute to clarifying if other techniques add significant value ^16^ by testing performance against base-learner pool diversity ^18^. The training of the base-learners was executed in two ways:

- by taking joined/combined time-group samples, i.e., collected 0-1, 0-2, 0-3, 0-4, 0-4+ YTD or
- by training the set of base-learners in each single time-group specific samples, i.e., 0-1, 1-2, 2-3, 3-4, 4+ YTD.

The first model forces the base-learners to learn specific and cross-time-group details together, whereas the second model creates specialized groups of base-learners per single time-group. We tested several staking procedures: by Bayesian Model Averaging (https://cran.r-project.org/web/packages/BMA/BMA.pdf) with an underlying logistic regression model (BMA stack), by averaging with an arithmetic mean (MEAN stack) and geometric mean across the probabilities attributed by each base-learner (GEOMEAN stack), or by taking the maximum probability across all base-learners (MAX stack). This class is named throughout this paper as JTG2L (see Fig. 1 and Figs. S19). For the second model we tested a 2-layer and a 3-layer stacked model. The first, referred to as STG2L (Fig. S20), took the base learners trained in each single time-group and applied the 4 stacks as mentioned above, although to a larger stack input space. If, for example, we are training with samples belonging to every single time-group, i.e., 0-1, 1-2, 2-3, 3-4, 4+, the stack feature input space will have 10 times 5 dimensions; each base-learner is trained on each single time-group, giving 5 models per base-learner and a total of 50 base-models (Fig. S20). Subsequently, the probability output from each base-learner model is concatenated and fed into the meta-learner. For the specific case of the STG2L protocol, we also tested an average neural network meta-learner model (AVNNET stack) trained on the concatenated probability matrix created from each base-learner probability output. The second, named STG3L (Fig. S21), stacks twice and, therefore, has 3 layers. First it stacks the base-learners per single time-group with a BMA stack and, subsequently, stacks the result, a 5-dimension feature space of probabilities with either a BMA stack, a MEAN stack, a GEOMEAN stack or a MAX stack, if, for example, we are training with samples belonging to every single time-group (see Fig. S21 for further details). This approach is reminiscent of stacking auto machine-learning procedures implemented in *AutoGluon* (https://github.com/awslabs/autogluon). Other combinations of time-groups were also tested, e.g., 0-1 plus 1-4, 0-2 plus 2-4, etc., but the stacked classifiers either underperformed or were not robust.

All base-models were trained by 5 times repeated 10-fold cross-validation with over-sampling of the minority class, in our dataset the PDAC cases (see Table 1). In order to avoid overfitting, we ranked each of the features, both biomarkers and clinical covariates, with the logistic regression model mentioned before, and scanned the ranked feature input space, in increments of 10 features, with the objective of finding the optimal performance across cross-validation folds, without bias ^69^. Despite some features not being significant according to the logistic regression model with bias correction when evaluated as a single predictor, the protocol we applied scanned over all 101 features, clinical covariates included. As described, the stacked models were trained on the probability matrices, i.e., generated by concatenating the vectors whose entries are the probability of being a case according to each base-learner. We opted for a 10-times 10-fold cross-validation resampling strategy for the meta-learner to further secure that the choice of models was robust. The extensive resampling strategy secured both at the base-learner level as well as the meta-learner level that the models learn and performed robustly across a large number of diverse folds, thus further ensuring that the longitudinal samples taken as being independent did not skew the training and reduce the generalization power of the PDAC classifiers. We also developed other classifiers trained in both real and synthetic data by applying state of the art techniques such as SMOTE ^70^ and non-parametric algorithms ^71^ (see Supplementary Information for details).

The fact that the PDAC cases had longitudinal samples which we considered as independent, did not affect the training of the models. The 5-times repeated 10-fold cross-validation resampling strategy secured that the random allocation of samples to training and validation folds during training avoids a systematic use of samples from the same individual in hyperparameter optimization. Regarding the stratification of training and test sets, given that this was done also with information of sample single time-groups, there wasn’t a consistent presence of samples from the same individual which would interfere with the performances reported here.

The variable importance routine selected for evaluating feature importance in each base-learner (see for example Fig. 2) was a model-agnostic method based on a simple feature importance ranking measure ^72^, implemented in the R package *vip* (https://cran.r-project.org/web/packages/vip/index.html). Model-agnostic interpretability separates interpretation from the model, is a more flexible approach and can be applied to any supervised learning algorithm. It was crucial in our case for understanding the variance in importance attributed by each base-learner since some classifiers have in-built routines, e.g., Random Forests (RRF, Fig. 2), and others don’t, e.g., Naïve Bayes (NB, Fig. 2).

The enrichment analysis for each of the signatures developed with single and joined time-groups was performed with the *gprofiler2* R package (https://cran.r-project.org/web/packages/gprofiler2/gprofiler2.pdf) version 0.2.0. In Fig. 3 only up to 15 significant terms are shown. Threshold for multiple comparison correction under false discovery rate was set at 0.05.

## Data Availability

Data requestors will need to sign a data access agreement and in keeping with patient consent for secondary use, obtain ethical approval for any new analyses. All packages used in the pre-processing of data and subsequent analysis have been identified to secure full reproducibility.

## Supplementary Information

In this supplementary information we provide the additional tables and figures cited in the main text, as well an extensive analysis of single biomarker association with PDAC and previously reported multi-dimensional models, further results of ensemble classifiers specialized in single time-group samples, the effects of training the base-learners with real and synthetic data and, finally, PDAC signatures developed with additional proteomic data for a sub-group of subjects.

## Acknowledgments

We thank the participants of the UKCTOCS trial; the management team, research nurses, interviewers, research assistants, and other staff who gathered the data that was used in this study

## Author contributions

NRN, JT and AZ conceived the study. JT, AZ, SP and UM secured funding. NRN did model building and statistical analysis, produced the figures, interpreted the data in collaboration with JT and AZ. NRN, JT and AZ drafted the paper. HJ performed the experiments for thrombospondin-2. All authors contributed to data acquisition and interpretation, and critically reviewed and approved the article.

## Supplementary Information

### Single biomarker association with PDAC and previously reported multi-dimensional models

Levels of the established tumour markers CA19-9, MUC16 and CEACAM5 showed a significant association with PDAC in samples taken less than 1 YTD (Figs. S1 and S6), under a logistic regression model developed in the training set (see details in the Methods section), which is consistent with previous reports ^12^. Notably, THBS2 also appeared to have a predictive value in samples taken up to a year to diagnosis (OR=1.73 (95% CI 1.17-2.70), *P*= 0.0054, AUC=0.65 (95% CI 0.55-0.75)) (Figs. 1 and S4), between 1 to 2 years (OR=2.34 (95% CI 1.23 - 4.87), *P*= 0.0081, AUC=0.67 (95% CI 0.55-0.79)) (Figs. 1 and S5), and above 4 YTD (OR=2.63 (95% CI 1.17 - 6.84), *P*= 0.018, AUC=0.67 (95% CI 0.52-0.82)) (Figs. 1 and S8). A link between THBS2 and an increased risk for pancreatic cancer has been reported before ^55^ and was shown to improve the ability of CA19-9 to distinguish PDAC from pancreatitis, with a specificity of 98% and a sensitivity of 87% for PDAC. Samples in that report, however, were collected after diagnosis. The samples used in our work were all gathered prior to diagnosis, the latest approximately 1 month before. THBS2, nevertheless, did not generate significant AUCs in the test set, whether with logistic regression models trained with single time-groups (Fig. S1) or combined time-groups (see also Fig. S6 to S14). The only other marker that remained a consistent predictor in more than one single time-group was CA19-9: OR^training^_(0-1)_=1.67 (*P*=2.92 x 10^-9^), AUC^training^_(0-1)_=0.81 (95% CI 0.71 - 0.89) and Sens^training^_(0-1)_= 0.64 (95% CI 0.48 - 0.77) at 90% Spec; OR^training^_(1-2)_=1.67 (*P*=2.92 x 10^-9^), AUC^training^_(1-2)_=0.66 (95% CI 0.53-0.78), Sens^training^_(1- 2)_= 0.31 (95% CI 0.10 - 0.49) at 90% Spec; OR^training^_(2-3)_=1.60 (*P*=0.0017), AUC^training^_(2-3)_=0.71 (95% CI 0.59-0.83), Sens^training^_(2-3)_= 0.33 (95% CI 0.061 - 0.61) at 90% Spec (Fig. S1). When attempting to predict PDAC status in the test set with the logistic regression models trained with time-group 0-1 samples and used to rank biomarkers, we still verified that CA19-9 was one of the two biomarker models, the other being CEACAM5 (OR^training^_(0-1)_=1.94 (95% CI 1.32 - 3.08), *P*= 0.00029 and AUC^training^_(0-1)_= 0.67 (95% CI 0.56 - 0.77) in the training set and AUC^test^_(0-1)_=0.84 (95% CI 0.65 −0.97) in the test set) (see Fig. S1 and Figs. S6 to S14), that achieved significant performances: AUC^test^_(0-1)_=0.73 (95% CI 0.52 - 0.93), Sens^test^_(0-1)_= 0.62 (95% CI 0.38 - 0.85) at 90% Spec. The combination of CA19-9 and THBS2 improved the performance in both training and the test sets, for 0-1 samples, i.e., AUC^training^_(0-1)_ (CA19-9, THBS2) = 0.82 (95% CI 0.73 - 0.89) and AUC^test^_(0-1)_ (CA19-9, THBS2) = 0.74 (95% CI 0.51 - 0.92), although the difference with respect to the CA19-9 single marker model was not significant, i.e., *P*=0.19 and *P*= 0.69, respectively, under a DeLong method for comparing AUCs from 2 different ROC curves (see Methods). Additionally, the combined 2-marker model CA19-9+CEACAM5 did not improve the diagnostic performance of CA19-9 alone in the training set (AUC^training^_(0-1)_=0.80 (95% CI 0.71-0.88)), but improved in the test set, i.e., AUC^test^_(0-1)_=0.81 (95% CI 0.62 - 0.95), although not sufficiently for the difference to be significant (*P*=0.24). MUC16 (commonly known as CA125), which was previously reported to provide additional sensitivity when combined with CA19-9 in a pre-diagnosis setting similar to ours ^12^, was ranked as significant in our study, with an OR^training^_(0-1)_=2.72 (95% CI 1.66 - 5.01), *P*= 8.7 x 10^-6^, AUC^training^_(0-1)_= 0.72 (95% CI 0.62 - 0.82) and Sens^training^ _(0-1)_= 0.64 (95% CI 0.29 - 0.67) at 90% Spec, but was borderline insignificant when tested against blinded samples, i.e., AUC^test^_(0-1)_=0.72 (95% CI 0.49 - 0.90). As was the case of the combined model with THBS2, the bi-dimensional model CA19-9+MUC16 had a similar performance to the CA19-9, i.e., AUC^training^_(0-1)_ (CA19-9, MUC16) = 0.81 (95% CI 0.72 - 0.88) and Sens^training^_(0-1)_= 0.58 (95% CI 0.46 - 0.75) at 90% Spec, and therefore with no significant improvement in performance (*P*= 0.91). When evaluated in the test set, the CA19-9 and MUC16 multi-marker model outperformed CA19-9 and the combination of CA19-9 and THBS2, i.e., AUC^test^_(0-1)_ (CA19-9, MUC16) = 0.76 (95% CI 0.55 - 0.93), but not significantly, *P*=0.76 and *P*=0.80, respectively. The CA19-9+CEACAM5 model was, therefore, the best multi-marker predictor in the test set built from frequently cited markers but once again the difference was not significant when compared to CA19-9 alone (*P*=0.44). We must stress that any of the bi-dimensional models reported above did not involve any interaction terms because if we resorted to non-linear terms in the training set the results were not significant for any combinations of the typical panel of analytes reported in the literature.

Several other multi-marker combinations have been put forward to add value to traditionally used panels ^73^ (see Fig. S1 for the typical single biomarkers used). In fact, as was observed in the training set 0-4+ samples, a large group of markers appeared significant: CA19-9, VWF, THBS2, MUC16, HGF, CEACAM5, VEGFA, FURIN, CTSV, NT5E, TLR3 (see Fig. S14). These findings further stimulated the exploration of multi-dimensional models in order to reach higher performances and robustness across all time-groups. If we resorted once again to the simple logistic regression approach and generated models with up to 3 features, selected from those reported in the literature ^3, 73, 74^, e.g., CA19-9, CA125, CEACAM5, HGF, THBS2, CEACAM1 (Fig. S1), included clinical covariates and chose the top 10 according to their performance across training cross-validation folds, we concluded that despite generating a significant performance in the test set, a clear advantage in building models from this restricted pool of features was lacking, especially when the sensitivity at 90% specificity values were relatively low (see Fig. S2 and, additionally, Fig. S15 for results obtained with single time-groups). What was notable about these 3-marker models was the presence of the covariate DIABETES and AGE in top ranks, as we had predicted (see Main Text). DIABETES was a predictor of PDAC in samples allocated to all single time-groups in the training set, with exception of 1-2 YTD (see Fig. S1). There was, therefore, the need for a systematic search over a larger panel, in our work composed of in-house and Olink biomarkers (see Table S1), and for the use of several different classifier types which we combined with an ensemble modelling technique (see Methods and Results sections in the Main Text).

### Ensemble classifiers specialized in single time-group samples

We also developed classifiers specialized in each single time-group, i.e. 0-1, 1-2, 2-3, 3-4 and 4+, here referred to as STG2L and STG3L (see Methods for details and Figs. S20 and S21), by resorting to the same framework, i.e. stacked ensemble with repeated cross-validation and identification of optimal size of input space, and combining the probability/risk of being a case of each sample generated by each specialized model either by taking only the output that had the maximum probability, by averaging, i.e. with a Bayesian model or a simple arithmetic average, or utilizing a mixture of experts, in our case, by calculating the geometric probability mean across all models. The latter is more conservative since a subject is only classified as a Case if there is a central tendency in the 10-model probability vector of outputs. Both the maximum probability principle as well as the mixture of experts underachieved significantly in the test set when compared with the standard approach (see Figs. S22 and S23). The problem with training specialized ensembles per single time-group lies with the sample size. Their performance in the training set indicates that these alternative models, despite the extensive re-sampling strategy are not robust both at the base-learner level (Fig. S16) and the meta-learner level (Figs. S22 and S23). Consequently, their potential as predictive indices is reduced systematically, despite the overall pair-wise diversity among base-learner predictions being high in the test set (Fig. S18, see also Fig. S27 for feature importance). Further expansion of the dataset should clarify if the use of single time-group specialized classifiers is advantageous.

### Training the base-learners with real and synthetic data

For the purposes of exploring the effects of re-balancing the imbalanced classes, in our work PDAC and Control, across all time-groups, we also developed other classifiers under the JTG2L class (see Methods for details and Figs. S19 and S22) but trained in both real and synthetic data we generated by fitting a high-dimensional Dirichlet Process Gaussian Mixture (HD-DPM) Model ^71^ model to the cases of the training set and, independently, to the controls. The high-dimensionality character of the HD-DPM method is required to capture cross-biomarker correlations, which would not have been preserved had we fitted a one-dimensional DPM model to each biomarker independently. For the sake of completeness, we also tested the Synthetic Minority Oversampling Technique (SMOTE) ^70^. The inclusion of the routines for synthetic data generation was done during re-sampling and prior to the application of each stacked algorithm. This is particularly important for the HD-DPM algorithm since a separate distribution is fitted to PDAC cases and controls. The number of synthetic samples needs to be similar to those of real samples, otherwise the classifiers learn the distribution of the synthetic data instead.

This did not improve the performance either in the training set or the test sets (see Figs. S29 and S30) and thus renders the option of synthesizing data and enlarging the number of representative examples to aid the training process not viable. Both techniques were only run for joined time-groups, as the number of samples in each single time-group was not sufficient to implement either the HD-DPM or the SMOTE algorithm. Moreover, there is evidence that for high-dimensional problems techniques such as over-sampling/under-sampling of the minority/majority class outperforms methodologies such as SMOTE ^70^. We also tested the ROSE algorithm ^75^ for synthesizing data during cross-validation resampling steps, a technique similar to HD-DPM despite not providing the benefits of non-parametric modelling. Overall, the results obtained with ROSE were far inferior to any of the other 3 algorithms, and as is the case for synthetizing samples with HD-DPM, a lengthy process. Therefore, we ran the remaining models with the oversampling of the minority class as a strategy to develop all the ensembles of base-learners reported in the Main Text.

### PDAC signatures developed with additional proteomic data for a sub-group of subjects

In addition to the data set used to generate the models reported in the Main Text, we also had additional biomarkers that had been measured only in sub-groups of samples. Sub-set I includes the biomarkers AFP, TEK and IGFBP1 (see Figs S31 and Tables S5, S6 and S7), while sub-set II includes the additional biomarkers LRG1, PIGR, REG3A, F12, AGR2, IL17RA, SERPINA1 and THBS1 (see Figs. 34 and Tables S8, S9 and S10). These were biomarkers motivated by a previous paper ^12^. The BMA ensemble stack developed in sub-set I of the training set showed inferior performances compared to the original model (Figs. S32 and S33), whereas the stack developed in the sub-set II improved the predictive potential in sub-set II of the test set, with performances as high as 0.94 (95% CI 0.75-1.00) and 0.88 (95% CI 0.71-0.99), in 0-1 and 0-2 samples, respectively (Figs. S35 and S36). Also noteworthy is the importance attributed to each new feature across base-learners (Fig. S37). For the models developed in sub-set II, 4 of the new biomarkers are ranked in the top 20 markers by feature importance across joined time-groups: IL17RA; AGR2; SERPINA and LRG1. REG3A ^12^ and THBS1^73^ have been studied before in association with PDAC. REG3A was found to be a late marker adding little to combined models with CA19-9 and MUC16 ^12^. Despite this, it is attributed an importance different from zero across the ensemble classifier developed here (Fig. S37).

## Supplementary Figures

**Fig. S1.**
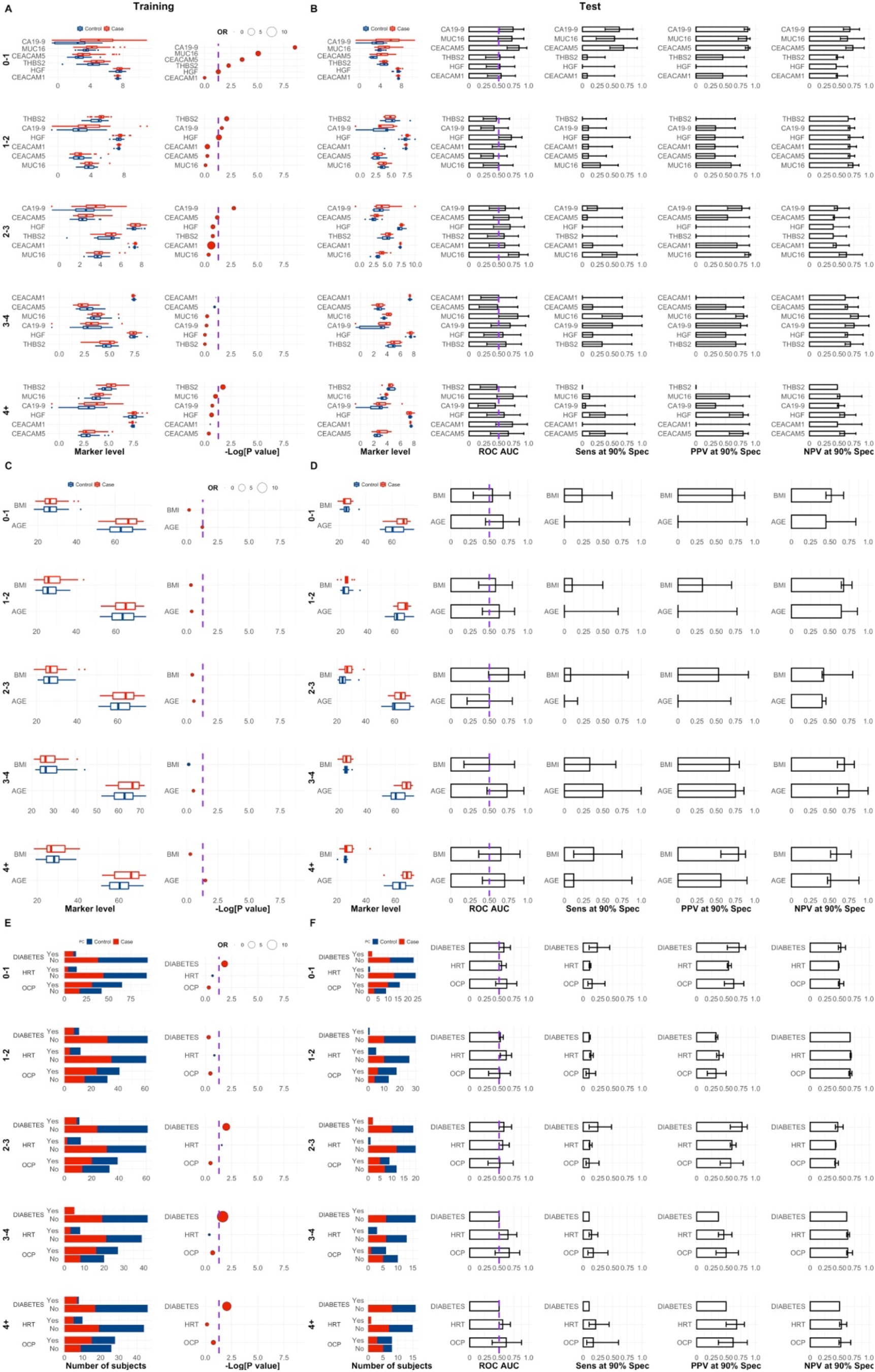
Distributions for state-of-the-art markers and clinical covariates in the training and test sets, per single time-groups. **A** Marker distributions and p-values for CA19-9, MUC16, CEACAM5, THBS2, HGF and CEACAM1, in the training set. P values are calculated according to a logistic regression model with a bias reduction method (see Methods). **B** Marker distribution, ROC AUC, sensitivity, positive predictive value and negative predictive at 90% specificity for the same biomarkers, but in the test set. AUCs were determined with the respective unidimensional marker model developed in the training set. **C** Marker values and respective p-values for AGE and BMI, in the training set. **D** Similar to B but for AGE and BMI. **E** and **F** Same as above but for Diabetes, HRT and OCP. Purple dashed lines in A, C and E represent the significance threshold associated with a p-value of 0.05. In B, D, F the ROC AUC significance threshold is also represented by a purple dashed line at 0.5. OR stands for odds-ratio. Red and blue OR points represent OR > 1 (favours PDAC status) and OR < 1 (favours Control status), respectively.

**Fig. S2.**
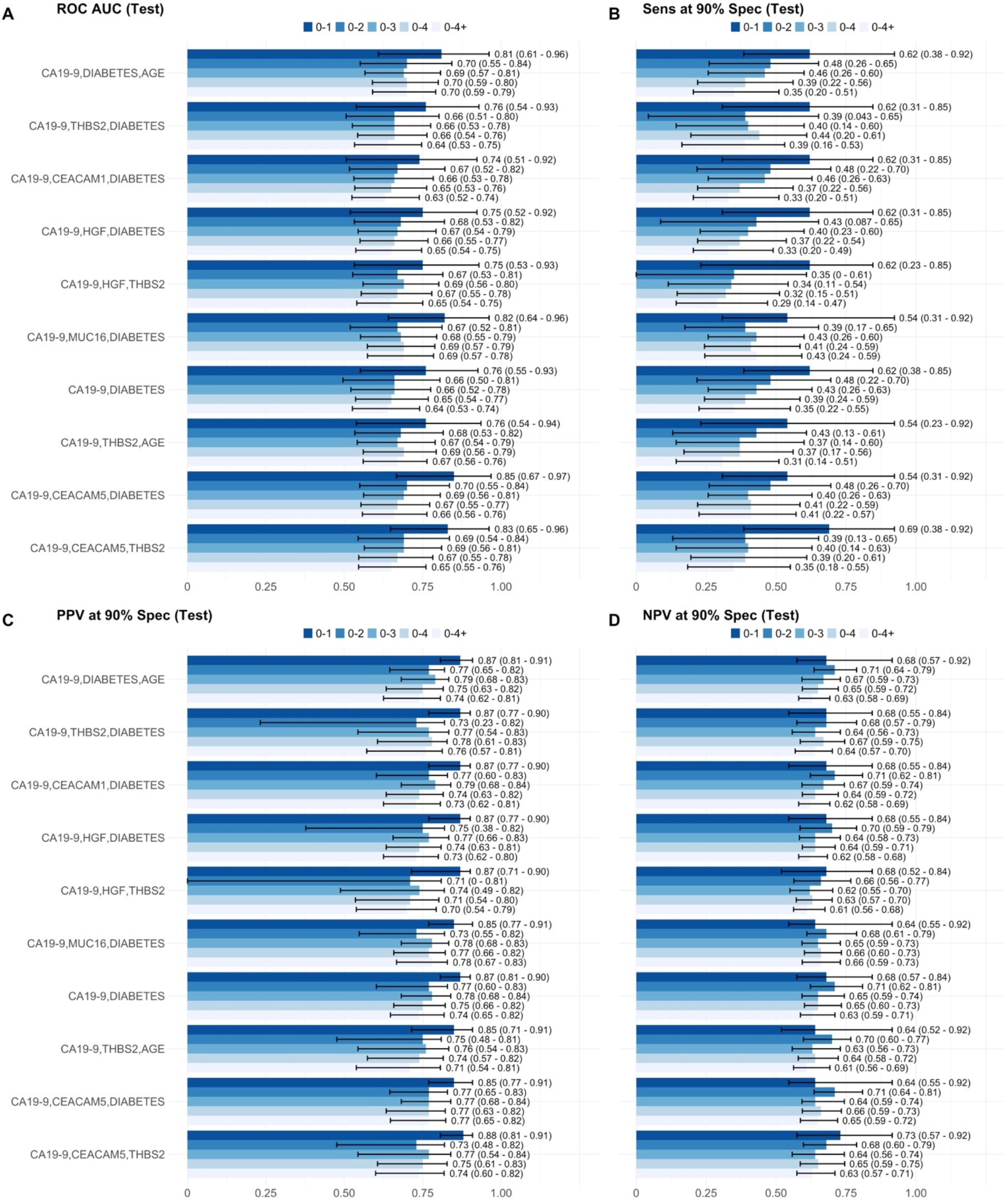
Performances for state-of-the-art multi-marker and clinical covariate combinations in a logistic regression model, top models for joined time-groups. ROC AUC (Test) values were calculated with the combinations of features developed in the training set. Models are ranked according to their performance in the training set with a 5 times 10-fold cross-validation resampling strategy. Error bars in figures correspond to 95% CI for AUCs, determined by stratified bootstrapping.

**Fig. S3.**
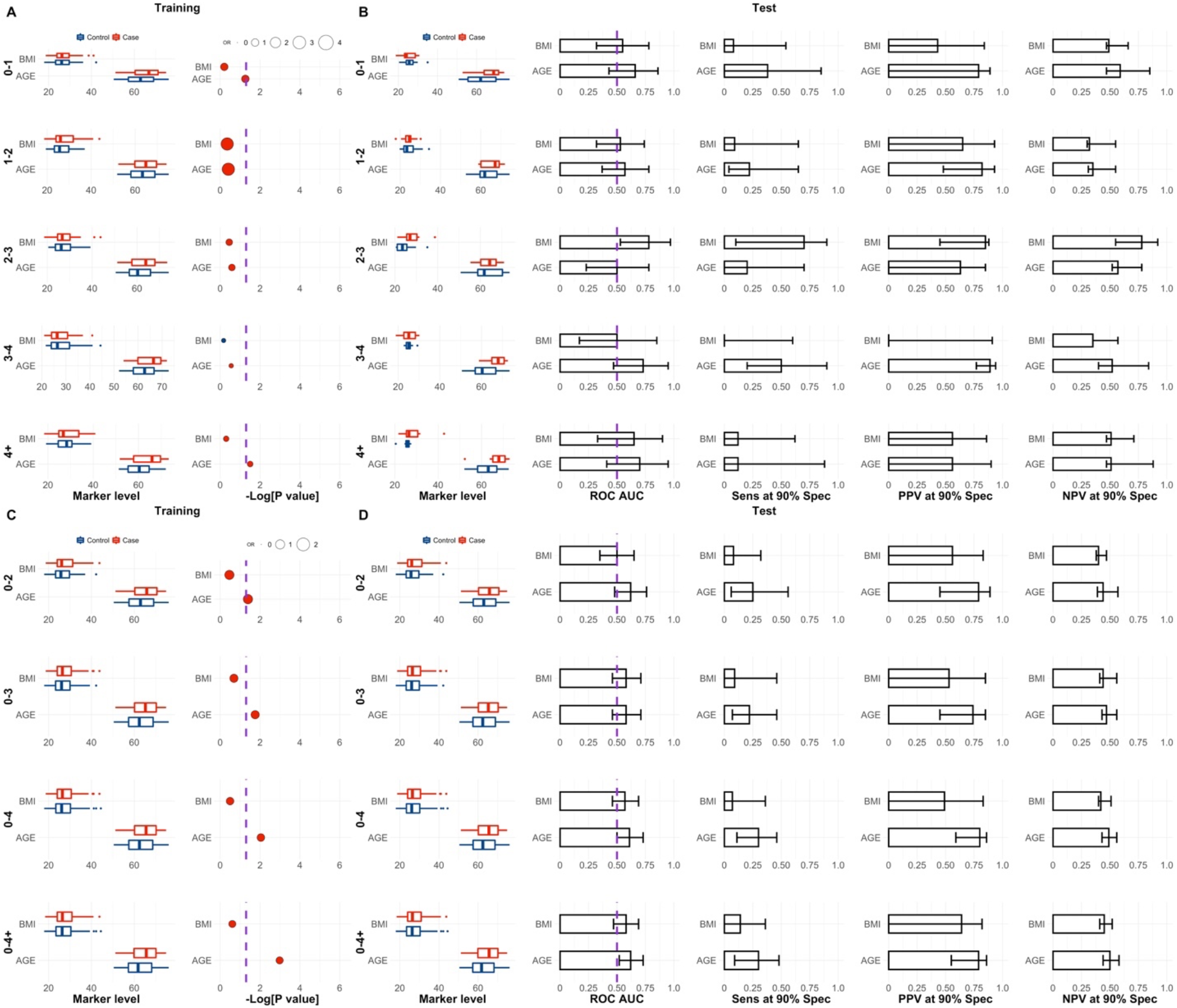
Characteristic distribution for BMI and AGE in the training and test sets. **A** and **B** Single time-groups. **C** and **D** Joined time-groups. P values are calculated according to a logistic regression model with a bias reduction method (see Methods). ROC AUC values were calculated with the single feature developed in the training set. Error bars in figures corresponding to the test set correspond to 95% CI for AUCs, calculated from 2000 bootstrapping samples.

**Fig. S4.**
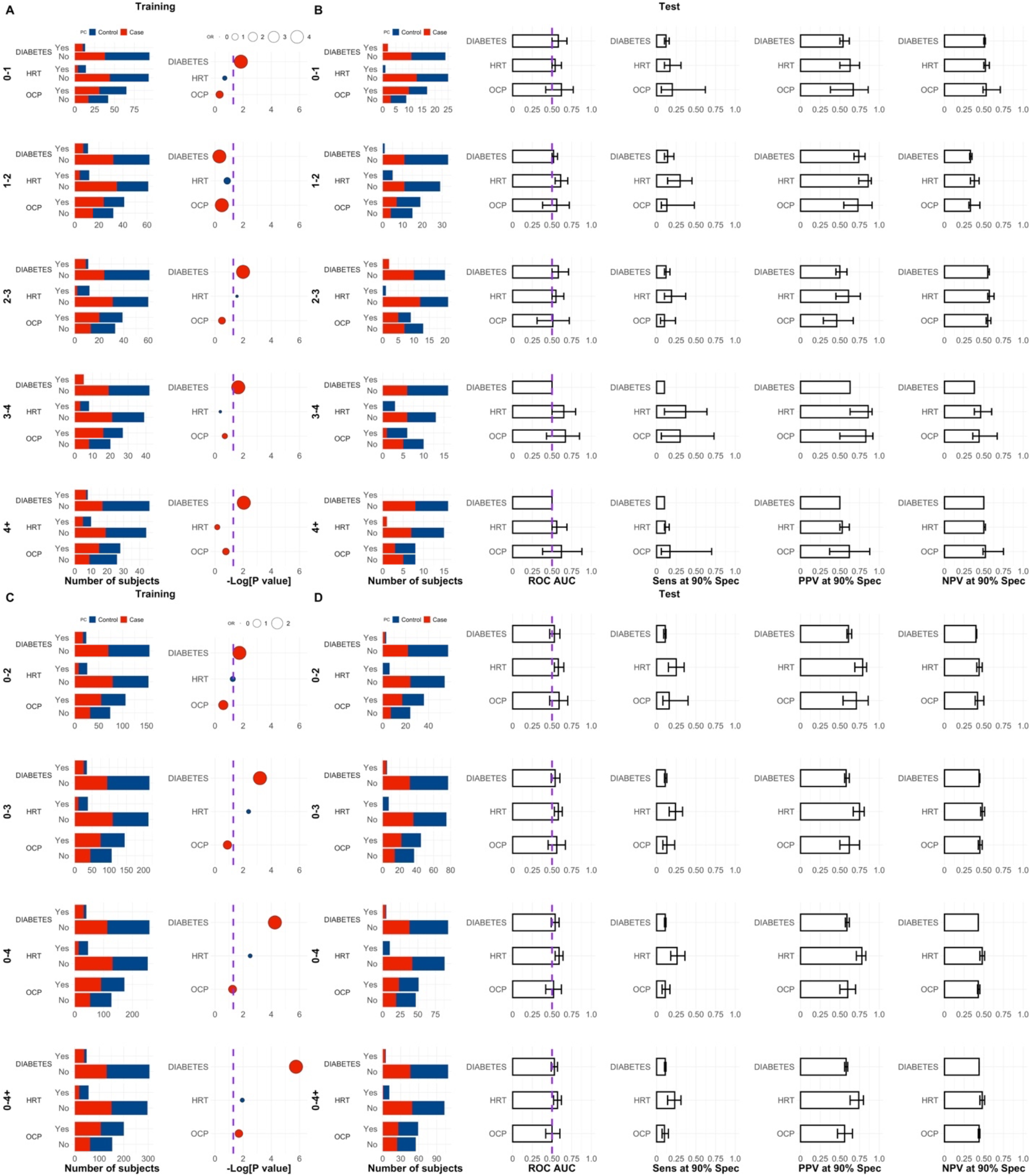
Characteristic proportions for DIABETES, HRT and OCP use in the training and test sets. **A** and **B** Single time-groups. **C** and **D** Joined time-groups. P values are calculated according to a logistic regression model with a bias reduction method (see Methods). ROC AUC values were calculated with the single feature developed in the training set. Error bars in figures corresponding to the test set correspond to 95% CI for AUCs, calculated from 2000 bootstrapping samples.

**Fig. S5.**
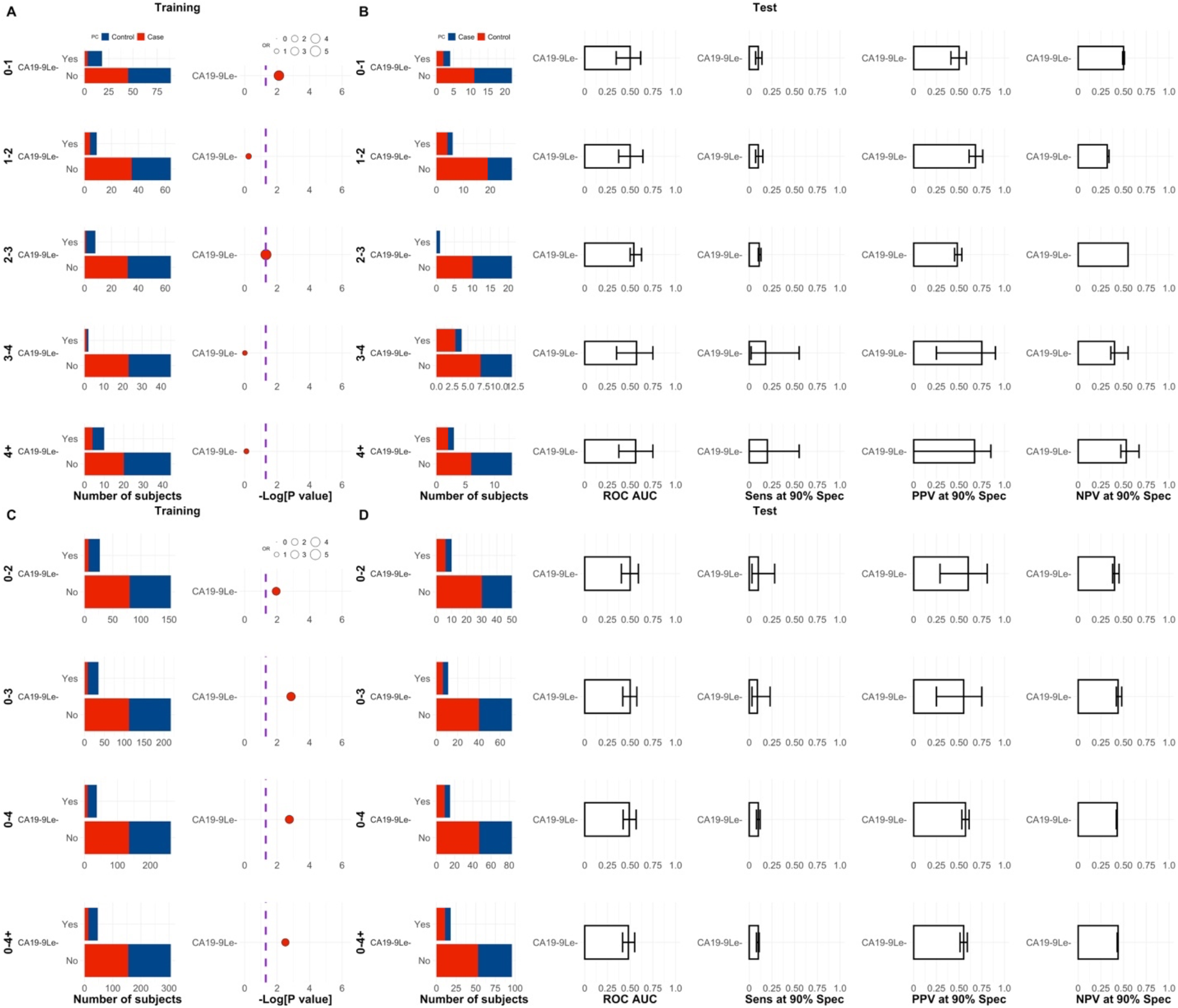
Number of subjects according to PDAC status and speculated CA19-9 Lewis negative status (Le-). **A** and **B** Single time-groups. **C** and **D** Joined time-groups. P values are calculated according to a logistic regression model with a bias reduction method (see Methods). ROC AUC values were calculated with the single feature developed in the training set. Error bars in test set figures correspond to 95% CI for AUCs, calculated with stratified bootstrapping. The Le-character associated with each subject was not determined by sequencing but by the absence of CA19-9 in the sample, which makes it a speculative feature

**Fig. S6.**
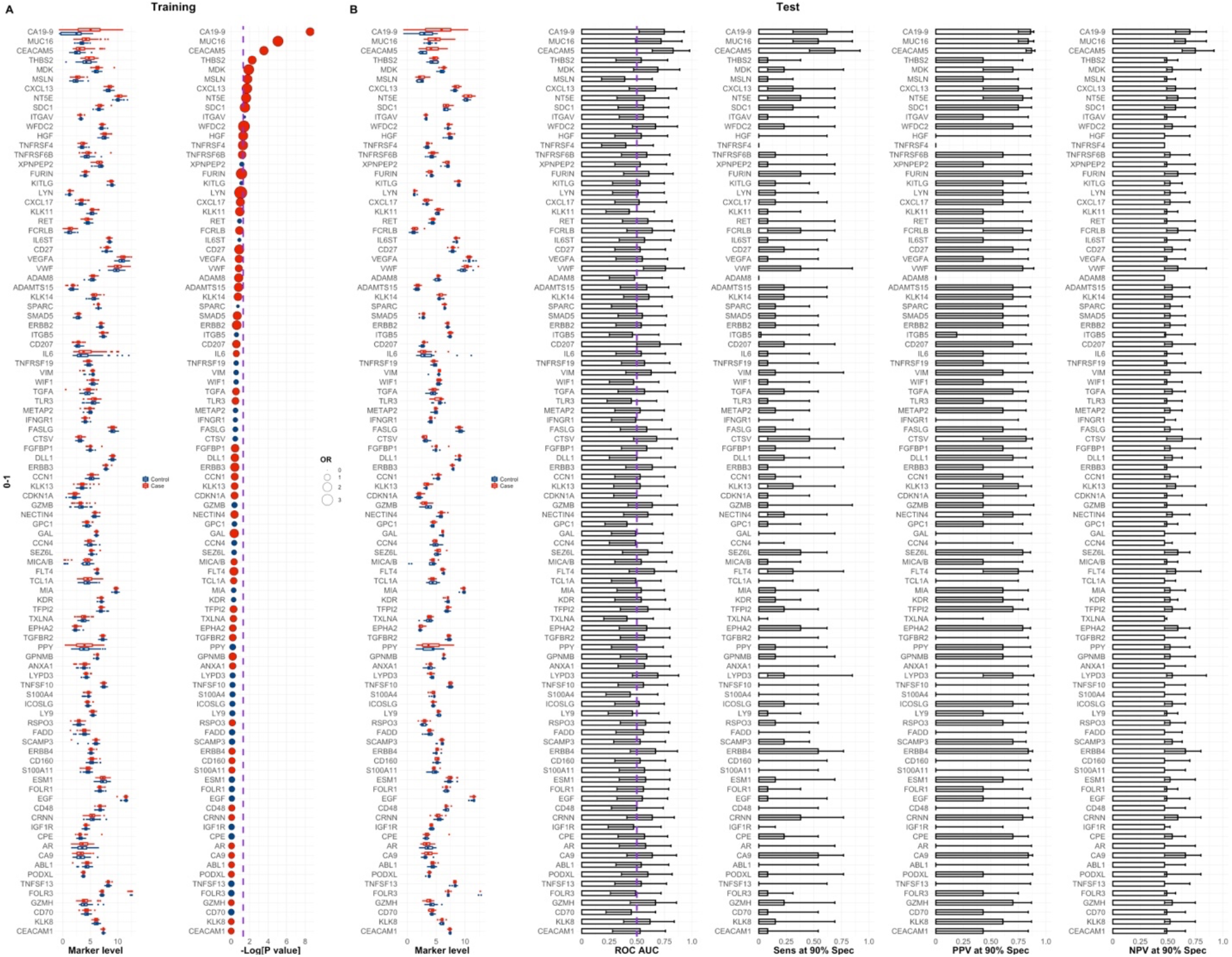
Feature ranks in the training set per single time group according to a logistic regression model with a bias reduction method. Time-group 0-1 years to diagnosis. OR stands for odds-ratio. Red and blue OR points represent OR > 1 (favours PDAC status) and OR < 1 (favours Control status), respectively. See also Methods.

**Fig. S7.**
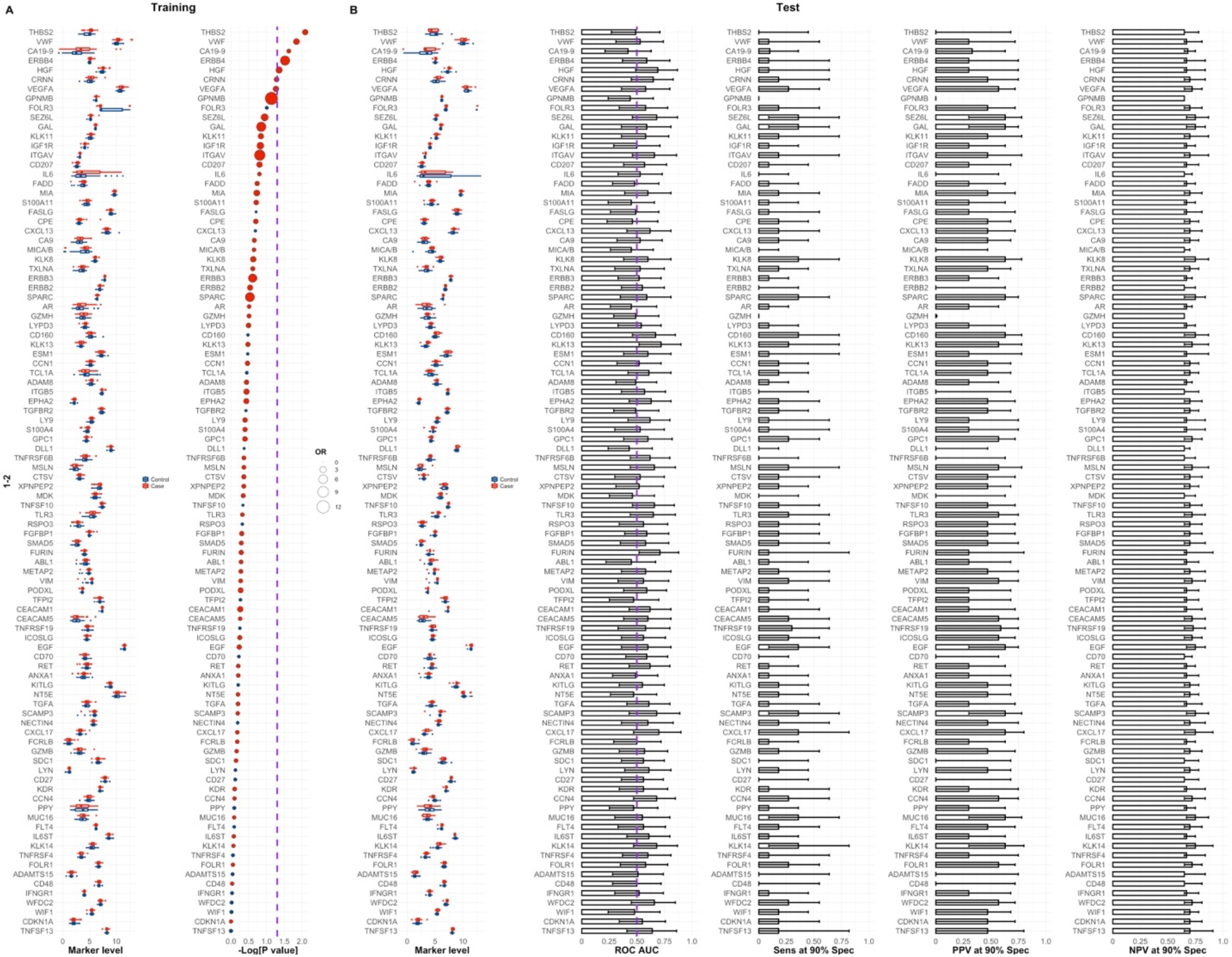
Feature ranks in the training set per time group according to a logistic regression model with a bias reduction method. Time-group 1-2 years to diagnosis. OR stands for odds-ratio. Red and blue OR points represent OR > 1 (favours PDAC status) and OR < 1 (favours Control status), respectively. See also Methods.

**Fig. S8.**
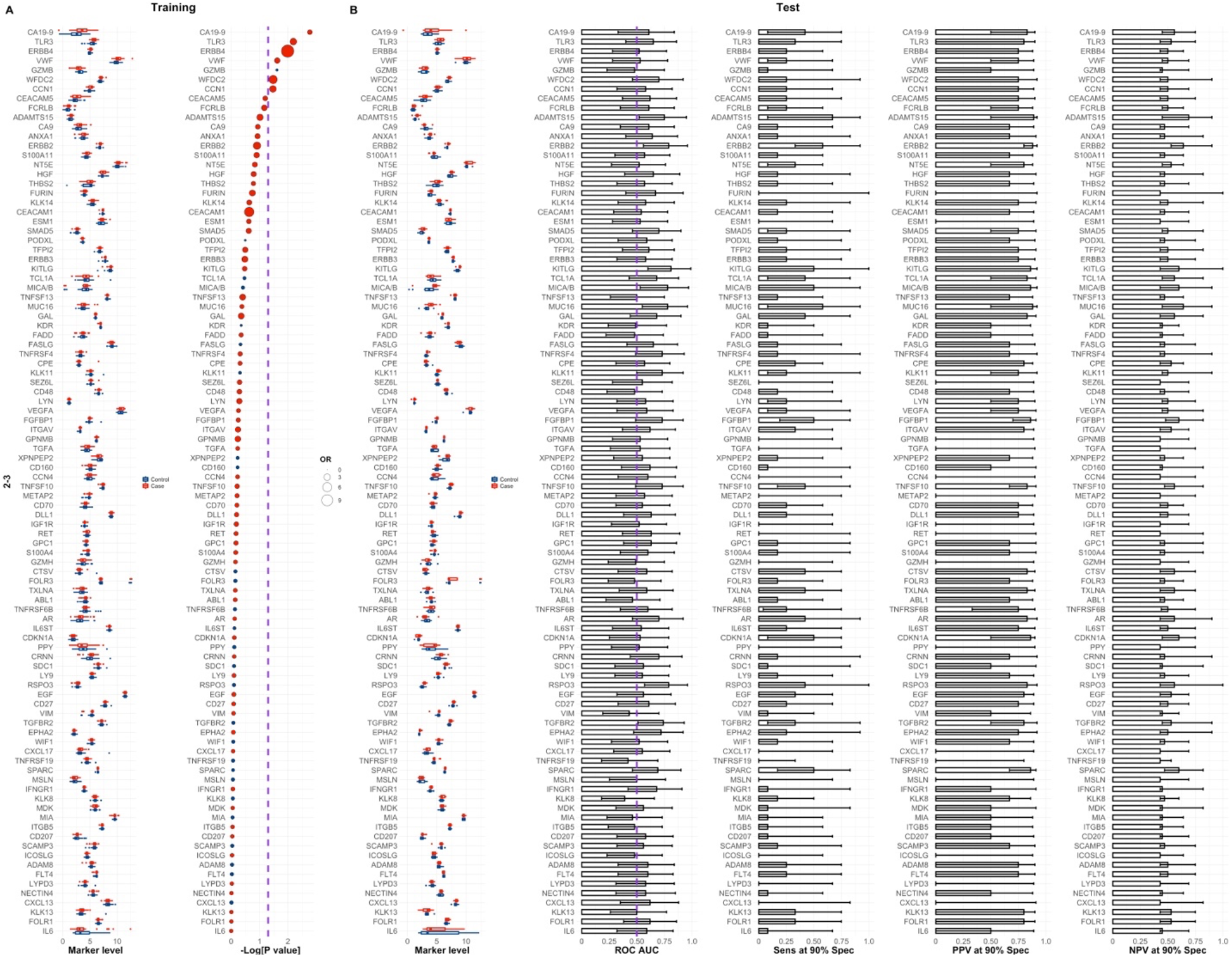
Feature ranks in the training set per time group according to a logistic regression model with a bias reduction method. Time-group 2-3 years to diagnosis. OR stands for odds-ratio. Red and blue OR points represent OR > 1 (favours PDAC status) and OR < 1 (favours Control status), respectively. See also Methods.

**Fig. S9.**
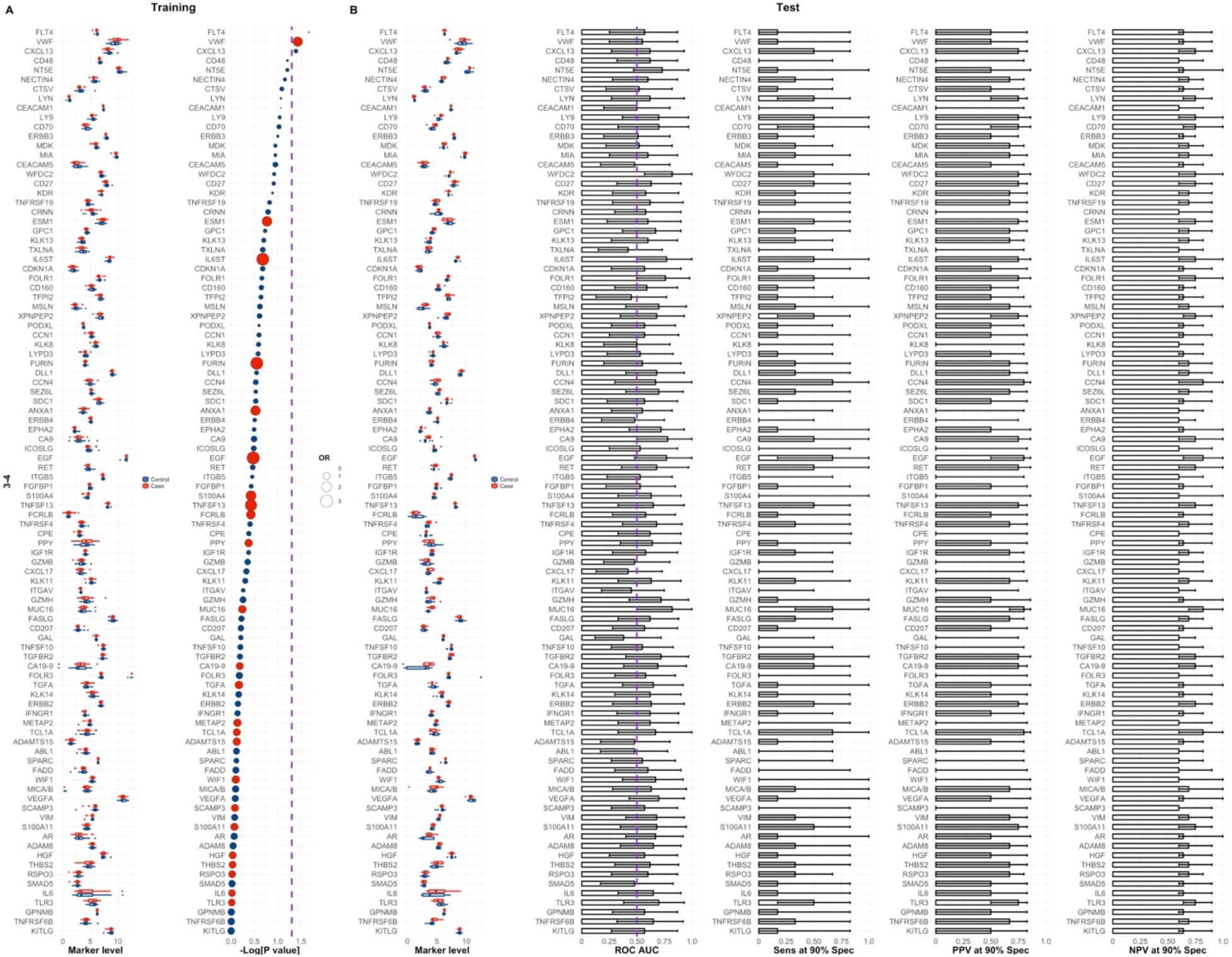
Feature ranks in the training set per time group according to a logistic regression model with a bias reduction method. Time-group 3-4 years to diagnosis. OR stands for odds-ratio. Red and blue OR points represent OR > 1 (favours PDAC status) and OR < 1 (favours Control status), respectively. See also Methods.

**Fig. S10.**
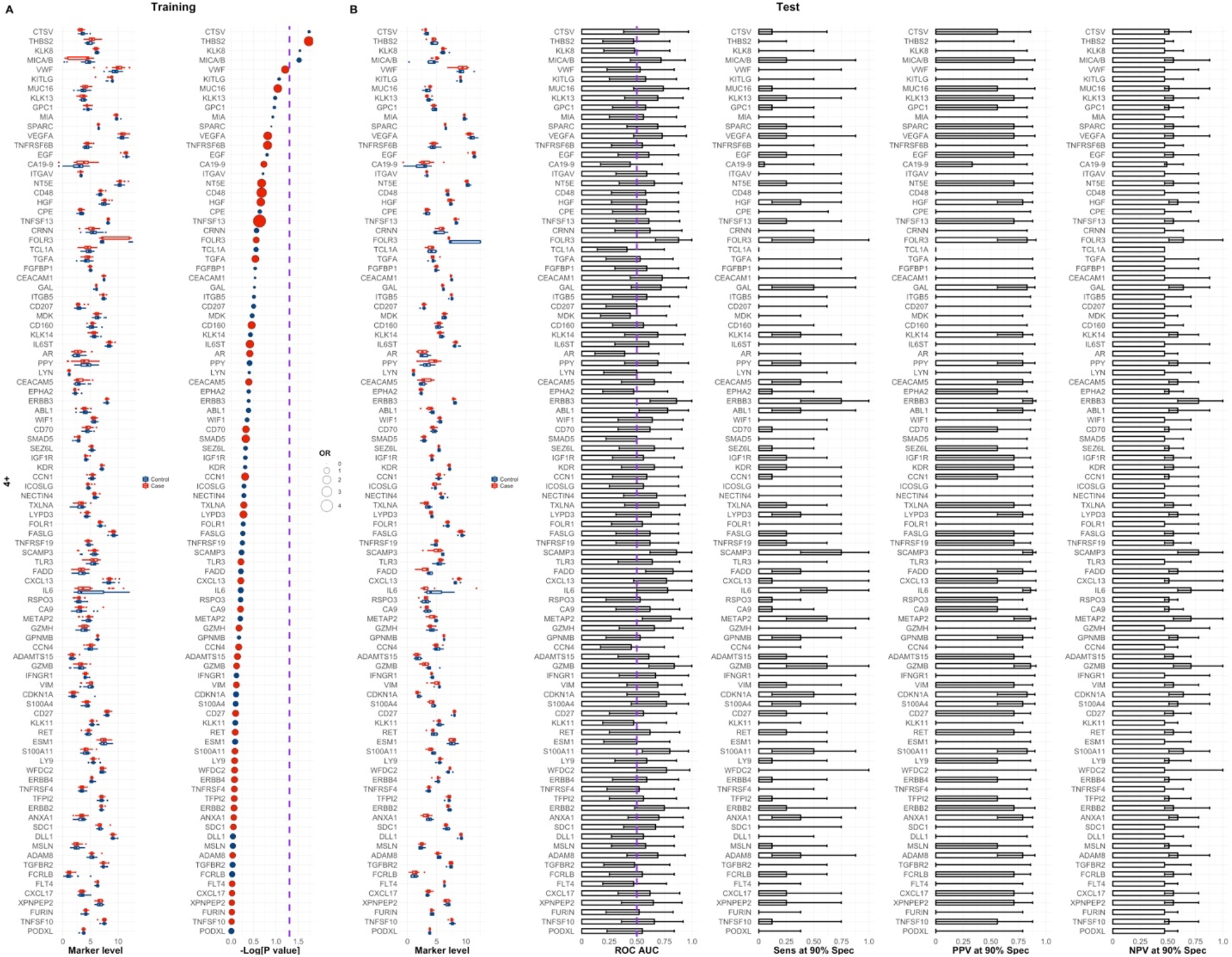
Feature ranks in the training set per time group according to a logistic regression model with a bias reduction method. Time-group 4+ years to diagnosis. OR stands for odds-ratio. Red and blue OR points represent OR > 1 (favours PDAC status) and OR < 1 (favours Control status), respectively. See also Methods.

**Fig. S11.**
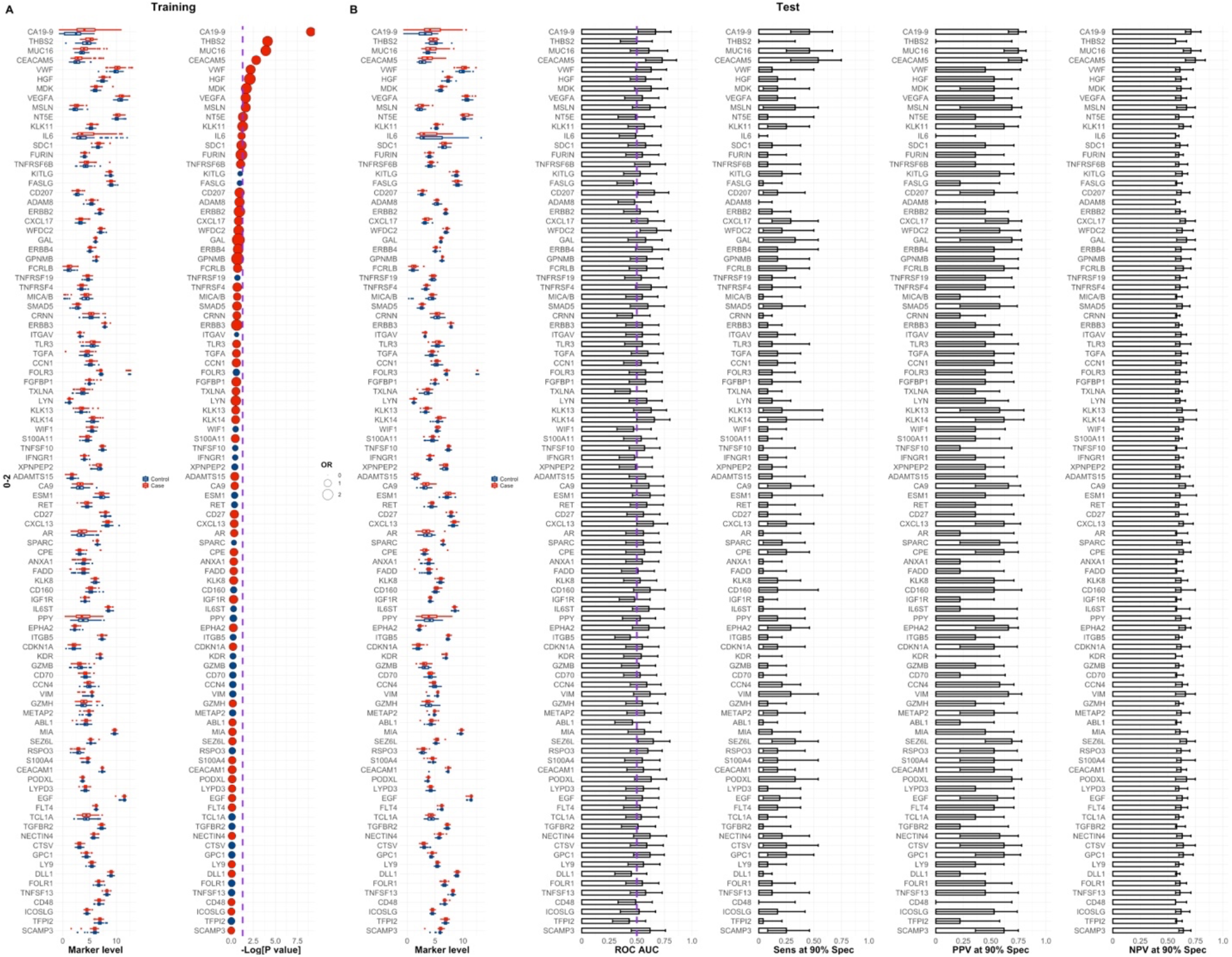
Feature ranks in the training set per joined time group according to a logistic regression model with a bias reduction method. Time-group 0-2 years to diagnosis. OR stands for odds-ratio. Red and blue OR points represent OR > 1 (favours PDAC status) and OR < 1 (favours Control status), respectively. See also Methods.

**Fig. S12.**
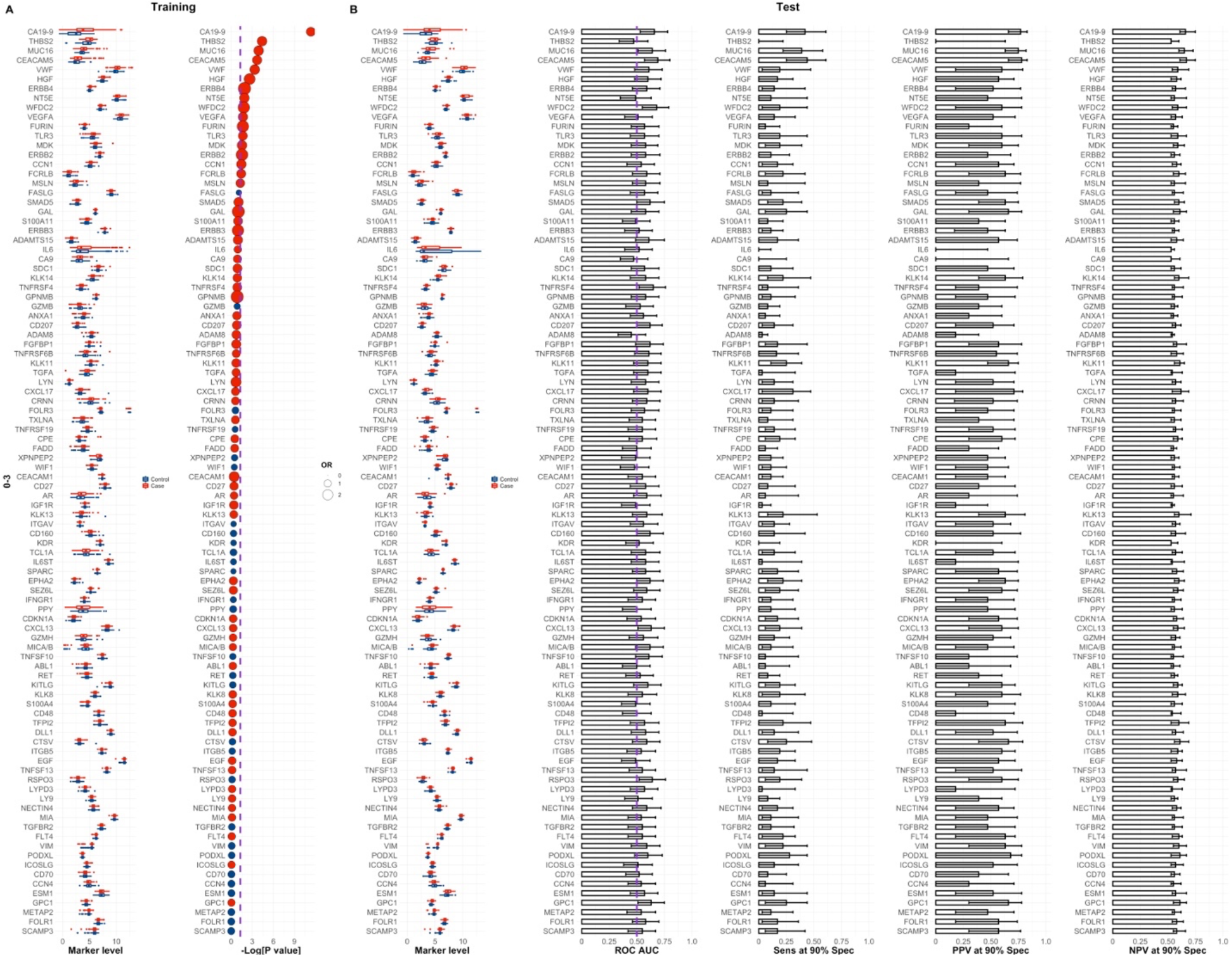
Feature ranks in the training set per joined time group according to a logistic regression model with a bias reduction method. Time-group 0-3 years to diagnosis. OR stands for odds-ratio. Red and blue OR points represent OR > 1 (favours PDAC status) and OR < 1 (favours Control status), respectively. See also Methods.

**Fig. S13.**
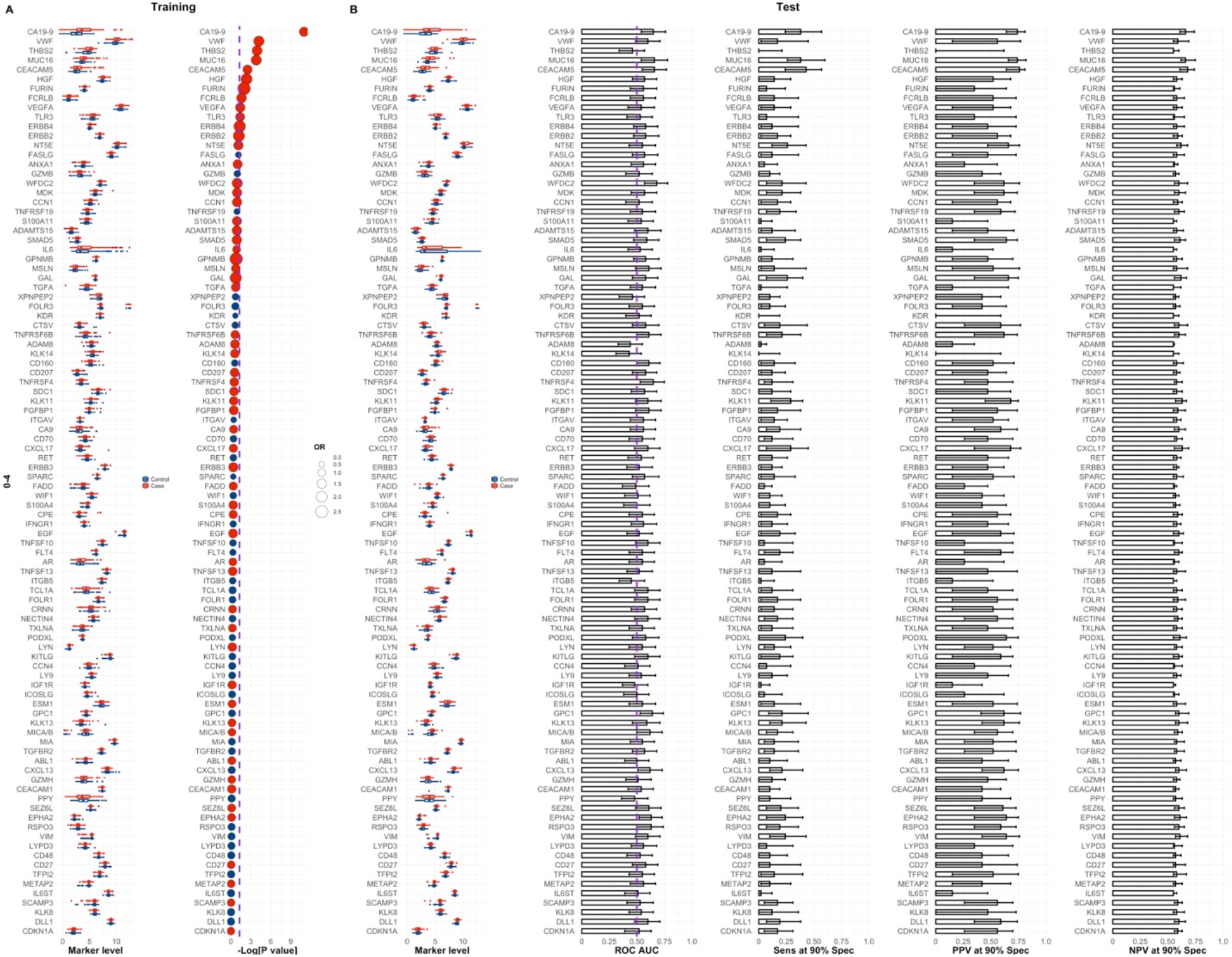
Feature ranks in the training set per joined time group according to a logistic regression model with a bias reduction method. Time-group 0-4 years to diagnosis. OR stands for odds-ratio. Red and blue OR points represent OR > 1 (favours PDAC status) and OR < 1 (favours Control status), respectively. See also Methods.

**Fig. S14.**
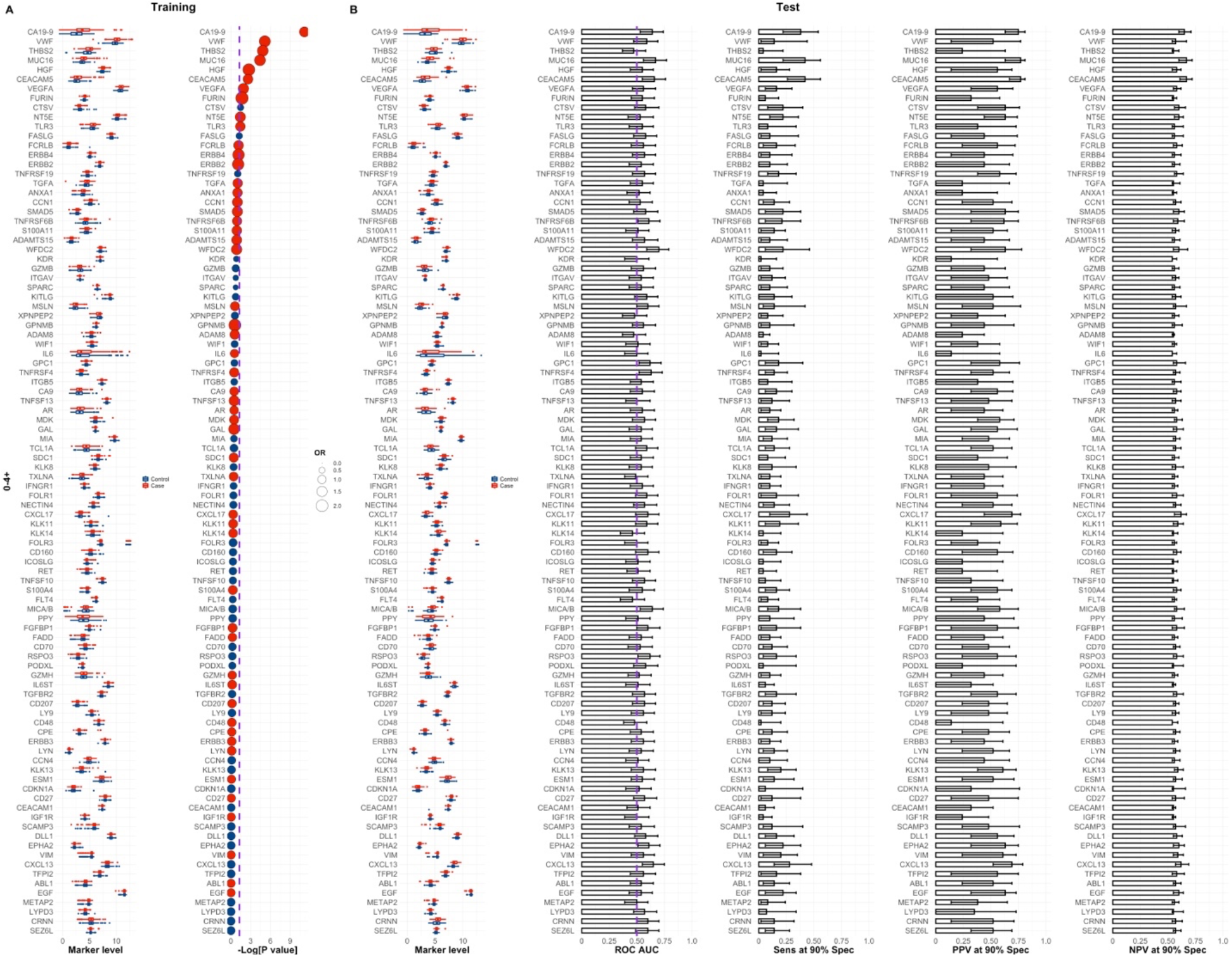
Feature ranks in the training set per joined time group according to a logistic regression model with a bias reduction method. Time-group 0-4+ years to diagnosis. OR stands for odds-ratio. Red and blue OR points represent OR > 1 (favours PDAC status) and OR < 1 (favours Control status), respectively. See also Methods.

**Fig. S15.**
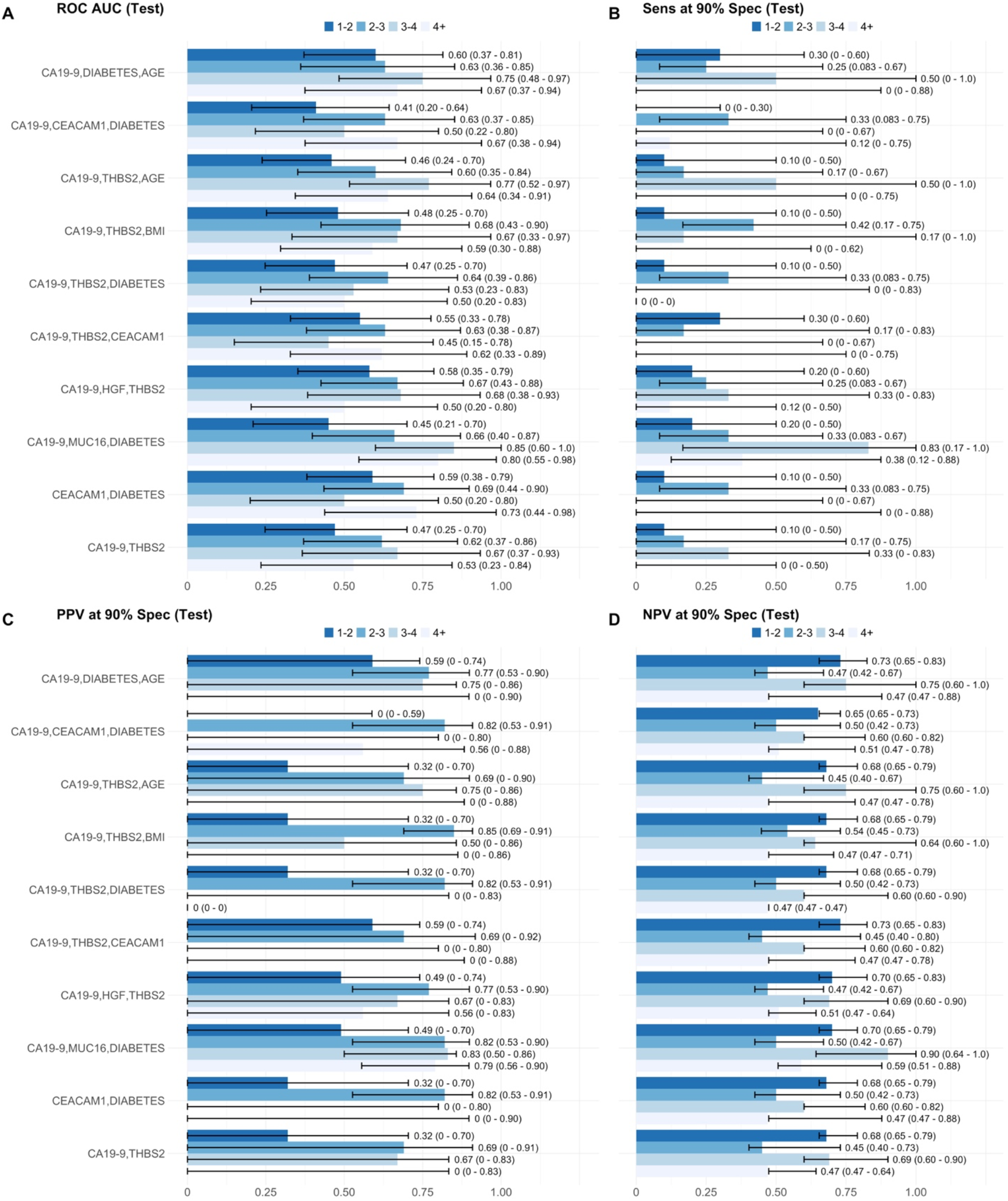
Performances for state-of-the-art biomarker combinations, top models for single time-groups. ROC AUC (Test) values were calculated with the combinations of features developed in the training set. Models are ranked according to their performance in the training set with a 5 times 10-fold cross-validation resampling strategy. Error bars in figures correspond to 95% CI for AUCs, determined by stratified bootstrapping.

**Fig. S16.**
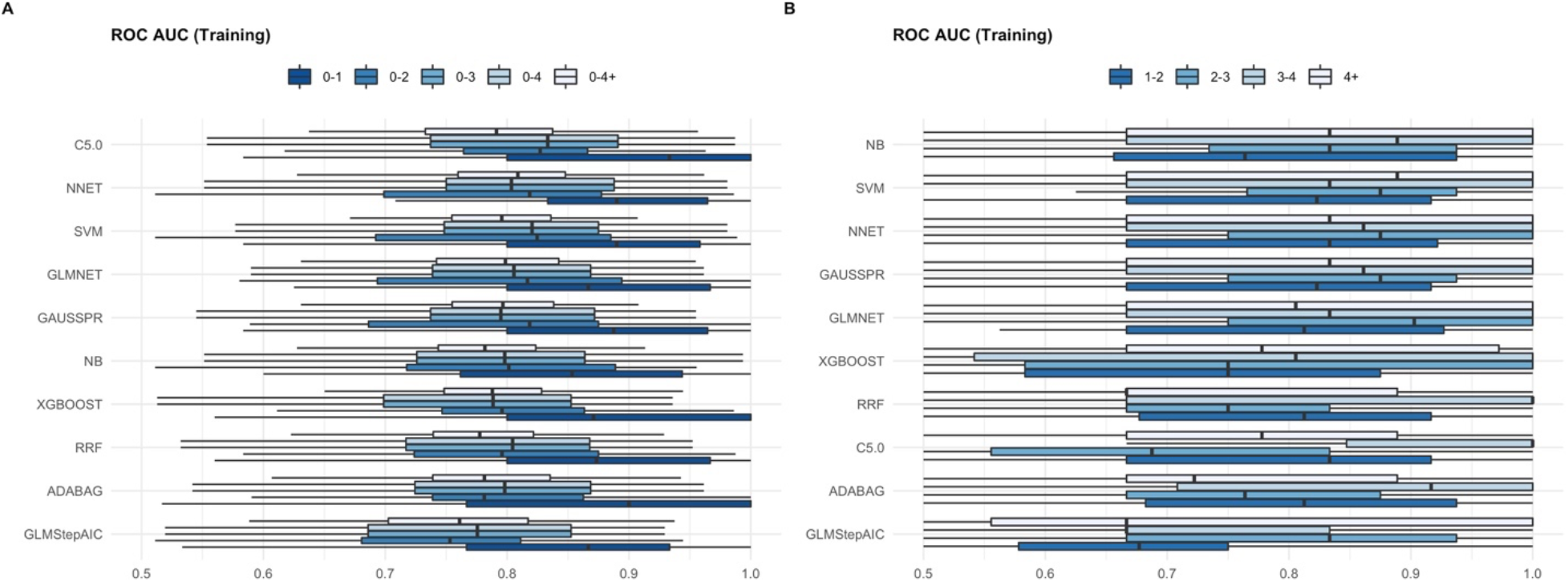
Performance of base-learners across training folds. **A** Joined time-groups. **B** Single time-groups. Results generated with oversampling of minority class. See also Methods.

**Fig. S17.**
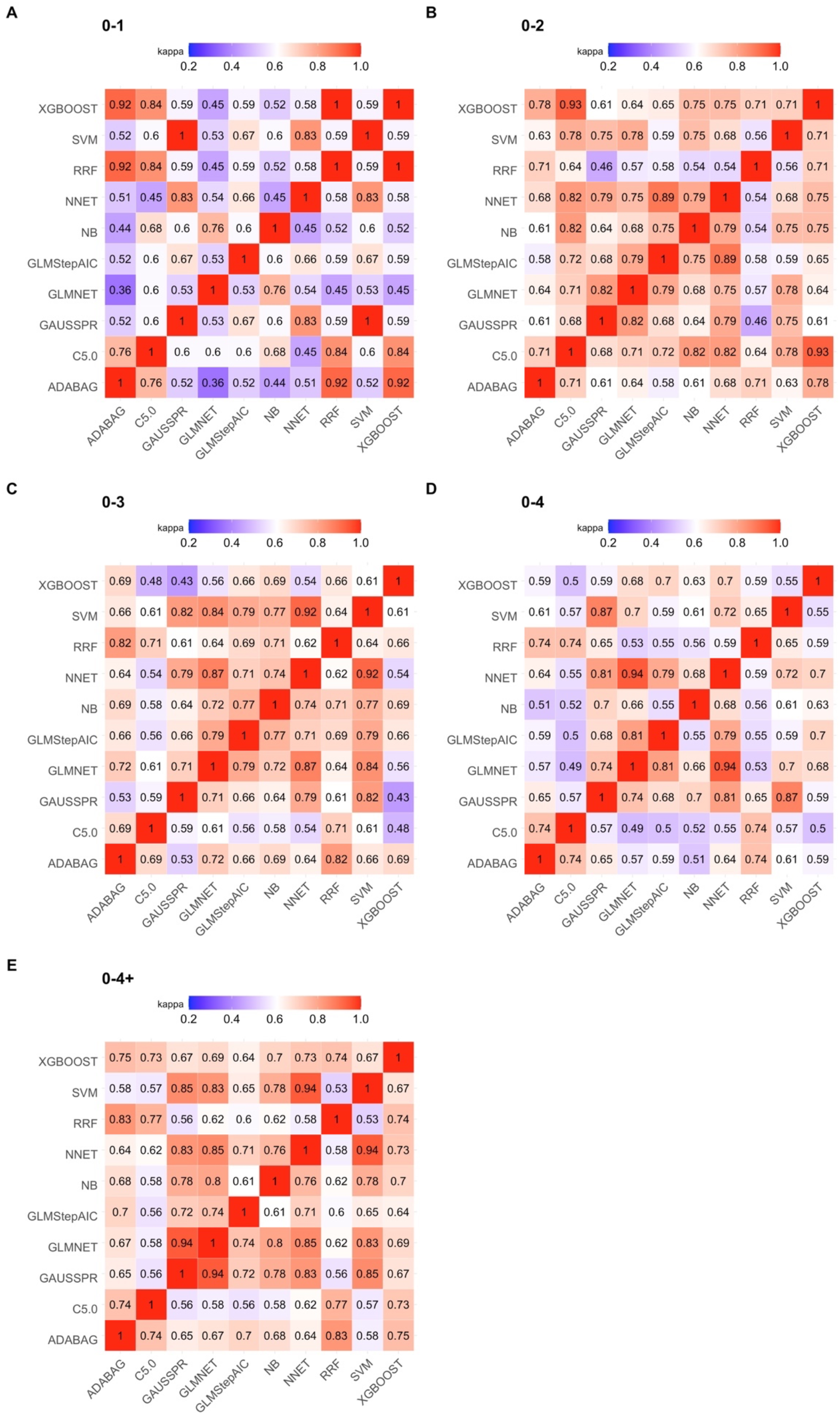
Heterogeneity between base-learners, joined time-groups. Cohen k-statistic (kappa) for pair-wise base-learner predictions in the test set. All values are significant. Results generated with oversampling of minority class. See also Methods.

**Fig. S18.**
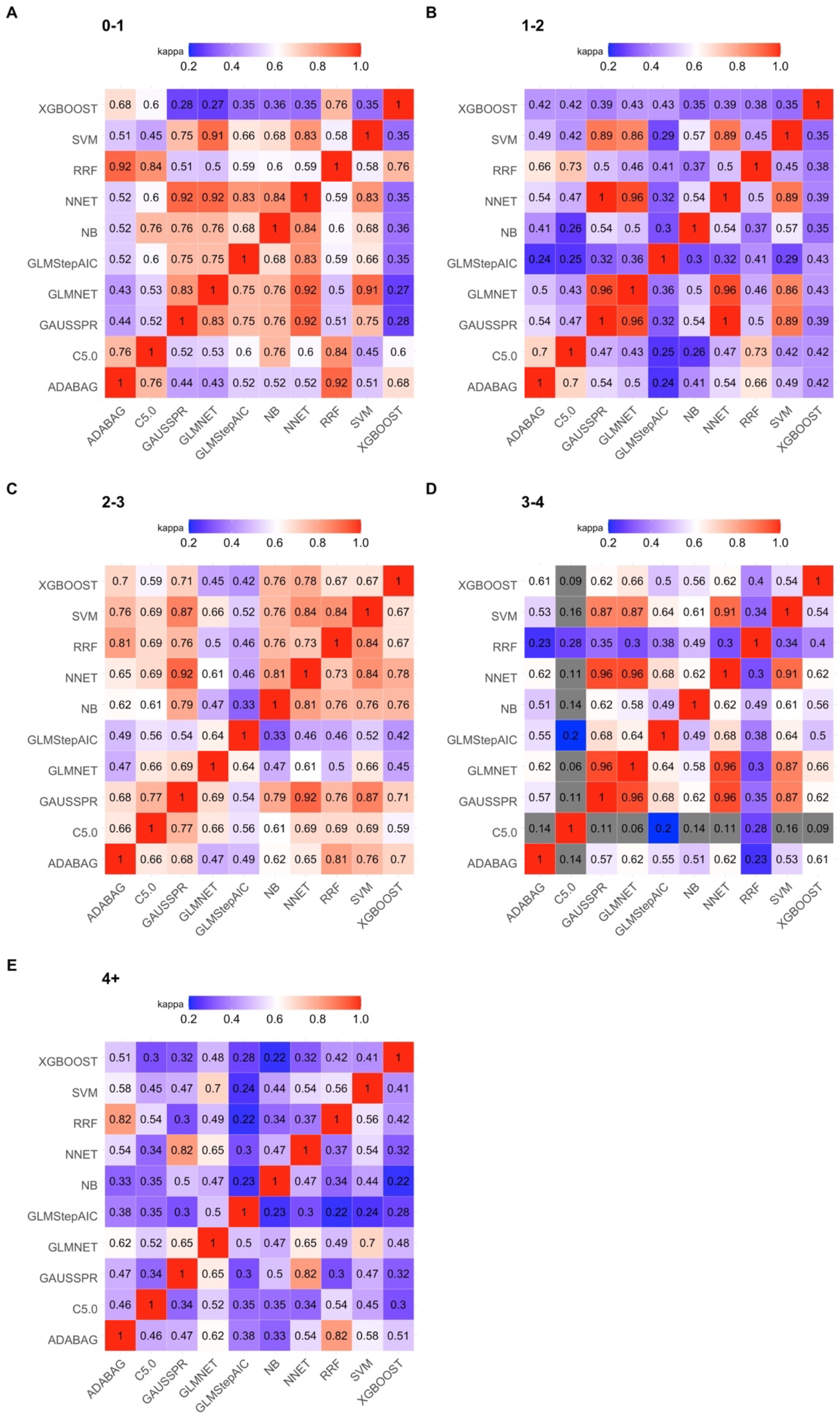
Heterogeneity between base-learners, single time-groups. Cohen k-statistic (kappa) for pair-wise base-learner predictions in the test set. All values are significant. Results generated with oversampling of minority class. See also Methods.

**Fig. S19.**
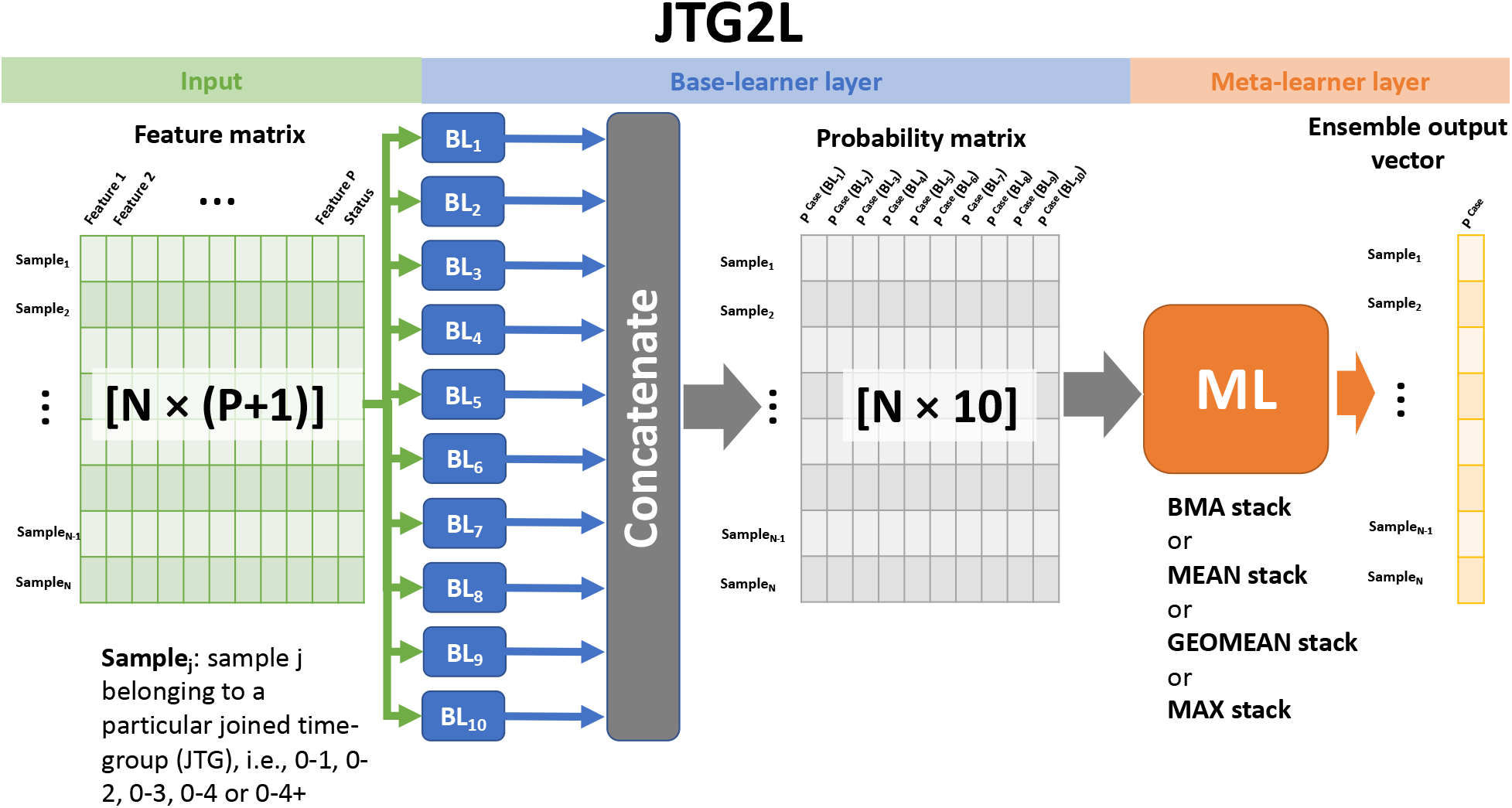
Flow diagram for the joined time-group ensemble classifier, JTG2L. Each JTG2L model presented in the main text, e.g., a model developed by training with 0-1 samples, was built according to this diagram. The feature matrix is used to train each of the base-learners. The output of this step is a probability vector of length N for each base-learner, where each entry corresponds to the probability of being a case according to the respective base-learner. The set of 10 probability vectors is then concatenated to generate a probability matrix that is used to train the meta-learner. From this a final probability output vector is computed. The base-learners and the meta-learner are trained with the resampling techniques highlighted in the methods section (main text). For the purposes of applying the resulting trained models to the test set, the flow of the diagram is the same as before but the feature matrix will have a different number of samples.

**Fig. S20.**
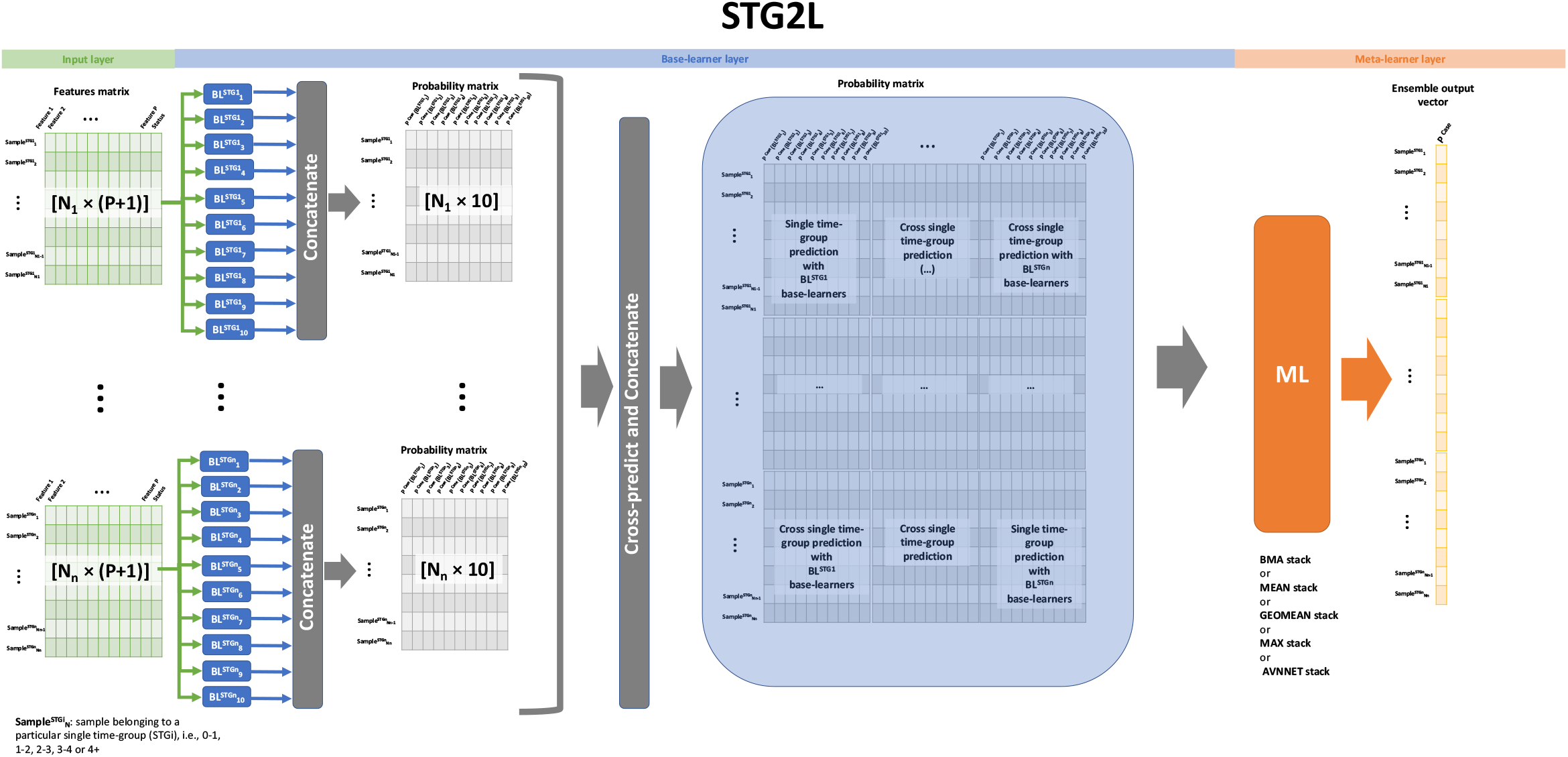
Flow diagram for the specialized single time-group ensemble classifier, STG2L. Each STG2L model presented in the main text, e.g., a model developed by training with 0-2 samples, was built according to this diagram, where base-learners are trained in single time-groups and subsequently stacked. The stacking procedure has 2 steps. First, the probability output vectors for each base-learner trained in each single time-group are concatenated, thus leading to n probability matrices, where n is the number of single time-groups within a joined time group that we want to predict in the test set. If we are predicting PDAC status in 0-2 samples, we will have 2 matrices. Second, these matrices are subsequently used to populate the diagonal blocks of a larger probability matrix. The off-diagonal probability blocks are generated by using the base models trained in a specific single time-group, which therefore amounts to computing cross time-group predictions. The resulting large matrix has 10*n columns and is then used to train the meta-learner which outputs the final vector. For the purposes of applying the resulting trained models to the test set, the flow of the diagram is the same as before, but the feature matrix will have a different number of samples.

**Fig. S21.**
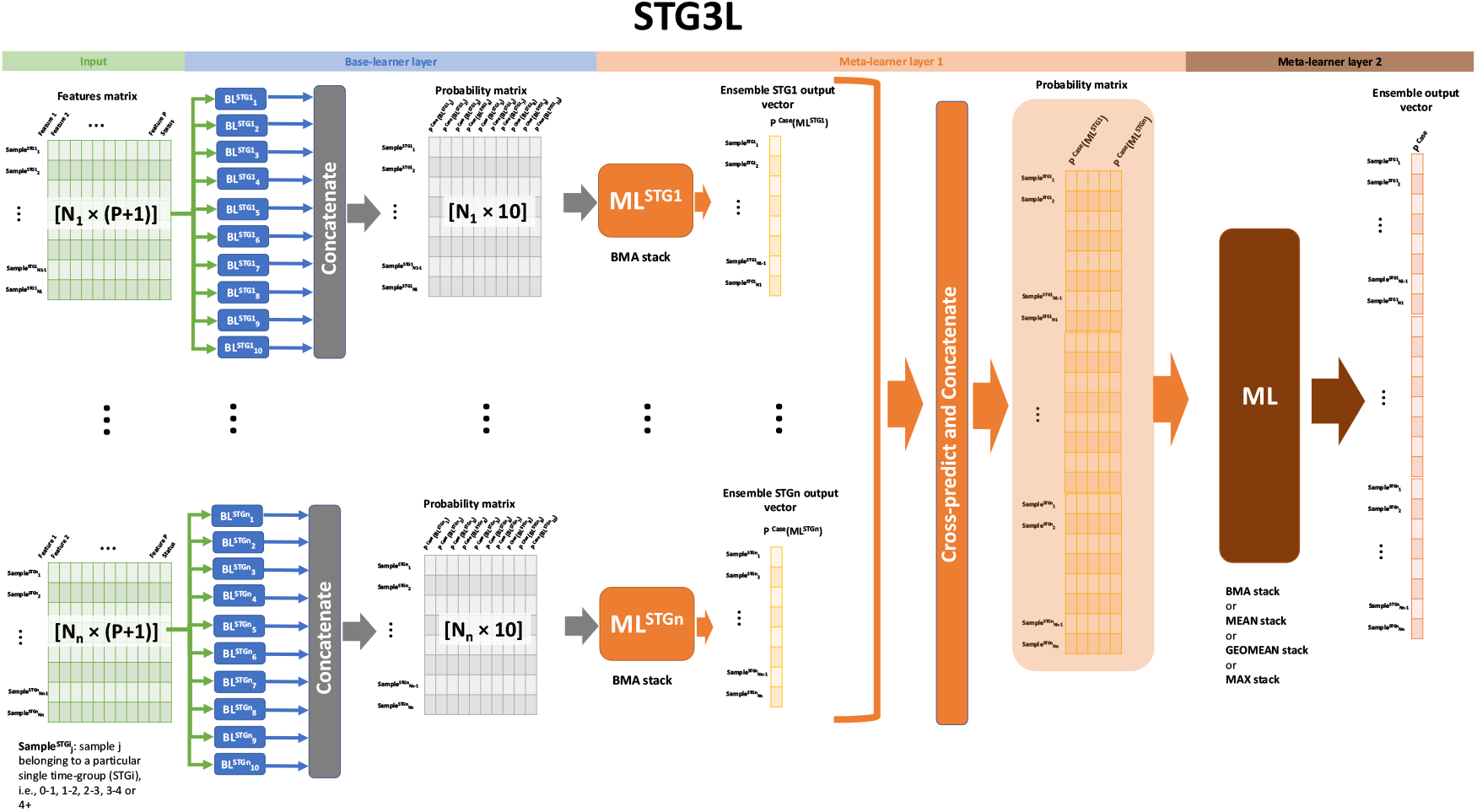
Flow diagram for the specialized single time-group ensemble classifier, STG3L. Each STG3L model presented in the main text, e.g., a model developed by training with 0-2 samples, was built according to this diagram, where base-learners are trained in single time-groups and subsequently stacked. The stacking procedure has 3 steps. First, the probability output vectors for each base-learner trained in each single time-group are concatenated, thus leading to n probability matrices, where n is the number of single time-groups within a joined time group that we want to predict in the test set. If we are predicting PDAC status in 0-2 samples, we will have 2 matrices. Second, these matrices are used to train a BMA stack outputting 1-column probability vectors. From this intermediate step we have to populate the diagonal 1-column blocks of a larger probability matrix with n columns. The off-diagonal probability blocks are generated by using the base models and intermediate BMA stack trained in a specific single time-group, thus leading to cross time-group predictions. For the purposes of applying the resulting trained models to the test set, the flow of the diagram is the same as before, but the feature matrix will have a different number of samples.

**Fig. S22.**
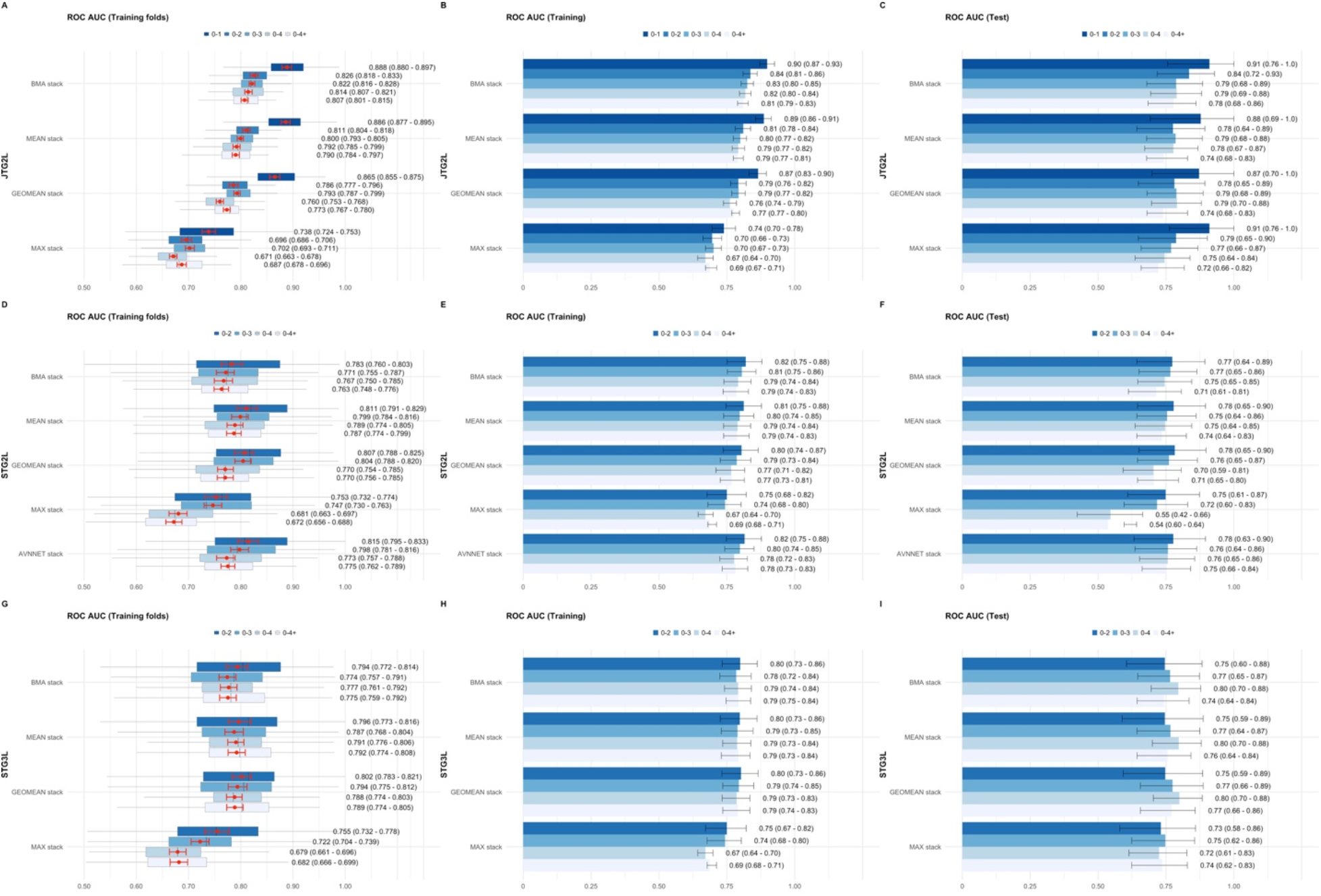
Performance for each of the meta-learners and models tested in this work. **A** ROC AUC across training folds for the JTG2L model. Red error bars and dot represent the 95% CI for the mean and the mean, generated by bootstrapping with the *boot* R package (version 1.3-25)**. B** ROC AUC when the underlying base-learners and the stack is fitted to the full training set. C ROC AUC in the test set with the respective model developed in the training set**. D, E** and **F** STG2L model**. G, H** and **I** STG3L model. All results were obtained with oversampling of minority class. See Methods section for details.

**Fig. S23.**
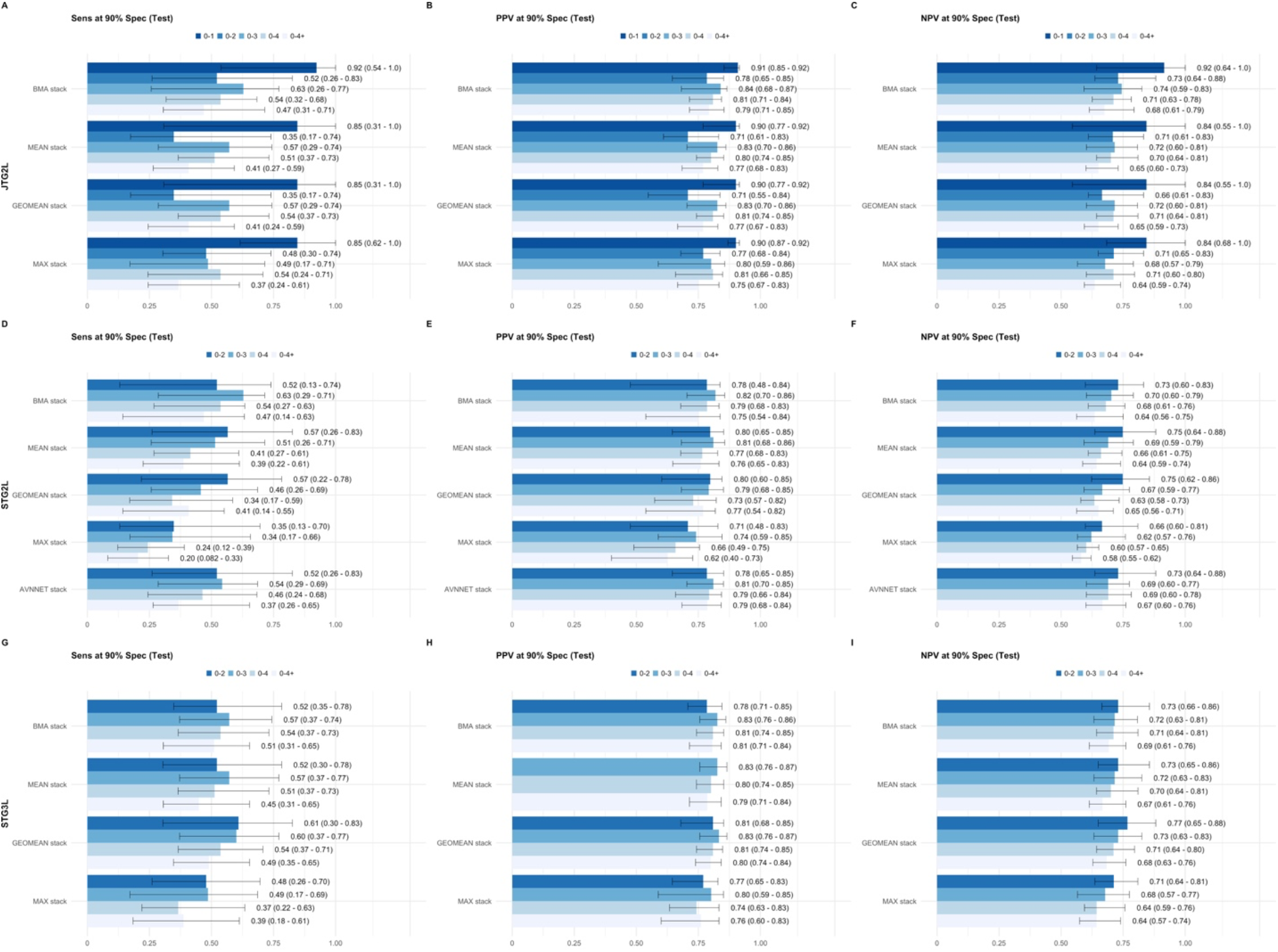
Sensitivity, positive and negative predictive value for meta-learners and models tested in this work. **A** Sensitivity, **B** Positive predictive value (PPV) and **C** Negative predictive value (NPV) at 90% specificity for the JTG2L model. **D, E** and **F** STG2L model**. G, H** and **I** STG3L model. All results were obtained with oversampling of minority class. See Methods section for details.

**Fig. S24.**
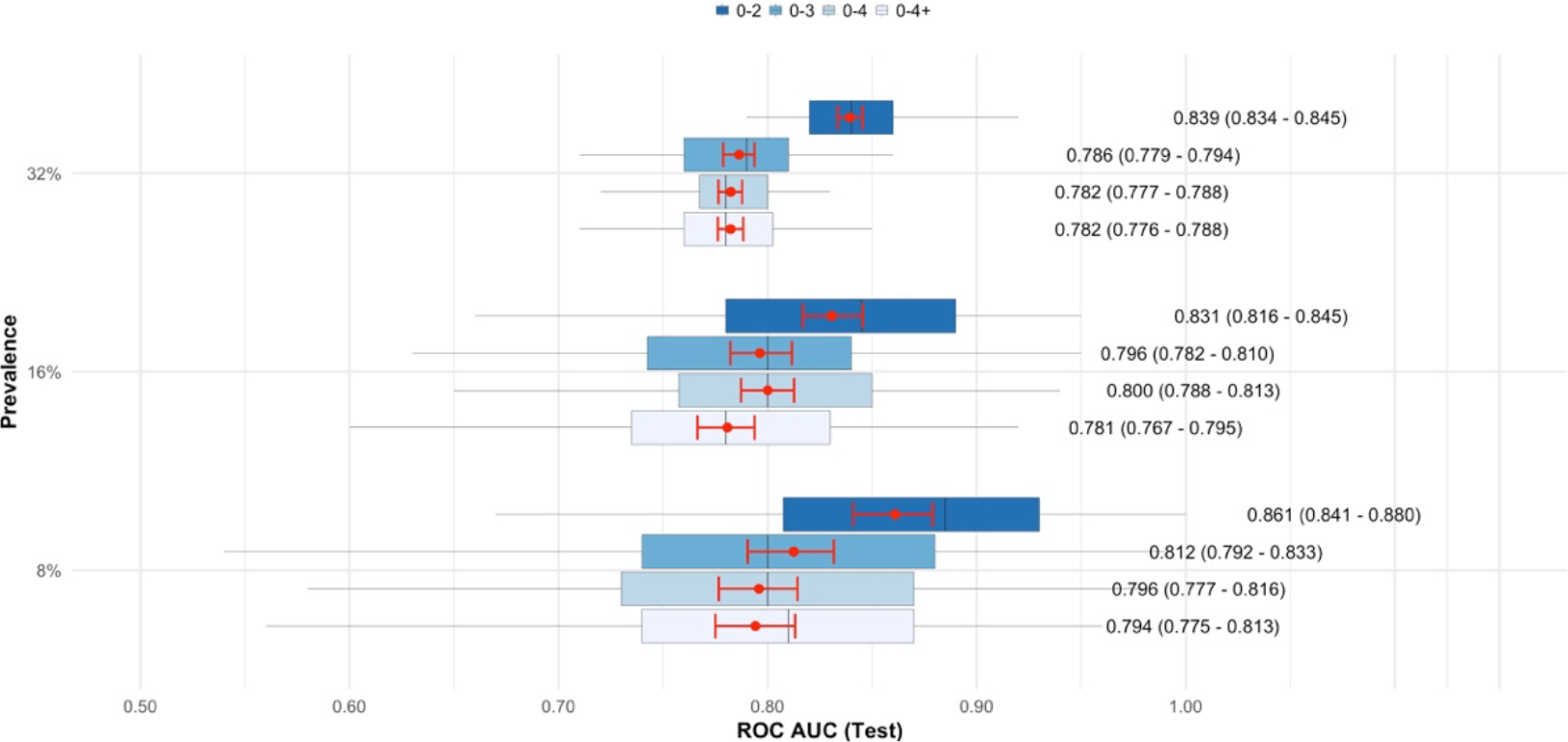
Performance at different prevalence values for the JTG2L BMA stack. Prevalence is determined by the #Cases/(#Cases+#Controls) ratio times 100. For each prevalence value, we select randomly (Prevalence* #Controls/(1-Prevalence)) Cases from the pool of Cases in the test set and determine the performance of the classifiers by using these Cases against the full set of Controls in the test set. This was repeated 1000 times. Red error bars and dot represent the 95% CI for the mean and the mean, generated by bootstrapping with the *boot* R package (version 1.3-25). The 0-1 time-group was not tested since the number of cases was not sufficient to perform the study at 8% and 16% prevalence. Results generated with oversampling of minority class.

**Fig. S25.**
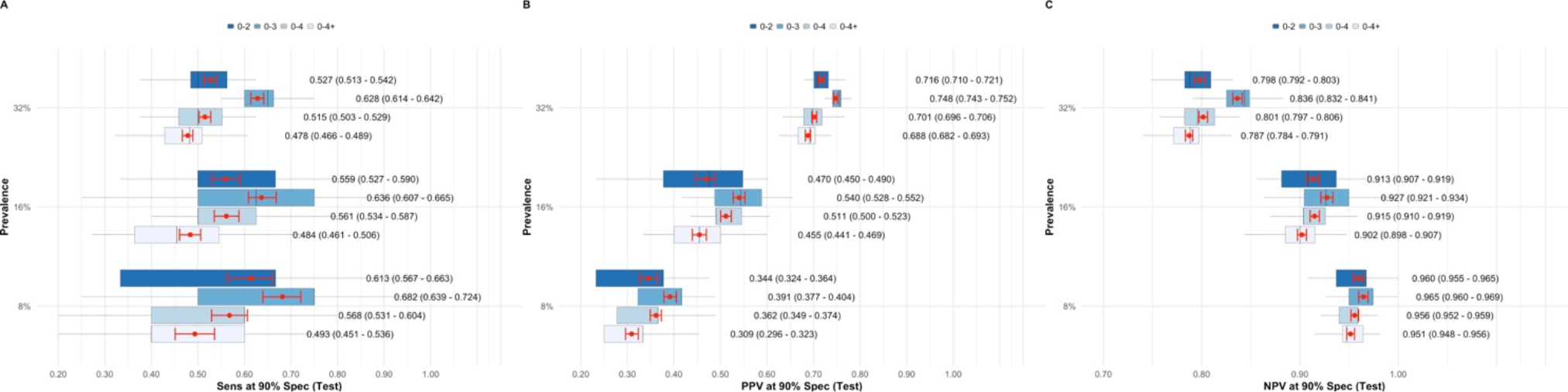
Sensitivity, positive predictive and negative predictive value at different prevalence values for the JTG2L BMA stack. Prevalence is determined by the #Cases/(#Cases+#Controls) ratio times 100. For each prevalence value, we select randomly (Prevalence* #Controls/(1-Prevalence)) Cases from the pool of Cases in the test set and determine the performance of the classifiers by using these Cases against the full set of Controls in the test set. This was repeated 1000 times. Red error bars and dot represent the 95% CI for the mean and the mean, generated by bootstrapping with the *boot* R package (version 1.3-25). The 0-1 time-group was not tested since the number of cases was not sufficient to perform the study at 8% and 16% prevalence. Results generated with oversampling of minority class.

**Fig. S26.**
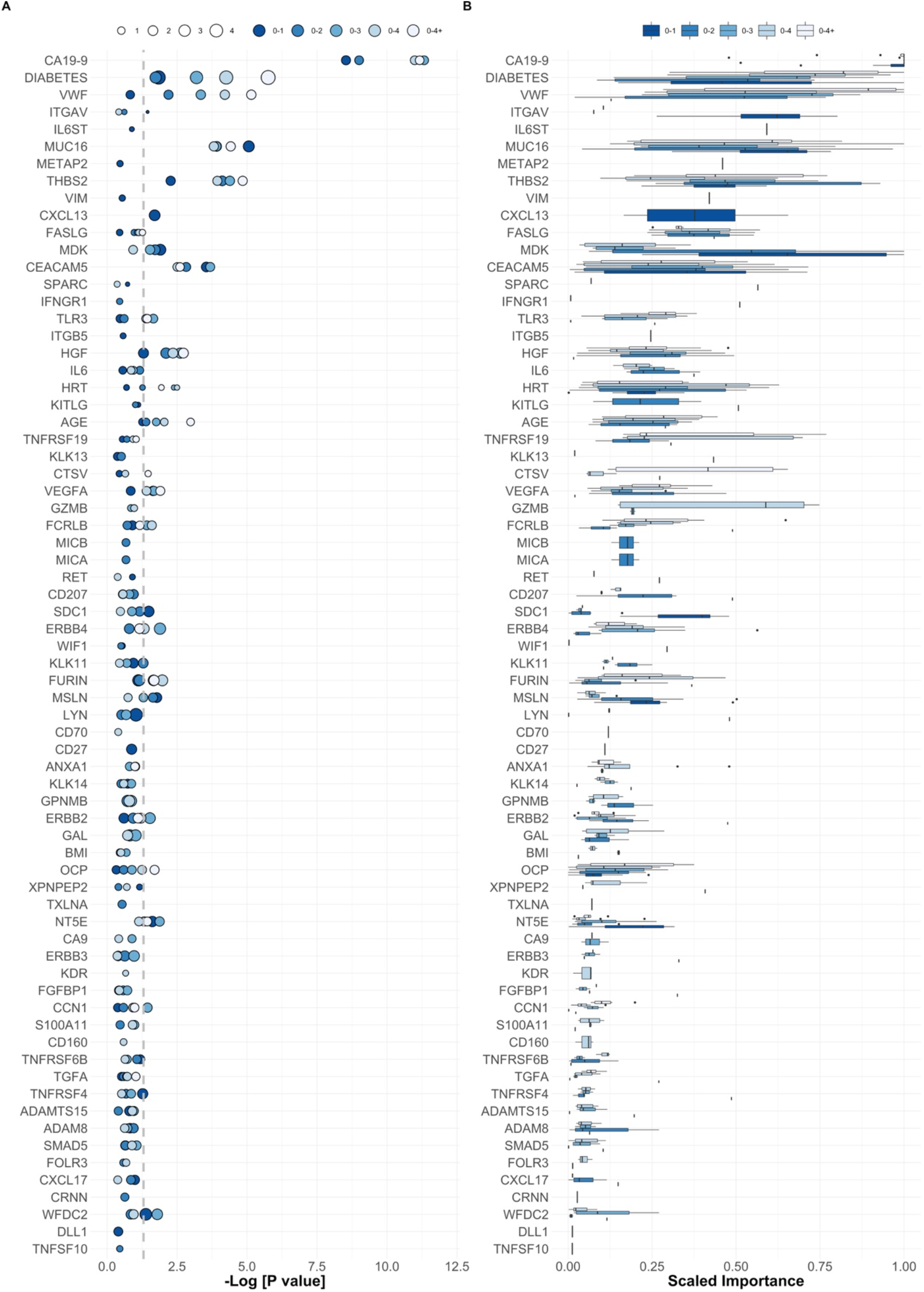
Signatures for each joined time-group across all base-learners. **A** P values calculated according to a logistic regression model with a bias reduction method for each of the features in a one-dimensional model (see Methods). Size corresponds to Odds ratio. **B** Feature importance across base-learners according to a model-agnostic method based on a simple feature importance ranking measure, implemented in the R package *vip* (https://cran.r-roject.org/web/packages/vip/index.html). Results generated with oversampling of minority class.

**Fig. S27.**
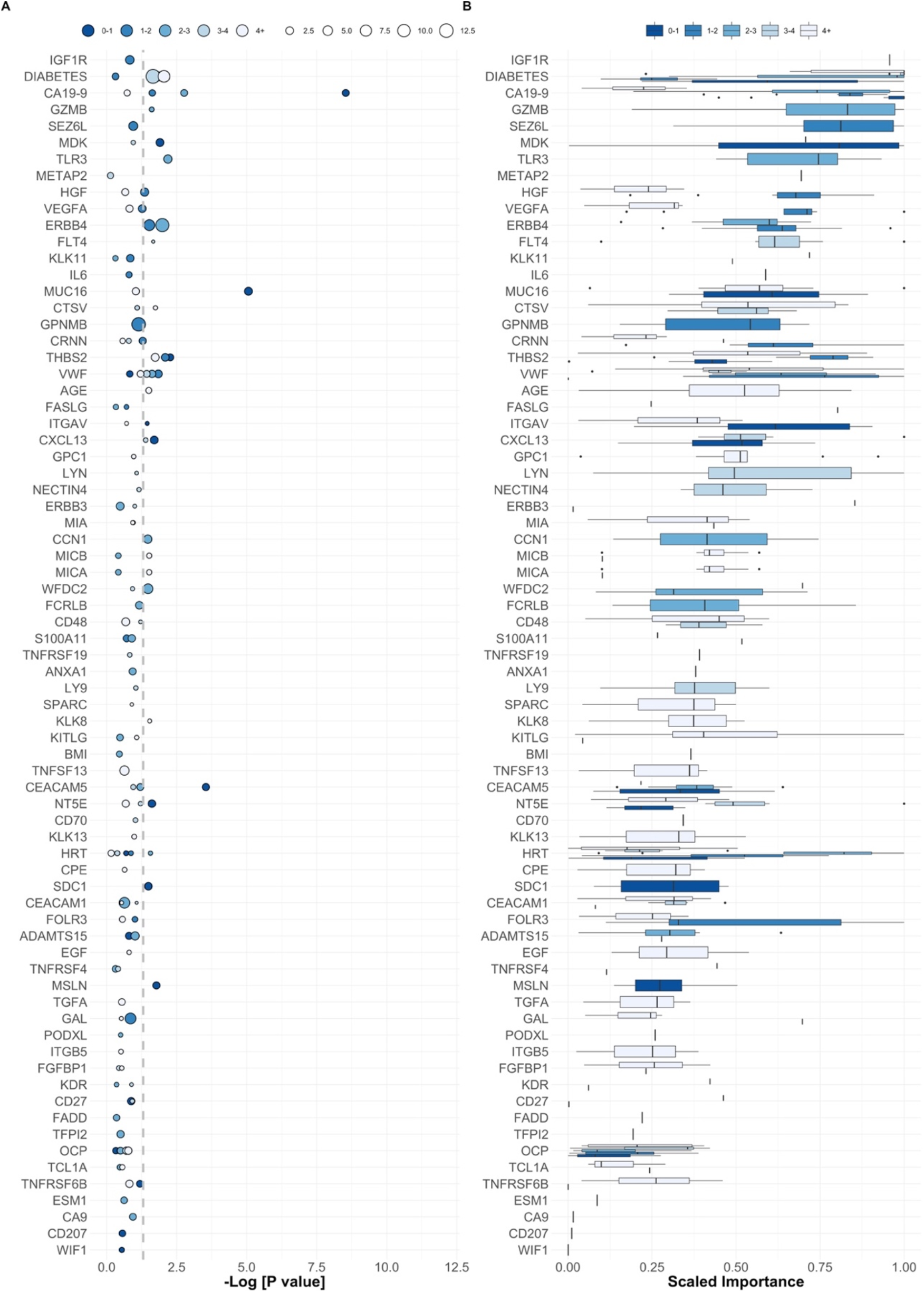
Signatures for each single time-group across all base-learners. **A** P values calculated according to a logistic regression model with a bias reduction method for each of the features in a one-dimensional model (see Methods). Size corresponds to Odds ratio. **B** Feature importance across base-learners according to a model-agnostic method based on a simple feature importance ranking measure, implemented in the R package *vip* (https://cran.r-roject.org/web/packages/vip/index.html). Results generated with oversampling of minority class.

**Fig. S28.**
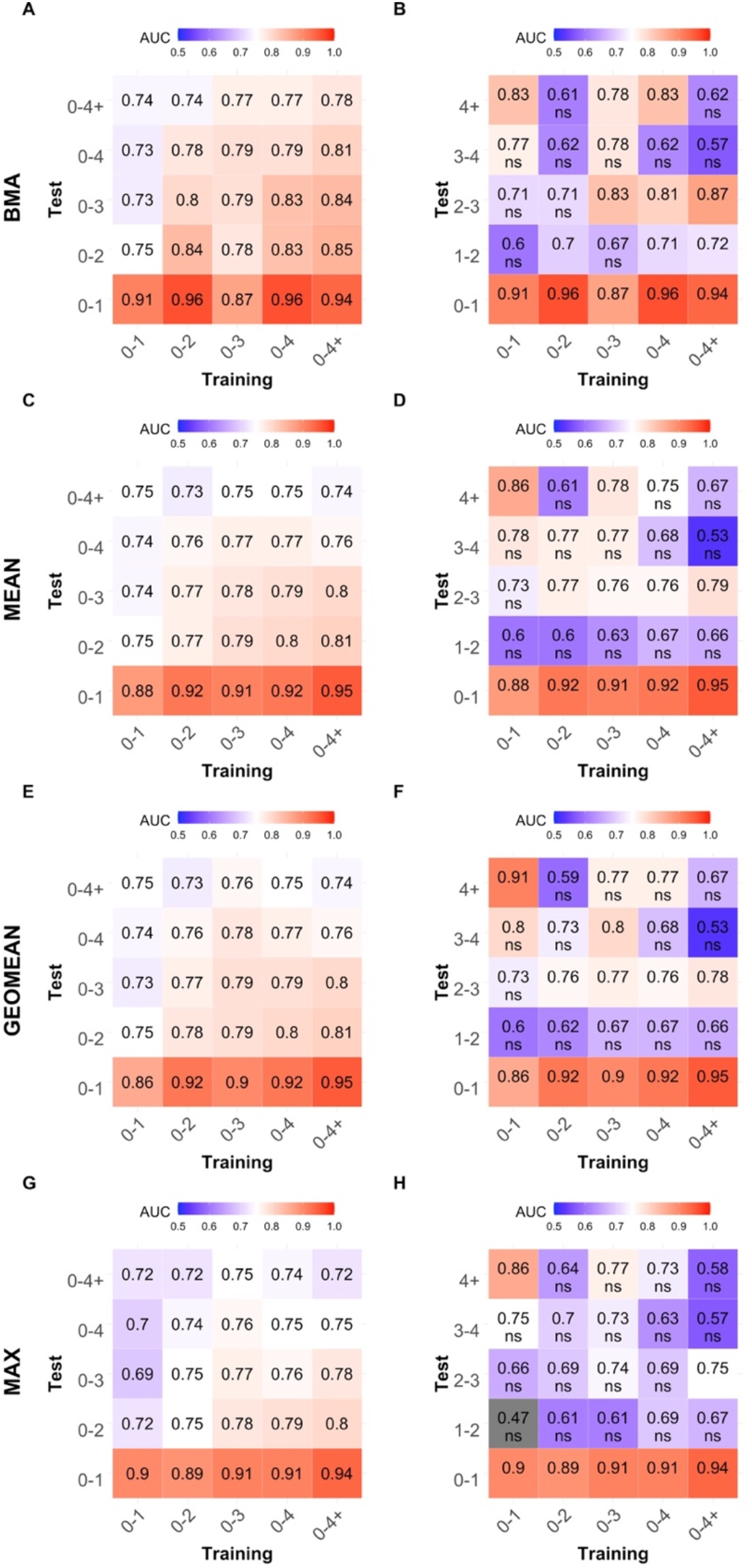
Cross time-group performance for meta-learners, JTG2L. **A** and **B.** BMA meta-learner trained in joined time-groups, predicting joined time-groups and single time-groups, respectively. **C** and **D.** MEAN meta-learner. **E** and **F.** GEOMEAN meta-learner. **G** and **H.** MAX meta-learner. ns represents performance whose 95% CI lower limit crosses the 0.5 threshold. Results generated with oversampling of minority class.

**Fig. S29.**
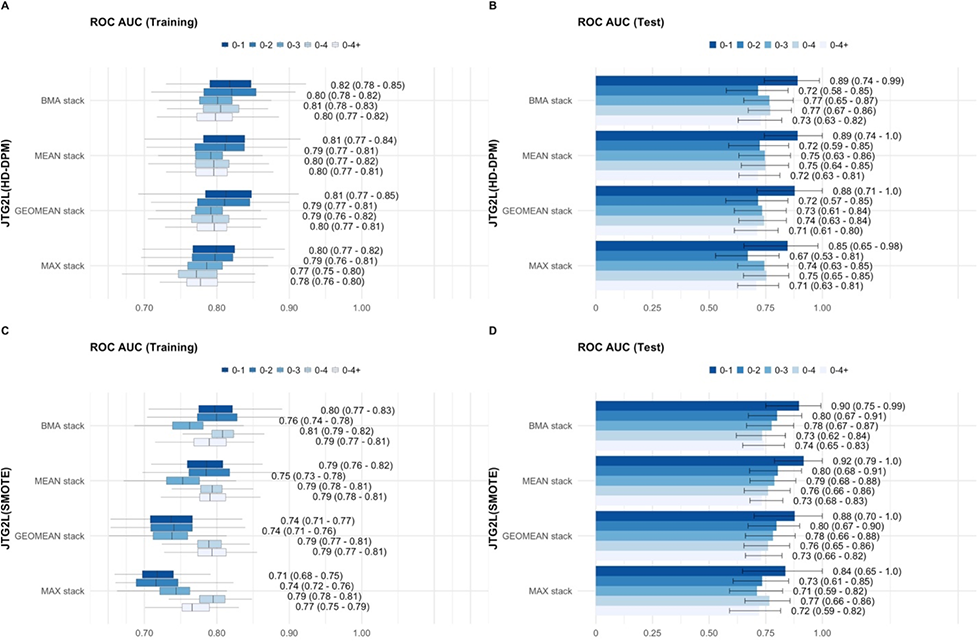
Performance for meta-learners with HD-DPM and SMOTE during resampling, JTG2L. See Methods for details.

**Fig. S30.**
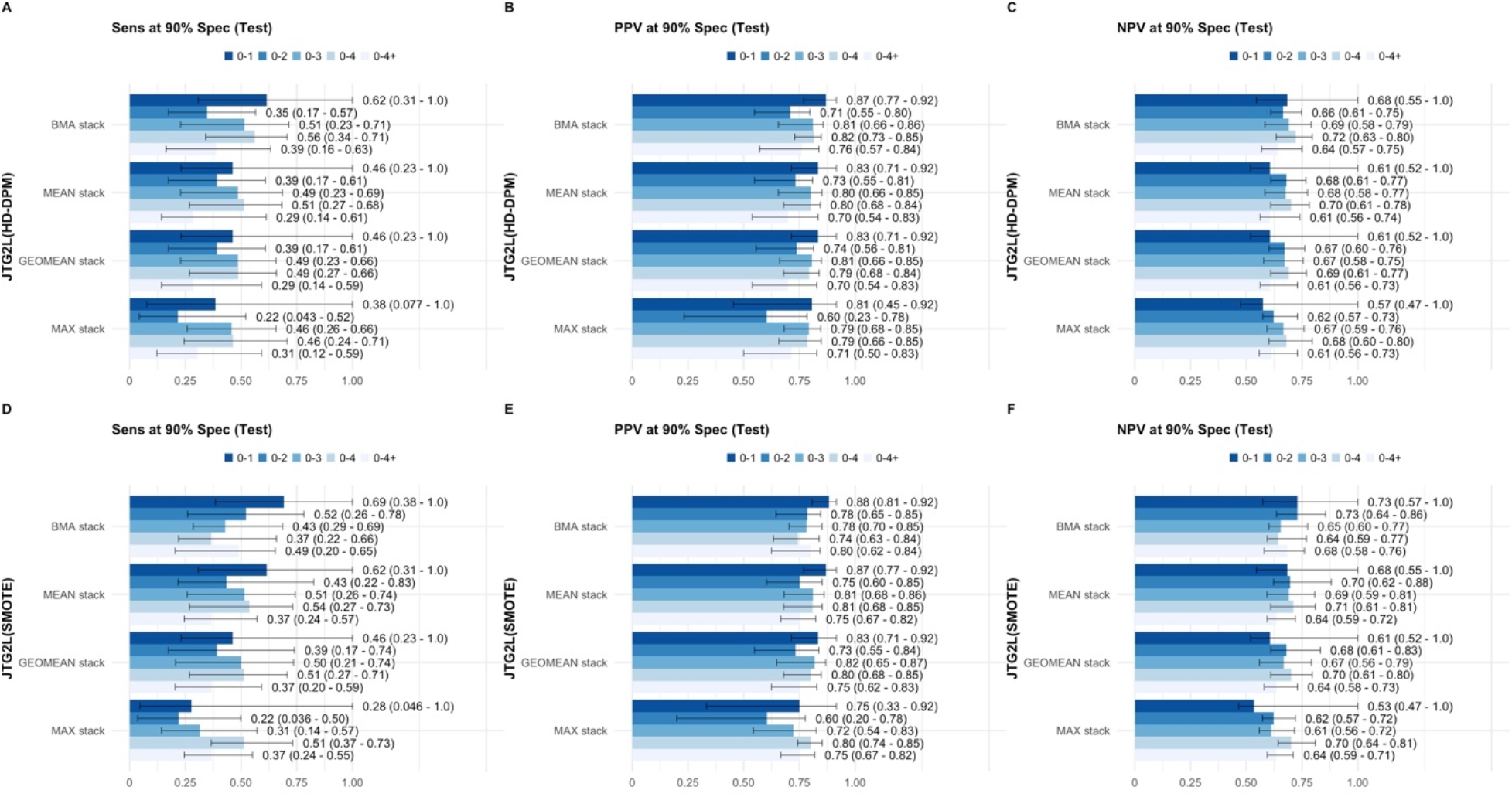
Sensitivity and positive predictive value for meta-learners with HD-DPM and SMOTE during resampling, JTG2L. See Methods for details.

**Fig. S31.**
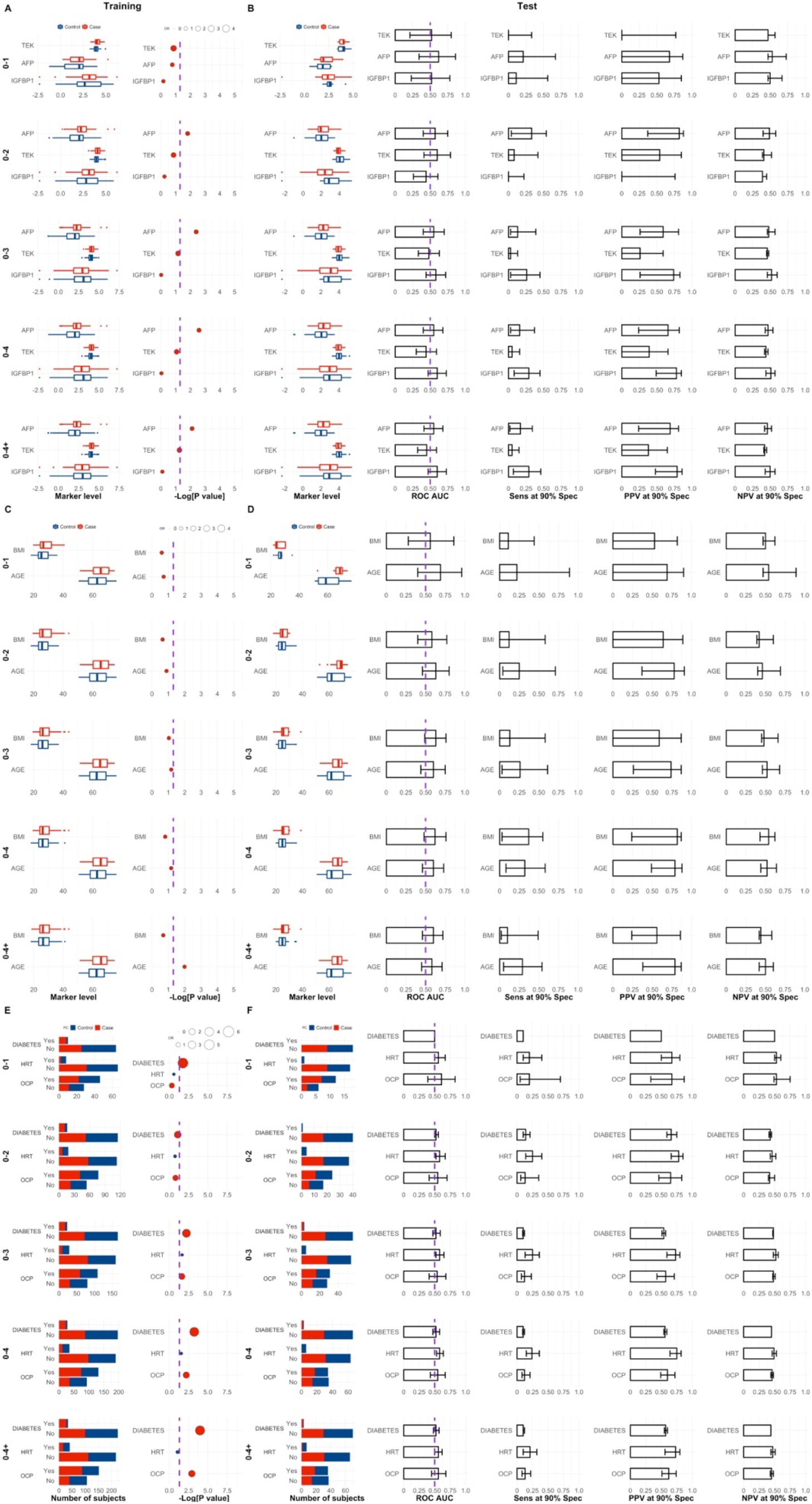
Characteristic proportions for DIABETES, HRT and OCP, and feature ranks in the training set per joined time group according to a logistic regression model with a bias reduction method for a subset of subjects for which AFP, TEK and IGFBP1 was measured.

**Fig. S32.**
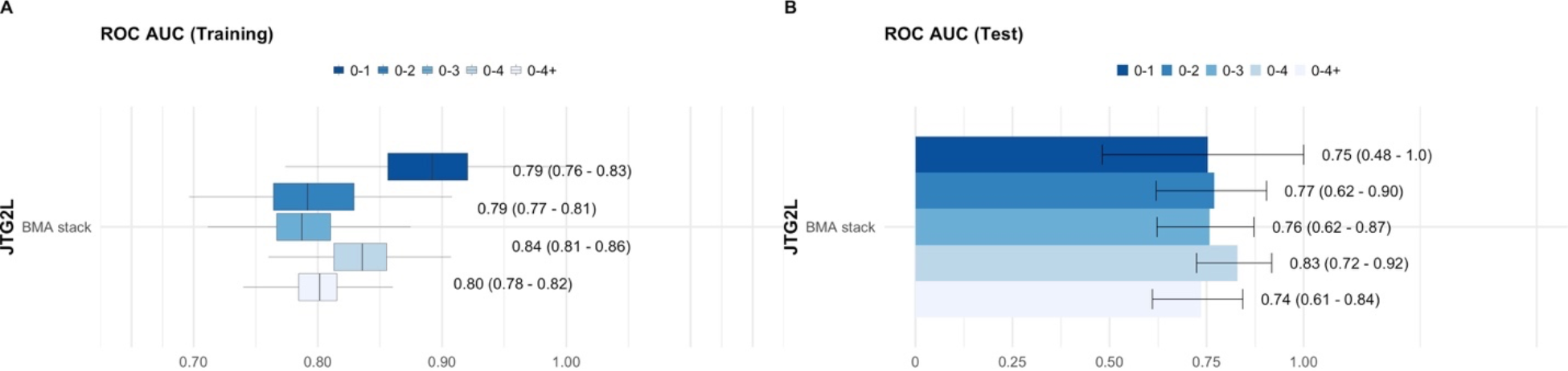
Performance with the BMA meta-learner for a subset of subjects for which AFP, TEK and IGFBP1 was also measured. **A** ROC AUC across training folds for the enhanced JTG2L model**. B** ROC AUC in the test set with the respective model developed in the training set.

**Fig. S33.**
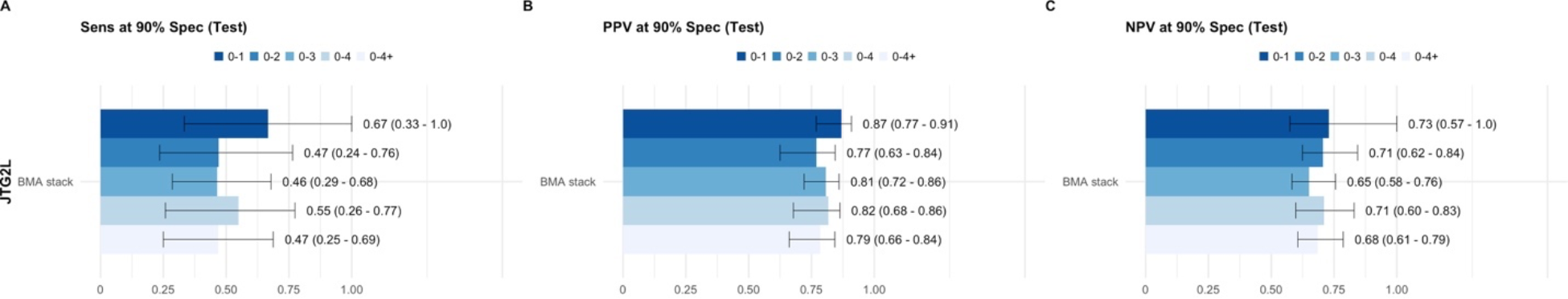
Sensitivity, positive and negative predictive value with the BMA meta-learner for a subset of subjects for which AFP, TEK and IGFBP1 was also measured. **A** Sensitivity, **B** Positive predictive value (PPV) and C Negative predictive value (NPV) at 90% specificity for the enhanced JTG2L model.

**Fig. S34.**
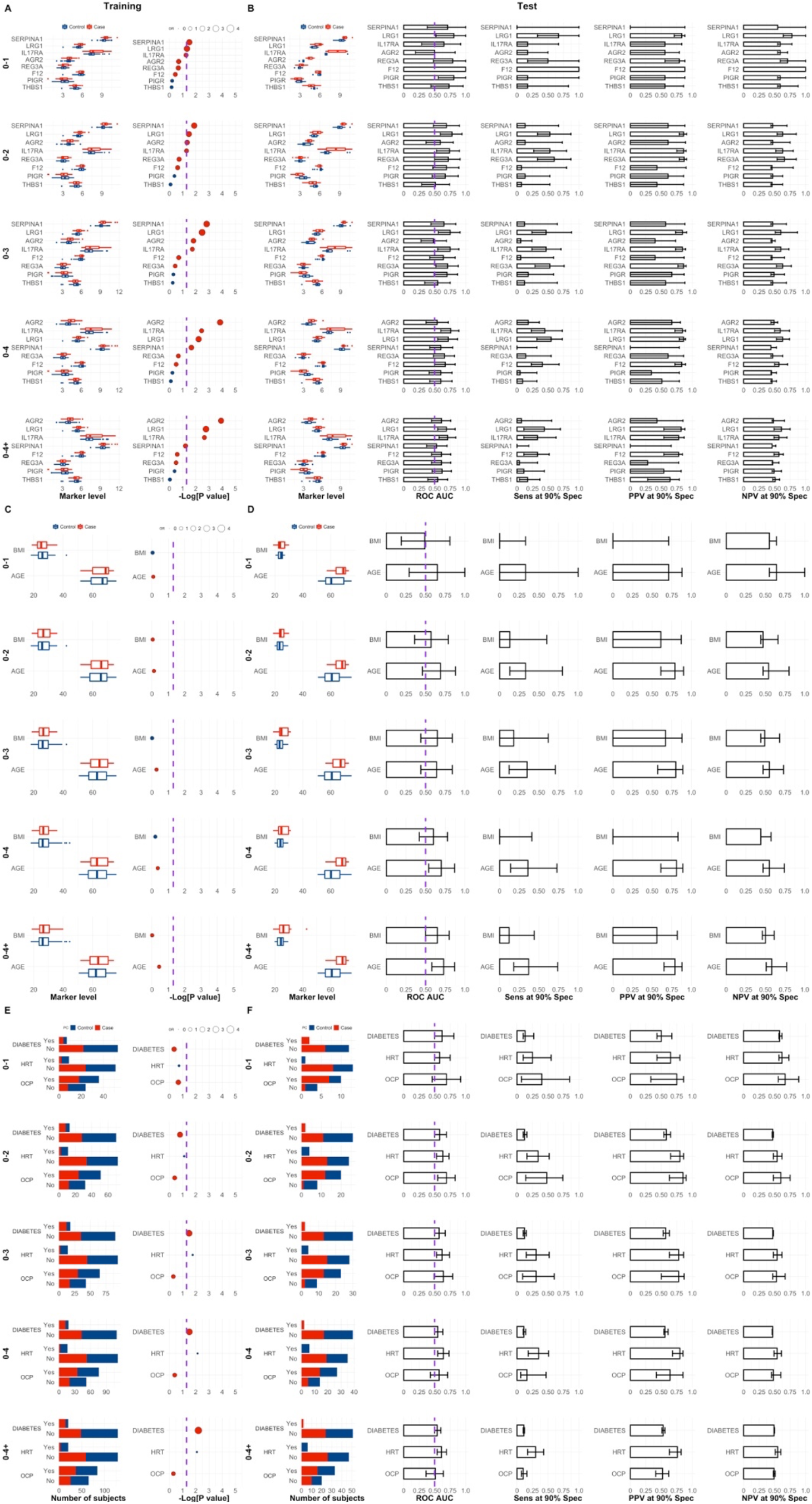
Characteristic proportions for DIABETES, HRT and OCP, and feature ranks in the training set per joined time group according to a logistic regression model with a bias reduction method for a subset of subjects for which LRG1, PIGR, REG3A, F12, AGR2, IL17RA, SERPINA1, THBS1 was measured.

**Fig. S35.**
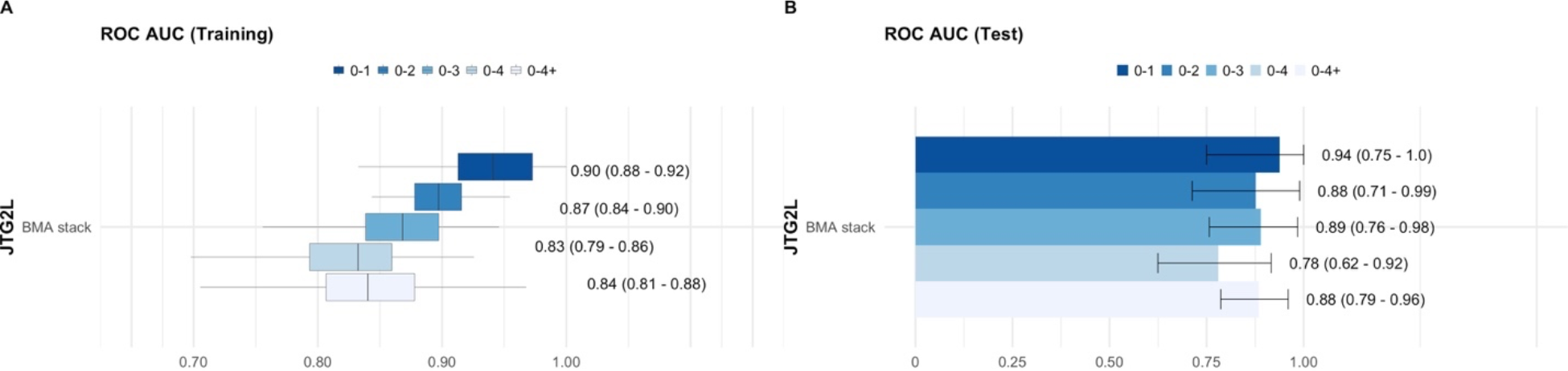
Performance with the BMA meta-learner for a subset of subjects for which LRG1, PIGR, REG3A, F12, AGR2, IL17RA, SERPINA1 and THBS1 were also measured. **A** ROC AUC across training folds for the enhanced JTG2L model. **B** ROC AUC in the test set with the respective model developed in the training set.

**Fig. S36.**
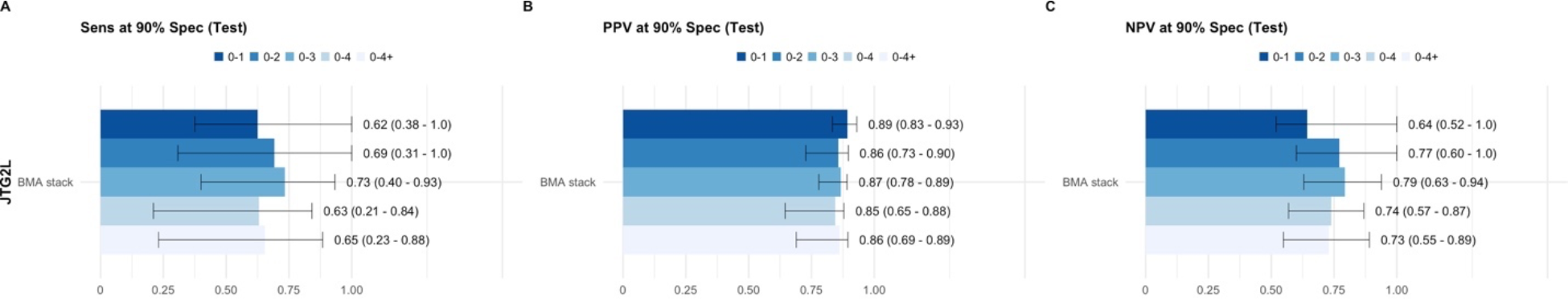
Sensitivity, positive and negative predictive value with the BMA meta-learner for a subset of subjects for which LRG1, PIGR, REG3A, F12, AGR2, IL17RA, SERPINA1 and THBS1 were also measured. **A** Sensitivity, **B** Positive predictive value (PPV) and C Negative predictive value (NPV) at 90% specificity for the enhanced JTG2L model.

**Fig. S37.**
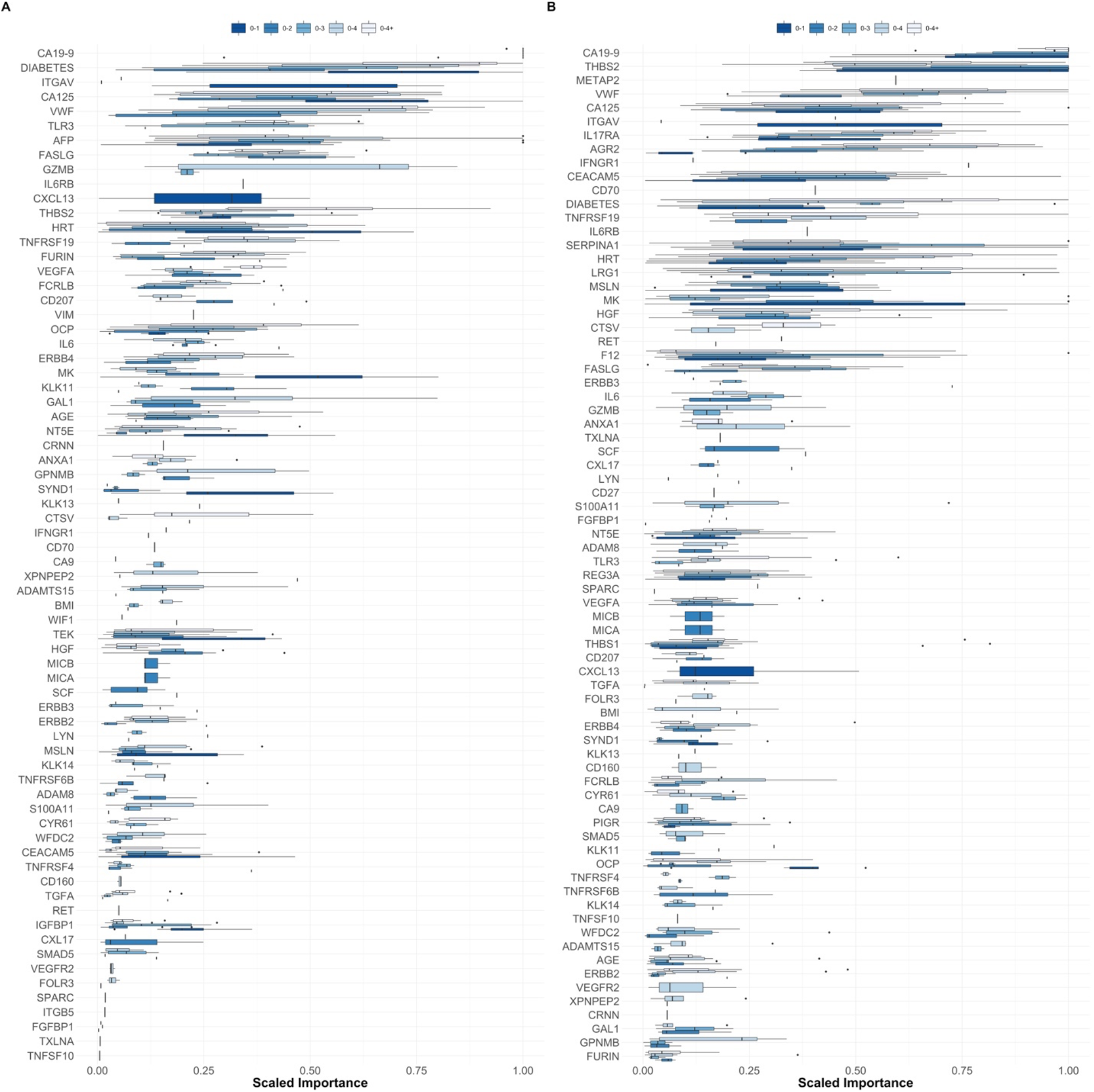
Signatures for each joined time-group. **A** Scaled feature importance across base-learners for the PDAC signature developed in a subset of subjects for which AFP, TEK and IGFBP1 were also measured. **B** Scaled feature importance across base-learners for the PDAC signature developed for a subset of subjects for which LRG1, PIGR, REG3A, F12, AGR2, IL17RA, SERPINA1 and THBS1 were also measured. Feature importance was calculated with a model-agnostic method based on a simple feature importance ranking measure, implemented in the R package *vip* (https://cran.r-roject.org/web/packages/vip/index.html). With oversampling of minority class.

## Supplementary Tables

**Table S1.**
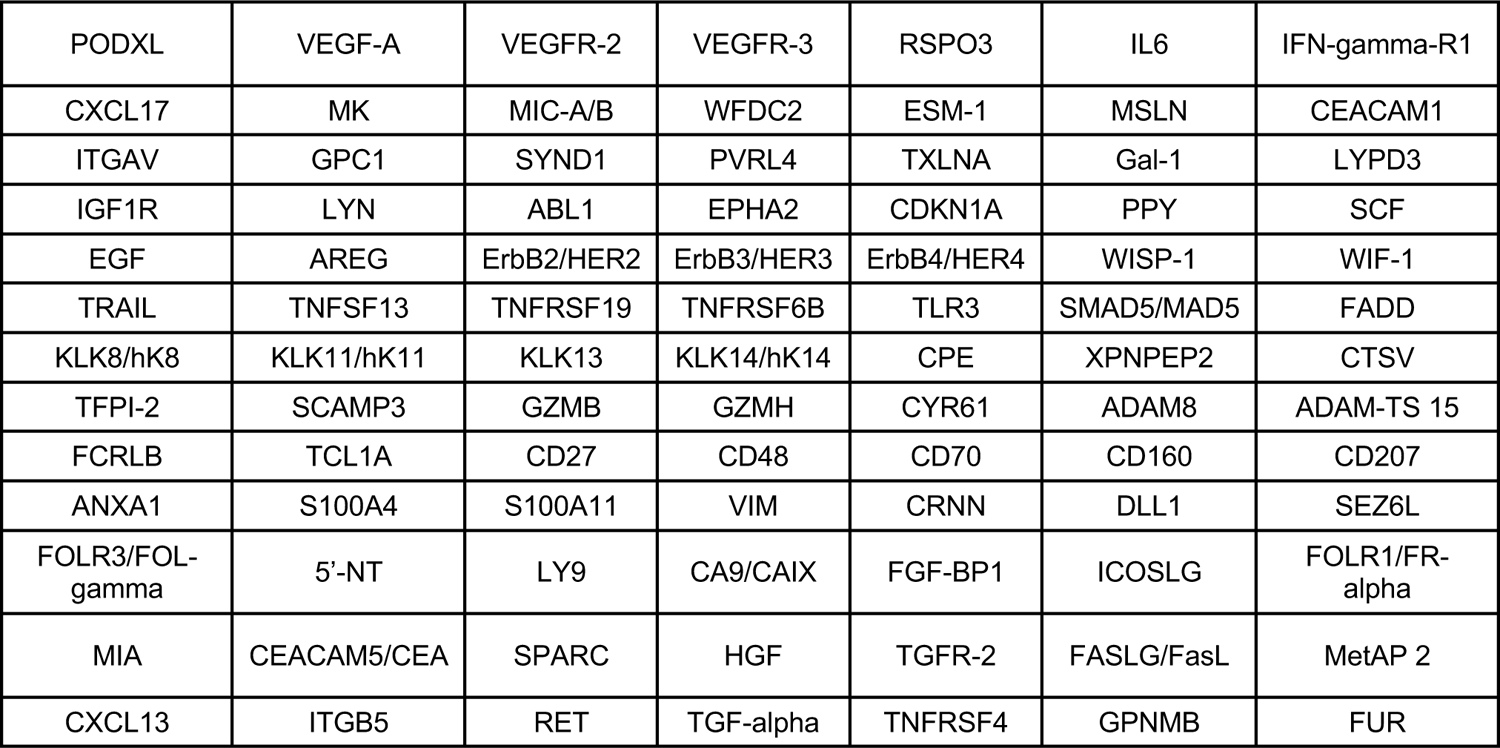
Cancer-associated proteins measured on the Olink Oncology II panel used in this project. The remaining biomarkers were done in-house.

**Table S2.** Training set characteristics. P values were calculated according to a logistic regression model with a bias reduction method (see Methods), for the whole distribution. This table corresponds to taking all samples from all time-groups, i.e., 0 to 4+. See also Fig. S3 and S4. na: not applicable.

| Variable | Cases | Controls | P value |
| --- | --- | --- | --- |
| No. individuals | 117 | 185 |  |
| No. samples | 168 | 185 |  |
| Tumour site |  |  |  |
| tail | 8 | na |  |
| body | 17 | na |  |
| body/tail | 2 | na |  |
| head | 67 | na |  |
| unspecified | 74 | na |  |
| Mean time to spin (h) (range) | 21.64 (4.65 – 46.53) | 21.85 (4.88 - 46.01) | 0.38 |
| Mean age at sample draw (yr) (range) | 64.63 (51.19 - 74.87) | 62.32 (50.44 - 76.08) | 0.0010<br>(OR=1.06) |
| Mean BMI (kg/m2) (range) | 27.83 (18.34 - 43.74) | 27.19 (17.91 – 44.39) | 0.25 |
| Mean time from sample collection to diagnosis (months) (range) | 25.9 (1.02 - 70.09) | na |  |
| HRT use (at randomisation) |  |  |  |
| yes | 18 | 38 | 0.012 |
| no | 150 | 147 | (OR=0.47) |
| OCP use (ever) |  |  |  |
| yes | 106 | 94 | 0.02 |
| no | 63 | 91 | (OR=1.65) |
| Diabetes |  |  |  |
| yes | 38 | 10 | <0.0001 |
| no | 130 | 175 | (OR=4.93) |

**Table S3.** Test set characteristics. P values were calculated according to a logistic regression model with a bias reduction method (see Methods), for the whole distribution. This table corresponds to taking all samples from all time-groups, i.e., 0 to 4+. See also Fig. S3 and S4. na: not applicable.

| Variable | Cases | Controls | P value |
| --- | --- | --- | --- |
| No. individuals | 40 | 64 |  |
| No. samples | 50 | 64 |  |
| Tumour site |  |  |  |
| tail | 5 | na |  |
| body | 2 | na |  |
| body/tail | 1 | na |  |
| head | 21 | na |  |
| unspecified | 21 | na |  |
| Mean time to spin (h) (range) | 22.03 (2 – 46.35) | 21.61 (0.32 – 46.24) | 0.75 |
| Mean age at sample draw (yr) (range) | 65.96 (52.28 - 74.12) | 62.96 (50.59 – 76.86) | 0.0088<br>(OR=1.08) |
| Mean BMI (kg/m2) (range) | 26.21 (17.80 – 42.72) | 25.04 (19.74 - 35.14) | 0.13 |
| Mean time from sample collection to diagnosis (months) (range) | 26.61 (0.99 - 66.07) | n.a. |  |
| HRT use (at randomisation) |  |  |  |
| yes | 1 | 10 | 0.013 |
| no | 49 | 54 | (OR=0.16) |
| OCP use (ever) |  |  |  |
| yes | 26 | 33 | 0.96 |
| no | 24 | 31 |  |
| Diabetes |  |  |  |
| yes | 4 | 1 | 0.11 |
| no | 46 | 63 |  |

**Table S4.** Number of samples per time-group per data set.

| Single time-group | Training set number of PC samples | Training set number of Control samples | Test set number of PC samples | Test set number of Control samples |
| --- | --- | --- | --- | --- |
| 0-1 | 48 | 59 | 13 | 13 |
| 1-2 | 39 | 34 | 11 | 23 |
| 2-3 | 33 | 39 | 12 | 10 |
| 3-4 | 24 | 23 | 6 | 10 |
| 4+ | 24 | 30 | 8 | 8 |

**Table S5.** Subset characteristics within the training set for which AFP, TEK and IGFBP1 were also measured. P values were calculated according to a logistic regression model with a bias reduction method (see Methods), for the whole distribution. This table corresponds to taking all samples from all time-groups, i.e., 0 to 4+. na: not applicable.

| Variable | Cases | Controls | P value |
| --- | --- | --- | --- |
| No. individuals | 89 | 132 |  |
| No. samples | 126 | 132 |  |
| Tumour site |  |  |  |
| tail | 6 | na |  |
| body | 12 | na |  |
| body/tail | 1 | na |  |
| head | 49 | na |  |
| unspecified | 58 | na |  |
| Mean time to spin (h) (range) | 21.38 (0.6 – 46.53) | 21.90 (0.58 – 46.01) | 0.56 |
| Mean age at sample draw (yr) (range) | 64.63 (51.19 – 74.87) | 62.56 (50.44 – 76.08) | 0.01<br>(OR=1.05) |
| Mean BMI (kg/m<sup>2</sup>) (range) | 27.83 (18.34 – 43.74) | 27.03 (41.09 – 17.91) | 0.20 |
| Mean time from sample collection to diagnosis (months) (range) | 26.31 (1.02 – 70.09) | n.a. |  |
| HRT use (at randomisation) |  |  |  |
| yes | 15 | 26 |  |
| no | 111 | 106 | 0.089 |
| OCP use (ever) |  |  |  |
| yes | 87 | 65 |  |
| no | 39 | 67 | 0.0012<br>(OR=2.28) |
| Diabetes |  |  |  |
| yes | 27 | 7 |  |
| no | 99 | 125 | 0.00011<br>(OR=4.62) |

**Table S6.** Subset characteristics within the test set for which AFP, TEK and IGFBP1 were also measured. P values were calculated according to a logistic regression model with a bias reduction method (see Methods), for the whole distribution. This table corresponds to taking all samples from all time-groups, i.e., 0 to 4+. na: not applicable.

| Variable | Cases | Controls | P value |
| --- | --- | --- | --- |
| No. individuals | 25 | 44 |  |
| No. samples | 32 | 44 |  |
| Tumour site |  |  |  |
| tail | 2 | na |  |
| body | 4 | na |  |
| body/tail | 0 | na |  |
| head | 12 | na |  |
| unspecified | 14 | na |  |
| Mean time to spin (h) (range) | 23.75 (3.22 – 46.35) | 21.37 (0.32 - 43.2) | 0.14 |
| Mean age at sample draw (yr) (range) | 65.34 (52.28 – 74.12) | 63.57 (50.62 – 76.86) | 0.26 |
| Mean BMI (kg/m2) (range) | 26.28 (17.8 – 38.70) | 25.33 (19.74 – 35.14) | 0.29 |
| Mean time from sample collection to diagnosis (months) (range) | 21.29 (0.99 - 52.01) | 24.18 (0.99 – 70.09) | 0.40 |
| HRT use (at randomisation) |  |  |  |
| yes | 1 | 6 |  |
| no | 31 | 38 | 0.14 |
| OCP use (ever) |  |  |  |
| yes | 18 | 18 |  |
| no | 14 | 26 | 0.19 |
| Diabetes |  |  |  |
| yes | 2 | 1 |  |
| no | 30 | 43 | 0.40 |

**Table S7.** Number of samples per time-group per data set for which AFP, TEK and IGFBP1 were also measured.

| Single time-group | Training set number of PC samples | Training set number of Control samples | Test set number of PC samples | Test set number of Control samples |
| --- | --- | --- | --- | --- |
| 0-1 | 33 | 41 | 9 | 9 |
| 1-2 | 30 | 27 | 8 | 17 |
| 2-3 | 26 | 32 | 11 | 8 |
| 3-4 | 20 | 16 | 3 | 7 |
| 4+ | 17 | 16 | 1 | 3 |

**Table S8.** Subset characteristics within the training set for which LRG1, PIGR, REG3A, F12, AGR2, IL17RA, SERPINA1 and THBS1 were also measured. P values were calculated according to a logistic regression model with a bias reduction method (see Methods), for the whole distribution. This table corresponds to taking all samples from all time-groups, i.e., 0 to 4+. na: not applicable.

| Variable | Cases | Controls | P value |
| --- | --- | --- | --- |
| No. individuals | 39 | 87 |  |
| No. samples | 62 | 87 |  |
| Tumour site |  |  |  |
| tail | 5 | na |  |
| body | 10 | na |  |
| body/tail | 2 | na |  |
| head | 23 | na |  |
| unspecified | 22 | na |  |
| Mean time to spin (h) (range) | 22.04 (0.48 – 46.40) | 21.56 (0.82 – 45.82) | 0.64 |
| Mean age at sample draw (yr) (range) | 63.81 (51.57 – 74.35) | 62.75 (50.56 – 76.08) | 0.36 |
| Mean BMI (kg/m<sup>2</sup>) (range) | 18.64 (27.44 – 40.06) | 27.43 (17.91 – 44.39) | 0.99 |
| Mean time from sample collection to diagnosis (months) (range) | 22.59 (1.02 – 66.01) | na |  |
| HRT use (at randomisation) |  |  |  |
| yes | 3 | 17 | 0.0083<br>(OR=0.24) |
| no | 59 | 70 |  |
| OCP use (ever) |  |  |  |
| yes | 37 | 47 | 0.50 |
| no | 25 | 15 |  |
| Diabetes |  |  |  |
| yes | 14 | 6 | 0.0064<br>(OR=3.75) |
| no | 48 | 81 |  |

**Table S9.** Subset characteristics within the test set for which LRG1, PIGR, REG3A, F12, AGR2, IL17RA, SERPINA1 and THBS1 were also measured. P values were calculated according to a logistic regression model with a bias reduction method (see Methods), for the whole distribution. This table corresponds to taking all samples from all time-groups, i.e., 0 to 4+. na: not applicable.

| Variable | Cases | Controls | P value |
| --- | --- | --- | --- |
| No. individuals | 20 | 28 |  |
| No. samples | 27 | 28 |  |
| Tumour site |  |  |  |
| tail | 1 | na |  |
| body | 2 | na |  |
| body/tail | 1 | na |  |
| head | 11 | na |  |
| unspecified | 12 | na |  |
| Mean time to spin (h) (range) | 20.09 (2 – 25.84) | 22.09 (4.64 – 46.24) | 0.27 |
| Mean age at sample draw (yr) (range) | 66.85 (56.17 – 73.11) | 61.81 (50.59 – 74.95) | 0.0054<br>(OR=1.12) |
| Mean BMI (kg/m2) (range) | 26.33 (18.64 – 42.72) | 24.35 (19.74 – 29.88) | 0.086 |
| Mean time from sample collection to diagnosis (months) (range) | 29.09 (0.99 – 66.07) | na |  |
| HRT use (at randomisation) |  |  |  |
| yes | 0 | 6 | 0.010 |
| no | 27 | 2 | (OR=0.063) |
| OCP use (ever) |  |  |  |
| yes | 17 | 18 | 0.92 |
| no | 10 | 10 |  |
| Diabetes |  |  |  |
| yes | 2 | 0 | 0.20 |
| no | 25 | 28 |  |

**Table S10.** Number of samples per time-group per data set for which LRG1, PIGR, REG3A, F12, AGR2, IL17RA, SERPINA1 and THBS1 were also measured.

| Single time-group | Training set number of PC samples | Training set number of Control samples | Test set number of PC samples | Test set number of Control samples |
| --- | --- | --- | --- | --- |
| 0-1 | 26 | 34 | 8 | 7 |
| 1-2 | 10 | 13 | 6 | 9 |
| 2-3 | 11 | 14 | 2 | 2 |
| 3-4 | 8 | 12 | 4 | 5 |
| 4+ | 7 | 14 | 7 | 5 |

